# Methods and participant characteristics in the Cancer Risk in Vegetarians Consortium: A cross-sectional analysis across 11 prospective studies

**DOI:** 10.1101/2024.01.15.24301161

**Authors:** Yashvee Dunneram, Jia Yi Lee, Cody Z. Watling, Gary E Fraser, Fayth Miles, Dorairaj Prabhakaran, Krithiga Shridhar, Dimple Kondal, Viswanathan Mohan, Mohammed K. Ali, Kabayam M. Venkat Narayan, Nikhil Tandon, Tammy Y. N. Tong, Tina H.T. Chiu, Ming-Nan Lin, Chin-Lon Lin, Hsin-Chou Yang, Yu-Jen Liang, Darren C. Greenwood, Huaidong Du, Zhengming Chen, Canqing Yu, Maria G. Kakkoura, Gillian K. Reeves, Keren Papier, Sarah Floud, Rashmi Sinha, Linda M. Liao, Erikka Loftfield, Janet E. Cade, Timothy J. Key, Aurora Perez-Cornago

## Abstract

**Background:** The associations of vegetarian diets with risks for site-specific cancers have not been estimated reliably due to the low number of vegetarians in previous studies. Therefore, the Cancer Risk in Vegetarians Consortium was established.

**Objective:** To describe and compare the baseline characteristics between non-vegetarian and vegetarian diet groups and between the collaborating studies.

**Methods:** We harmonised individual-level data from 11 prospective cohort studies in the UK, US, India, China, and Taiwan. Comparisons of food intakes, sociodemographic and lifestyle factors were made between diet groups and between cohorts using descriptive statistics.

**Results:** 2.3 million participants were included; 66% women and 34% men, with mean ages at recruitment of 57 (SD: 7.8) and 57 (8.6) years, respectively. There were 2.1 million meat eaters, 60,903 poultry eaters, 44,780 pescatarians, 81,165 vegetarians, and 14,167 vegans. Food intake differences between the diet groups varied across the cohorts; for example, fruit and vegetable intakes were generally higher in vegetarians than in meat eaters in all the cohorts except in China. BMI was generally lower in vegetarians, particularly vegans, except for the cohorts in India and China. In general, but with some exceptions, vegetarians were also more likely to be highly educated and physically active and less likely to smoke. In the available resurveys, stability of diet groups was high in all the cohorts except in China.

**Conclusions:** Food intakes and lifestyle factors of both non-vegetarians and vegetarians varied markedly across the individual cohorts, which may be due to differences in both culture and socioeconomic status, as well as differences in questionnaire design. Therefore, care is needed in the interpretation of the impacts of vegetarian diets on cancer risk.

## 1. Introduction

Cancer is a leading cause of death and an important barrier to increasing life expectancy throughout the world (1). The World Cancer Research Fund has estimated that between 30 and 50% of all cancer cases might be preventable by following a healthy lifestyle, including following a healthy diet (2). However, the effects of diet, and more specifically dietary patterns, on cancer risk are not fully understood (3).

Diets that exclude red and processed meat, such as vegetarian and vegan diets, may influence cancer risk because of the omission of meat, which has been associated with a higher risk of colorectal cancer and/or because of their higher content of plant foods and associated nutrients (e.g. dietary fibre) (3, 4). In Western cohorts, the risks of cancer in vegetarians and vegans compared to meat eaters have been reported from several large cohorts; the Adventist Health Study-2 (5), EPIC-Oxford (6), the Oxford Vegetarian study (7), the UK Women’s Cohort Study (8), the Netherlands Cohort Study - Meat Investigation Cohort (9), and UK Biobank (10). While some individual studies have identified significant associations the overall findings for specific cancer sites remain inconclusive, which may be partly due to the small number of cancer cases and/or the limited number of vegetarians included in these individual studies. For example, a lower risk of prostate cancer was found among vegetarians in UK Biobank (10) but not in the Adventist Health Study-2 (5) or EPIC-Oxford (6).

Therefore, to enhance the statistical power to assess the associations of vegetarian diets with the risks of site-specific cancers, we harmonised data from pre-existing prospective cohort studies with a substantial number of vegetarians to establish the Cancer Risk in Vegetarians Consortium. The primary aim of the current paper is to describe the data harmonisation methods and to characterise differences between diet groups (meat eaters, poultry eaters, pescatarians, lacto-ovo vegetarians, lacto vegetarians, ovo vegetarians and vegans) and populations included in the consortium. For this, we describe and compare the dietary, demographic, lifestyle, and anthropometric characteristics of the participants in the 11 contributing cohorts. The secondary aim is to describe the consistency in diet groups over time by comparing the diet group at baseline to the diet group at resurvey for individuals in those cohorts with information on dietary intakes during the follow-up period.

## 2. Methods

### 2.1 Study-level inclusion criteria

Studies were identified from literature searches using PubMed and Web of Science, reference lists from published papers, and discussions with colleagues. Studies were considered eligible if available publications suggested that the cohorts were likely to meet the following criteria: 1) the cohort had targeted recruitment to include a high proportion of vegetarians (typically >25%), or the cohort was very large with ≥500,000 participants and was therefore likely to include ∼5,000 vegetarians (assuming that ∼1% of many populations may be vegetarian); 2) the cohort had reliable follow-up for cancer occurrence (e.g. linkage to cancer registry, medical records, verbal autopsy).

### 2.2 Participating prospective studies and data collection

In total, 11 prospective studies were identified as likely to meet our criteria, and their principal investigators were invited to collaborate and contribute data to this consortium; all accepted. Seven cohorts had a large proportion of vegetarians: the Adventist Health Study-2 (11), the Center for cArdiometabolic Risk Reduction in South Asia-1 (CARRS-1) (12, 13), CARRS-2 (13), EPIC-Oxford (14), the Oxford Vegetarian Study (15), the Tzu Chi Health Study (16) and the UK Women’s Cohort Study (17). Four cohorts were very large (≥500,000 participants) and thus included a substantial number of vegetarians: the China Kadoorie Biobank (18), the Million Women Study (19), the National Institutes of Health-AARP Diet and Health Study (NIH-AARP) (20), and the UK Biobank (21). With reference to our aim to identify cohorts expected to include a large proportion and/or number of vegetarians, the harmonised data showed that three cohorts did not reach these targets: CARRS-1 and the UK Women’s Cohort Study had substantial proportions of vegetarians and vegans, but somewhat less than the target of 25% (23% and 14% respectively), while the NIH-AARP had only 1,943 vegetarians and vegans (0.4% of the whole cohort). These cohorts were retained in the consortium nevertheless because they provide substantial information on vegetarians in diverse settings.

Each study had ethical approval covering the current research topic, and participants in each study gave consent at the time of recruitment. **Table 1** and the Supplementary Methods provide a brief description of these 11 studies, while further details can be found in the original publications from the individual studies (11–21).

**Table 1.** Characteristics of the cohorts included in the Cancer Risk in Vegetarians Consortium (n = 2,337,152)

| Cohort | Location | Recruitment years | Baseline sample size<sup>1</sup> | Eligible age range at recruitment (years) | No. of foods assessed in baseline dietary questionnaire | Dietary resurvey years | Follow-up time to cancer incidence (years), median (IQR) |
| --- | --- | --- | --- | --- | --- | --- | --- |
| *Cohorts with large proportions of vegetarians* |  |  |  |  |  |  |  |
| Adventist Health Study-2 (11) | United States | 2002–2007 | 64,555 | ≥30 | 130 | - | 8 (7–10) |
| CARRS-1 (12) | India | 2010–2011 | 12,218 | ≥20 | 26 | 2017–2018 | 6 (5–6) |
| CARRS-2 (12, 13) | India | 2014–2016 | 9,530 | ≥20 | 33 | - | 4 (4–4) |
| EPIC-Oxford (14) | United Kingdom | 1993–1999 | 53,752 | ≥20 | 130 | 2010 | 22 (21–23) |
| Oxford Vegetarian Study (15) | United Kingdom | 1980–1984 | 10,527 | ≥15 | 28 | 1993–1999<sup>2</sup> | 27 (14–37) |
| Tzu Chi Health Study (16) | Taiwan | 2007–2009 | 5,520 | ≥18 | 64 | - | 11 (10–12) |
| UK Women’s Cohort Study (17) | United Kingdom | 1995–1998 | 30,148 | 35–69 | 217 | 2000 | 20 (19–21) |
| *Very large cohorts* |  |  |  |  |  |  |  |
| China Kadoorie Biobank (18) | China | 2004–2008 | 510,145 | 30–79 | 12 | 2013–2014 | 12 (11–13) |
| Million Women Study (19) | United Kingdom | 1996–2001 | 639,026 | 50–64 | 130 | 2010–2020 | 16 (14–18) |
| NIH-AARP (20) | United States | 1995–1996 | 527,691 | 50–69 | 124 | - | 16 (10–16) |
| UK Biobank (21) | United Kingdom | 2006–2010 | 474,040 | 40–69 | 16 | 2012–2013 | 12 (11–12) |

Abbreviations: CARRS, Centre for cArdiometabolic Risk Reduction in South Asia; EPIC, European Prospective Investigation into Cancer and Nutrition; FFQ, food frequency questionnaire; NIH-AARP, National Institutes of Health-AARP Diet and Health Study.
<sup>1</sup>The baseline sample size corresponds to the number of participants in the pooled dataset after exclusions as listed in Figures S1–S11.
<sup>2</sup>There are 2,837 participants in the Oxford Vegetarian Study who also took part in EPIC-Oxford, therefore the baseline survey in EPIC-Oxford is considered as a resurvey for this subsample of Oxford Vegetarian Study participants.

A detailed data dictionary including the variables requested and their required format was circulated to the collaborators (Table S1). Individual participant data were requested for dietary intake, which is described below. Date of birth or date and age at recruitment, as well as socio-demographics, lifestyle factors, medical history, women-specific data, vitamin and mineral supplement use, were requested if available. Follow-up dietary data (as described below) were also requested, where available. Additionally, data on cancer incidence and death (i.e. site and histology of the tumour, date of cancer diagnosis, date of death, hormone receptor status for breast cancer, stage and grade of tumour for prostate cancer) were requested.

In addition to the above variables, data dictionaries, survey questionnaires, and coding algorithms for derived variables were also requested. Data transfer agreements were signed with the collaborating institutions before transferring the individual participant data to the University of Oxford. Due to the law in Taiwan, individual participant data from the Tzu Chi Health Study (after linkage to cancer registry) could not be transferred to Oxford, and instead the collaborators conducted the statistical analyses at the Health and Welfare Data Science Center (HWDC), Ministry of Health, Taiwan and provided us with the tabulations of results (i.e. no individual data were shared).

### 2.3 Food intake assessment

In all the studies, a dietary questionnaire (mainly food frequency questionnaires, FFQ) was used to assess the food intake of participants at baseline (the number of foods assessed by each dietary questionnaire can be found in **Table 1**). The baseline dietary assessment tools in most of the studies were validated for key dietary components in the relevant study population or a similar population (16, 22–26). After the data were transferred to the University of Oxford, food frequencies were converted into weights in grams/day (g/d) using standard portion sizes specific to the study’s population (see Supplementary Methods for further details). Table S2 lists the foods for which data on food intake were requested from the individual studies. Intakes of energy and nutrients, as calculated by the collaborators for each study, were also requested and provided by the Adventist Health Study-2, EPIC-Oxford, UK Women’s Cohort Study, Million Women Study, and NIH-AARP.

Resurvey dietary data were available for a subsample of participants from the baseline cohort in seven out of the eleven cohorts: CARRS-1 (n = 7,389; 61% of the whole cohort), EPIC-Oxford (n = 27,529; 51%), Oxford Vegetarian Study (n = 2,837; 27%; these are participants who also took part in EPIC-Oxford, therefore the baseline survey in EPIC-Oxford is considered as a resurvey for this subsample of Oxford Vegetarian Study participants), UK Women’s Cohort Study (n = 1,652; 5%), China Kadoorie Biobank (n = 24,586; 5%), Million Women Study (n = 23,525; 4%) and UK Biobank (n = 17,426, 4%), which were used to examine whether participants in each diet group changed their food intakes over time. Further details on the repeat dietary assessments are available in the Supplementary Methods.

### 2.4 Classification of diet groups

The consumption frequencies of animal foods (i.e., pork, beef, lamb, poultry, fish, eggs, dairy products) or dishes containing animal foods were used to classify participants into one of the seven diet groups defined *a priori* (**Table 2**). Meat eaters were defined as participants who consumed red and/or processed meat. To categorise participants as poultry eaters, pescatarians, vegetarians, or vegans, the lowest consumption frequency of the animal food not consumed by the respective diet group was used to define absence of that food from the diet (Table S3). For example, in the Adventist Health Study-2 and the China Kadoorie Biobank, lacto-ovo vegetarians were defined as those who reported consuming dairy products and eggs but reported “never/rarely” for their consumption of red and processed meat, poultry, and fish, because the option “never” was not available on the questionnaires. At follow-up, we also classified the subsamples of participants with follow-up dietary data into the diet groups as defined in **Table 2** to assess the proportion of participants who had changed diet groups over median follow-up periods ranging between 2.5 and 14.3 years. Further details on the diet group classification in each study can be found in the Supplementary Methods.

**Table 2.** *A priori* ideal definitions of the diet groups.

| **Diet group** | **Definition** |
| --- | --- |
| Meat eaters | Consume red and processed meat |
| Poultry eaters | Do not consume red or processed meat but consume poultry |
| Pescatarians | Do not consume meat or poultry but consume fish |
| Lacto-ovo vegetarians | Do not consume meat, poultry, or fish, but consume eggs and dairy products |
| Lacto vegetarians | Do not consume meat, poultry, fish, or eggs, but consume dairy products |
| Ovo vegetarians | Do not consume meat, poultry, fish, or dairy products but consume eggs |
| Vegans | Do not consume meat, poultry, fish, dairy products, or eggs |

### 2.5 Data harmonisation

After receiving the datasets from the individual cohorts, each variable was examined to identify outliers and inconsistencies; in such cases, the study collaborators were consulted and a decision was agreed upon to resolve the issue. For physical activity, cut-off points of the metabolic equivalent of task (MET)–hours per day specific to each population were used to categorize participants as inactive, moderately active or highly active in most of the studies. In the Oxford Vegetarian Study, educational status had not been assessed and therefore socio-economic status was used instead as a proxy. A detailed description of the data harmonisation process is available in the Supplementary Methods.

### 2.6 Participant-level exclusions

Prior to the data harmonisation process, participants were excluded from individual studies based on specific criteria unique to each study (referred to as “initial exclusions” in Figures S1-S11). The following exclusions were made across all studies: 1) participants with prevalent invasive cancers as identified by the cancer registry or self-report (CARRS-1 and CARRS-2); 2) participants who could not be linked to a cancer registry (except for CARRS-1 and CARRS-2); 3) those with no follow-up data; 4) participants with more than 80% missing dietary data; 5) those who reported implausible energy intakes (male: <3,347 or >16,736 kJ/day; female: <2,092 or >14,644 kJ/day) (if such data were available) (27); and 6) those above the age of 89 years at baseline. After applying the above exclusions, a total of 2,337,152 participants were included in the analyses.

### 2.7 Statistical analysis

The numbers (and percentages) of participants in each diet group are presented by cohort. The baseline demographic and lifestyle characteristics of the participants by sex and by diet group are presented as means and standard deviations (SD) for continuous variables and numbers and percentages for categorical variables across the cohorts. The women-specific characteristics of the cohorts are also presented.

The food intakes of major food groups by cohort within each diet group are also presented as means (SDs). In the results section, cohorts are ordered by their design (distinction was made between cohorts with large proportions of vegetarians and very large cohorts) as both types of cohorts have inherently different recruitment strategies which could lead to differing participant characteristics.

## 3. Results

### Baseline and demographic characteristics

The consortium comprises data from 2,337,152 participants, including 81,165 vegetarians and 14,167 vegans (**Tables 3** and S4). Moreover, 1,546,217 (66%) of the participants were women and 790,935 (34%) were men, with mean (SD) ages of 56.9 (7.8) years and 57.3 (8.6) years at recruitment, respectively (**Table 4**). Two studies, the UK Women’s Cohort Study and the Million Women Study, recruited women only. In the UK cohorts and NIH-AARP, ≥90% of participants were of white European ancestry, whereas in the Adventist Health Study-2, 27.6% of women and 20.9% of men were of African American or Caribbean ethnicity (classified as Black) (**Table 5**). Women-specific characteristics are shown in **Table 6** (and with a more detailed categorisation in Table S5). Further information on the baseline demographic characteristics of the participants can be found in **Table 4**, with detailed categorisations shown in Tables S4, S5 and S6.

**Table 3.** Number of participants in each diet group (n = 2,337,152)

| Cohort | Meat eaters | Poultry eaters | Pescatarians | Vegetarians |  |  |  | Vegans |
| --- | --- | --- | --- | --- | --- | --- | --- | --- |
|  |  |  |  | Lacto-ovo vegetarians | Lacto vegetarians | Ovo vegetarians | All vegetarians<sup>2</sup> |  |
| *Cohorts with large proportions of vegetarians* |  |  |  |  |  |  |  |  |
| Adventist Health Study-2 | 23,245 (36.0) | 10,494 (16.3) | 6,202 (9.6) | 12,798 (19.8) | 5,915 (9.2) | 676 (1.0) | 19,389 (30.0) | 5,225 (8.1) |
| CARRS-1 | 7,214 (59.0) | 1,958 (16.0) | 180 (1.5) | 365 (3.0) | 1,920 (15.7) | 129 (1.1) | 2,414 (19.8) | 452 (3.7) |
| CARRS-2 | 4,786 (50.2) | 1,938 (20.3) | 167 (1.8) | 323 (3.4) | 2,061 (21.6) | 47 (0.49) | 2,431 (25.5) | 208 (2.2) |
| EPIC-Oxford | 26,498 (49.3) | 1,703 (3.2) | 8,128 (15.1) | 11,390 (21.2) | 3,948 (7.3) | 95 (0.18) | 15,433 (28.7) | 1,990 (3.7) |
| Oxford Vegetarian Study<sup>3</sup> | 5,180 (49.2) | - | 998 (9.5) | 3,576 (34.0) | 388 (3.7) | 52 (0.49) | 4,016 (38.1) | 333 (3.2) |
| Tzu Chi Health Study | 3,519 (63.8) | 120 (2.2) | 233 (4.2) | 1,277 (23.1) | 234 (4.2) | 93 (1.7) | 1,604 (29.1) | 44 (0.80) |
| UK Women’s Cohort Study | 21,180 (70.3) | 929 (3.1) | 3,804 (12.6) | 3,300 (10.9) | 743 (2.5) | 32 (0.11) | 4,075 (13.5) | 160 (0.53) |
| *Very large cohorts* |  |  |  |  |  |  |  |  |
| China Kadoorie Biobank | 485,796 (95.2) | 1,949 (0.38) | 1,703 (0.33) | 2,452 (0.48) | 730 (0.14) | 12,405 (2.4) | 15,587 (3.1) | 5,110 (1.0) |
| Million Women Study | 618,450 (96.8) | 3,414 (0.53) | 10,947 (1.7) | 5,361 (0.84) | 742 (0.12) | 33 (0.01) | 6,136 (0.96) | 79 (0.01) |
| NIH-AARP | 491,098 (93.1) | 32,960 (6.2) | 1,690 (0.32) | 969 (0.18) | 842 (0.16) | 14 (0.00) | 1,825 (0.35) | 118 (0.02) |
| UK Biobank | 449,171 (94.8) | 5,438 (1.1) | 10,728 (2.3) | 7,140 (1.5) | 940 (0.20) | 175 (0.04) | 8,255 (1.7) | 448 (0.09) |
| All cohorts combined | 2,136,137 (91.4) | 60,903 (2.6) | 44,780 (1.9) | 48,951 (2.1) | 18,463 (0.79) | 13,751 (0.59) | 81,165 (3.5) | 14,167 (0.61) |

Abbreviations: CARRS, Centre for cArdiometabolic Risk Reduction in South Asia; EPIC, European Prospective Investigation into Cancer and Nutrition; NIH-AARP, National Institutes of Health-AARP Diet and Health Study.
<sup>1</sup>Values are N (%). Categorisation into diet groups necessarily used the questions asked in each study, therefore some participants in the vegetarian categories may have very low but not zero intakes of meat, and similarly for the other animal foods.
<sup>2</sup>All vegetarians include lacto-ovo, lacto and ovo vegetarians.
<sup>3</sup>In the Oxford Vegetarian Study, poultry eaters could not be determined as poultry intake was not assessed.

**Table 4:**
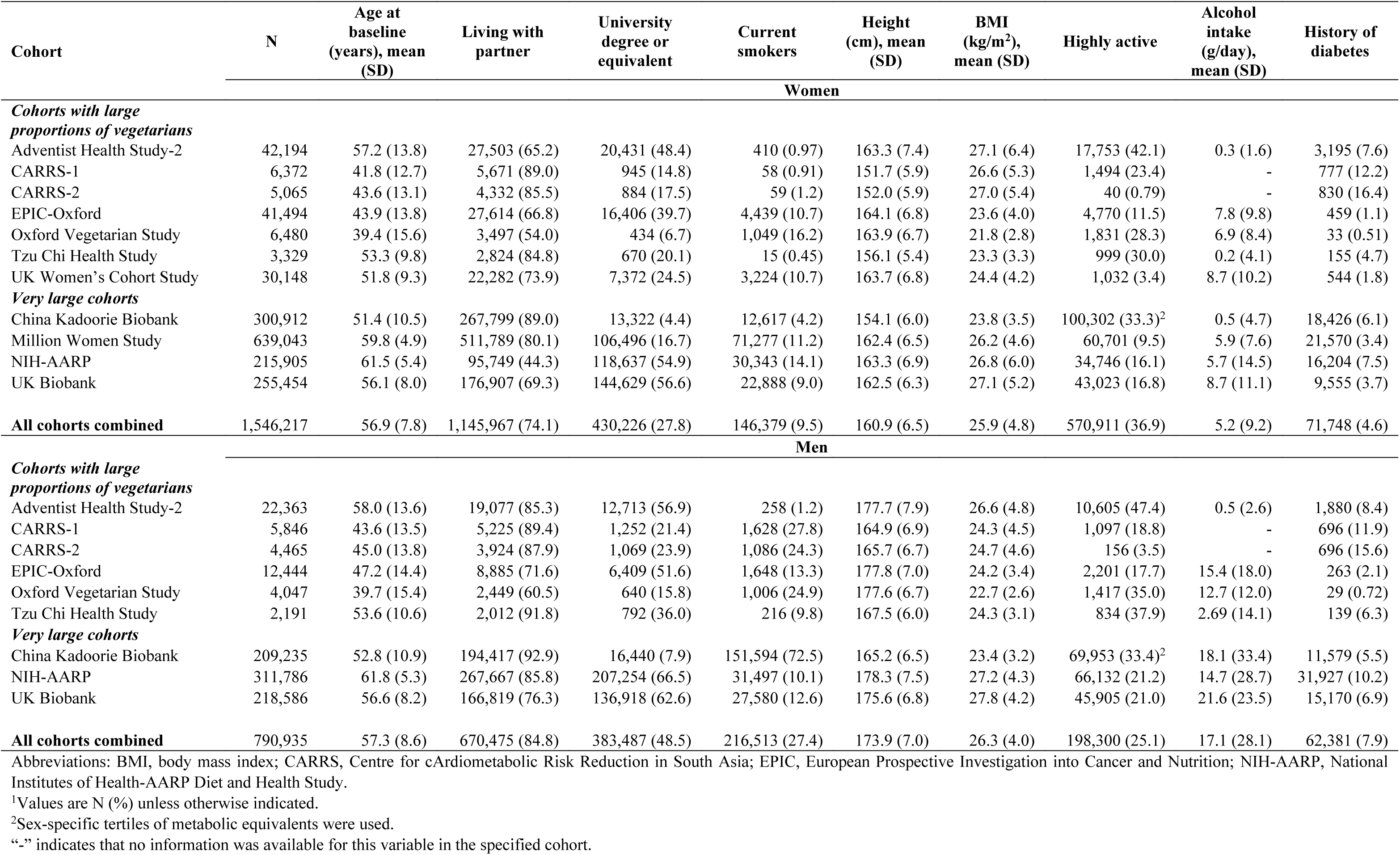
Baseline and demographic charecteristiscs of participants by sex and cohort (n = 2,337,152)

**Table 5:** Ethnicity of participants by sex and cohort (n = 2,337,152)

| Cohort | White | Black | Hispanic | Asian<sup>2</sup> | Other |
| --- | --- | --- | --- | --- | --- |
| Women (n = 1,546,217) |  |  |  |  |  |
| *Cohorts with large proportions of vegetarians* |  |  |  |  |  |
| Adventist Health Study-2 | 26,727 (63.3) | 11,632 (27.6) | 1,878 (4.5) | 83 (0.20) | 1,874 (4.4) |
| CARRS-1 | - | - | - | 6,372 (100)<sup>3</sup> | - |
| CARRS-2 | - | - | - | 5,065 (100)<sup>3</sup> | - |
| EPIC-Oxford | 39,701 (96.0) | 149 (0.36) | - | 221 (0.53) | 357 (0.86) |
| Oxford Vegetarian Study | 6,480 (100)<sup>3</sup> | - | - | - | - |
| Tzu Chi Health Study | - | - | - | 3,329 (100)<sup>3</sup> | - |
| UK Women’s Cohort Study | 28,953 (96.0) | 41 (0.14) | - | 165 (0.55) | 179 (0.59) |
| *Very large cohorts* |  |  |  |  |  |
| China Kadoorie Biobank | - | - | - | 300,909 (100)<sup>3</sup> | - |
| Million Women Study | 627,121 (98.1) | 1,112 (0.17) | - | 1,814 (0.28) | 2,004 (0.31) |
| NIH-AARP | 194,213 (90.0) | 11,280 (5.2) | 3,917 (1.8) | 2,355 (1.1) | 960 (0.44) |
| UK Biobank | 240,675 (94.2) | 4,492 (1.8) | - | 5,336 (2.1) | 4,220 (1.7) |
| All cohorts combined<sup>4</sup> | 1,163,870 (75.3) | 28,706 (1.9) | 5,795 (0.37) | 325,649 (21.1) | 9,594 (0.62) |
| Men (n = 790,935) |  |  |  |  |  |
| *Cohorts with large proportions of vegetarians* |  |  |  |  |  |
| Adventist Health Study-2 | 15,763 (70.5) | 4,671 (20.9) | 888 (4.0) | 79 (0.35) | 960 (4.3) |
| CARRS-1 | - | - | - | 5,846 (100)<sup>3</sup> | - |
| CARRS-2 | - | - | - | 4,465 (100)<sup>3</sup> | - |
| EPIC-Oxford | 11,845 (95.4) | 32 (0.26) | - | 102 (0.82) | 171 (1.4) |
| Oxford Vegetarian Study | 4,047 (100)<sup>3</sup> | - | - | - | - |
| Tzu Chi Health Study | - | - | - | 2,191 (100)<sup>3</sup> | - |
| *Very large cohorts* |  |  |  |  |  |
| China Kadoorie Biobank | - | - | - | 209,236 (100)<sup>3</sup> | - |
| NIH-AARP | 290,102 (93.0) | 7,786 (2.5) | 5,618 (1.8) | 3,915 (1.3) | 1,069 (0.34) |
| UK Biobank | 205,686 (94.1) | 3,251 (1.5) | - | 5,713 (2.6) | 2,986 (1.4) |
| All cohorts combined<sup>4</sup> | 527,443 (66.7) | 15,740 (2.0) | 6,506 (0.82) | 231,547 (29.3) | 5,186 (0.66) |

Abbreviations: CARRS, Centre for cArdiometabolic Risk Reduction in South Asia; EPIC, European Prospective Investigation into Cancer and Nutrition; NIH-AARP, National Institutes of Health-AARP Diet and Health Study.
<sup>1</sup>Values are N (%).
<sup>2</sup>Asian in CARRS-1 and CARRS-2 refers to South Asians, while Asian in China Kadoorie Biobank and Tzu Chi Health Study refers to East Asians.
<sup>3</sup>All participants in CARRS-1, CARRS-2, China Kadoorie Biobank and Tzu Chi Health Study were considered to be Asians and all participants in the Oxford Vegetarian Study were considered to be White Europeans.
<sup>4</sup>These numbers do not add up to the total number stated above because some participants have missing ethnicity data which is not shown in this table.
“-” indicates that no information was available for this ethnic group in the specified cohort.

**Table 6.** Women-specific characteristics by cohort (n = 1,546,217)

| Cohort | Age at menarche<br>≤12 years | Parous | Age at first birth<br>≥25 years | Postmenopausal | Age at menopause<br>≥50 years<sup>2</sup> | Ever used oral<br>contraceptive | Ever used<br>HRT |
| --- | --- | --- | --- | --- | --- | --- | --- |
| *Cohorts with large proportions of vegetarians* |  |  |  |  |  |  |  |
| Adventist Health Study-2 | 51.4 | 83.1 | 31.7 | 77.0 | 64.5 | 58.8 | 39.4 |
| CARRS-1 | 12.2 | 94.1 | - | 36.9 | 19.7 | 3.5 | - |
| CARRS-2 | 12.9 | 93.8 | - | 41.9 | 23.4 | 6.9 | - |
| EPIC-Oxford | 39.8 | 61.0 | 38.2 | 40.2 | 54.8 | 73.7 | 18.2 |
| Oxford Vegetarian Study | - | 47.5 | 29.4 | 26.6 | - | 55.3 | - |
| Tzu Chi Health Study | 8.6 | 91.8 | - | 63.0 | 54.2 | 15.7 | 19.6 |
| UK Women's Cohort Study | 41.6 | 77.2 | 44.8 | 60.9 | 48.0 | 66.9 | 27.2 |
| *Very large cohorts* |  |  |  |  |  |  |  |
| China Kadoorie Biobank | 5.5 | 98.7 | 32.5 | 57.2 | 43.1 | 9.8 | - |
| Million Women Study | 38.8 | 88.4 | 36.4 | 100 | 56.8 | 61.3 | 53.4 |
| NIH-AARP | 48.6 | 83.7 | 23.5 | 100 | 39.6 | 34.5 | 52.9 |
| UK Biobank | 37.6 | 81.2 | 47.1 | 77.8 | 63.3 | 81.1 | 37.7 |
| **All cohorts combined** | **33.5** | **87.3** | **35.3** | **84.2** | **52.1** | **50.6** | **37.8** |

Abbreviations: CARRS, Centre for cArdiometabolic Risk Reduction in South Asia; HRT, hormone replacement therapy; EPIC, European Prospective Investigation into Cancer and Nutrition; NIH-AARP, National Institutes of Health-AARP Diet and Health Study.
<sup>1</sup>Values are % of women within cohort.
<sup>2</sup>Postmenopausal women only.
“-” indicates that no information was available for this variable in the specified cohort.

### Diet groups at baseline

In total, 2,136,137 (91.4%) participants in this consortium were meat eaters, 60,903 (2.6%) poultry eaters and 44,780 (1.9%) were pescatarians. The largest numbers of vegetarians were in the Adventist Health Study-2 (n = 19,389), EPIC-Oxford (n = 15,433), and the China Kadoorie Biobank (n = 15,587) (**Table 3**). In the UK and US cohorts as well as the Tzu Chi Health Study, vegetarians mostly followed a lacto-ovo vegetarian diet, while in the Indian cohorts a lacto-vegetarian diet was more common. Moreover, the Adventist Health Study-2 (n = 5,225), EPIC-Oxford (n = 1,990), and the China Kadoorie Biobank (n = 5,110) contributed the largest numbers of vegans.

### Food intakes in vegetarians and vegans across cohorts

The food intakes of participants in each diet group are presented across the individual studies in **Tables 7** (all vegetarians combined, not including vegans) and S7 (showing each individual diet group separately). Given that the dietary assessment tools used in the individual studies are not closely comparable (e.g. the number of questions asking about consumption of foods of the same category varied), the differences in mean intakes should be interpreted with caution.

Vegetarians in the Adventist Health Study-2, EPIC-Oxford, and the UK Women’s Cohort Study consumed a higher amount of wholegrains than refined grains in comparison to vegetarians in the other cohorts, while the opposite was observed in the other studies (**Table 7**). The highest total vegetable intakes were among vegetarians in the Tzu Chi Health Study [mean: 520 (SD: 347) g/d], NIH-AARP [471 (317) g/d], and CARRS-1 [449 (203) g/d]. Moreover, vegetarians from the two Indian studies also had the highest mean intakes of legumes. Fresh fruit intake was higher in the UK and US cohorts in comparison to the Asian cohorts, except for the Oxford Vegetarian Study where reported intake was low. For sweets, preserves, cakes and confectionery, the mean intake was highest in vegetarians from EPIC-Oxford and the UK Women’s Cohort Study. Vegetarians in the NIH-AARP had the highest consumption of sugar-sweetened beverages (this variable includes diet versions in some of the studies). Total dairy intake was lowest among vegetarians in the China Kadoorie Biobank [39 (32) g/d] and Tzu Chi Health Study [43 (69) g/d] compared to vegetarians in the other cohorts, who reported more than double the intake.

**Table 7.** Food intakes in all vegetarians combined (not including vegans)^1^ by cohort (n = 81, 165)^2.^

| Cohort | Plant foods, g/day |  |  |  |  |  |  |
| --- | --- | --- | --- | --- | --- | --- | --- |
|  | Refined grains | Wholegrains | Total vegetables | Legumes | Fresh fruit | Nuts and seeds | Sweets, preserves, cakes & confectionery |
| *Cohorts with large proportions of vegetarians* |  |  |  |  |  |  |  |
| Adventist Health Study-2 | 83.3 (77.5) | 176.1 (129.4) | 304.9 (194.6) | 93.1 (82.8) | 332.7 (256.8) | 26.2 (24.8) | 17.4 (23.8) |
| CARRS-1 | 219.4 (230.8) | 126.9 (89.7) | 449.0 (202.8) | 136.1 (111.5) | 66.1 (88.7) | 6.9 (9.6) | 49.0 (49.4) |
| CARRS-2 | 201.7 (339.4) | 178.0 (122.0) | 406.4 (224.7) | 137.2 (107.9) | 100.0 (128.5) | 4.1 (24.1) | 21.6 (36.7) |
| EPIC-Oxford | 99.8 (70.4) | 113.8 (77.5) | 272.9 (148.9) | 74.9 (52.2) | 276.4 (221.2) | 9.5 (14.2) | 68.6 (54.2) |
| Oxford Vegetarian Study | 116.8 (66.9) | 29.5 (24.8) | 120.1 (57.3) | 81.7 (44.3) | 85.8 (34.0) | 19.6 (16.3) | 31.8 (25.7) |
| Tzu Chi Health Study | 347.7 (244.9) | 66.4 (74.6) | 519.9 (347.1) | 20.2 (36.0) | 154.9 (147.2) | 5.6 (11.4) | 11.1 (18.9) |
| UK Women's Cohort Study | 93.4 (69.9) | 101.5 (74.8) | 280.9 (141.5) | 93.8 (56.2) | 345.4 (250.4) | 12.0 (15.1) | 65.1 (51.9) |
| *Very large cohorts* |  |  |  |  |  |  |  |
| China Kadoorie Biobank<sup>3</sup> | 242.3 (43.3) | 89.9 (66.7) | 235.6 (10.1) | - | 65.0 (27.0) | - | - |
| Million Women Study | 111.9 (65.6) | 99.3 (64.2) | 134.7 (90.6) | 23.8 (20.8) | 356.5 (220.2) | 11.0 (15.5) | 39.1 (41.1) |
| NIH-AARP | 132.4 (99.3) | 90.1 (101.2) | 471.4 (317.4) | 74.2 (84.4) | 494.2 (389.9) | 10.6 (18.7) | 32.6 (38.2) |
| UK Biobank<sup>3</sup> | 142.2 (16.6) | 90.6 (27.7) | 210.5 (48.6) | - | 207.2 (88.4) | - | - |
| Cohort | Plant foods, g/day |  | Animal foods, g/day |  |  |  |  |
|  | Plant milks | Sugar sweetened beverages | Total dairy products<sup>4</sup> | Dairy milk | Cheese | Yogurt | Eggs & egg dishes |
| *Cohorts with large proportions of vegetarians* |  |  |  |  |  |  |  |
| Adventist Health Study-2 | 115.8 (161.2) | 76.3 (222.6) | 104.6 (178.8) | 66.9 (157.0) | 15.6 (24.2) | 22.1 (49.1) | 7.8 (13.5) |
| CARRS-1 | - | 53.5 (60.5) | 217.3 (135.2) | - | - | - | 4.3 (9.8) |
| CARRS-2 | - | 19.0 (41.9) | 229.0 (190.8) | 143.8 (157.5) | - | 85.2 (90.3) | 3.9 (16.1) |
| EPIC-Oxford | 32.4 (97.8) | 94.7 (168.1) | 376.0 (240.8) | 256.6 (203.7) | 30.1 (25.2) | 33.8 (41.3) | 10.6 (12.1) |
| Oxford Vegetarian Study | - | - | 160.0 (109.6) | 113.8 (103.2) | 46.2 (27.1) | - | 25.7 (21.6) |
| Tzu Chi Health Study | 85.7 (126.9) | 13.5 (76.3) | 42.6 (68.8) | 33.0 (56.5) | 1.1 (3.6) | 8.7 (32.5) | 16.4 (16.7) |
| UK Women's Cohort Study | 24.7 (89.2) | 54.0 (120.6) | 366.4 (227.0) | 253.1 (199.6) | 34.8 (31.1) | 57.1 (69.0) | 14.7 (15.4) |
| *Very large cohorts* |  |  |  |  |  |  |  |
| China Kadoorie Biobank<sup>3</sup> | - | - | 38.9 (32.3) | - | - | - | 33.0 (9.5) |
| Million Women Study | 24.1 (60.2) | 101.2 (216.9) | 345.9 (214.0) | 223.8 (186.3) | 31.1 (21.5) | 81.0 (78.6) | 15.9 (14.8) |
| NIH-AARP | - | 609.6 (977.4) | 302.7 (390.2) | 195.6 (360.6) | 9.1 (13.7) | 33.3 (64.5) | 5.3 (11.3) |
| UK Biobank<sup>3</sup> | - | - | - | - | 18.3 (6.1) | - | - |

Abbreviations: CARRS, Centre for cArdiometabolic Risk Reduction in South Asia; EPIC, European Prospective Investigation into Cancer and Nutrition; NIH-AARP, National Institutes of Health-AARP Diet and Health Study.
<sup>1</sup>All vegetarians include lacto-ovo, lacto and ovo vegetarians.
<sup>2</sup>Values are mean (SD).
<sup>3</sup>Mean intakes assessed at resurvey have been assigned to baseline categories based on quintiles of intakes to estimate the baseline intakes.
<sup>4</sup>Total dairy products include dairy milk, cheese, yogurt and other dairy products if available in the specified cohort.
“-” indicates that no information was available for this food group in the specified cohort.

In vegans, broadly similar differences in food intakes between studies as in the vegetarians were observed (Table S7).

### Food intakes of meat eaters across cohorts

Among all meat eaters, the reported total red and processed meat intake was highest in the Oxford Vegetarian Study [76 (39) g/d] followed by the UK Women’s Cohort Study [71 (45) g/d] and NIH-AARP [69 (51) g/d], while the lowest intake was reported in AHS-2 [24 (27) g/d] (**Figure 1**). Total vegetable intake was the highest in the Tzu Chi Health Study [428 (292) g/d], while the lowest intake was reported in the Oxford Vegetarian Study [92 (49) g/d] (**Figure 2**). For fresh fruit (**Figure 2**), the highest intakes were reported in the UK and US cohorts, except for the Oxford Vegetarian Study, while participants in CARRS-1, CARRS-2, and the China Kadoorie Biobank reported the lowest mean intakes.

**Figure 1.**
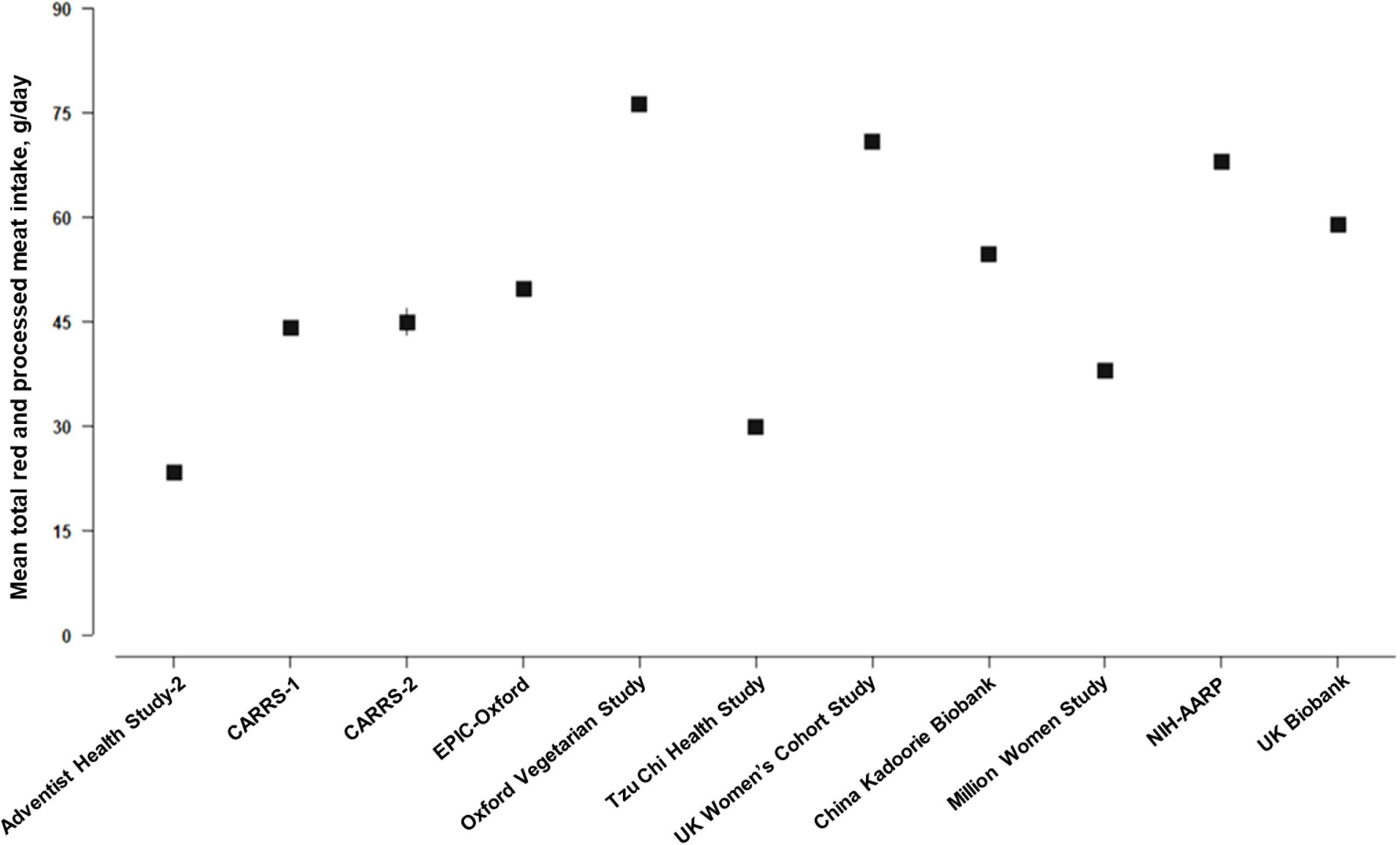
Mean (95% CI) total red and processed meat intake among meat eaters by cohort. Abbreviations: CARRS, Centre for cArdiometabolic Risk Reduction in South Asia; EPIC, European Prospective Investigation into Cancer and Nutrition; NIH-AARP, National Institutes of Health-American Association of Retired Persons Diet and Health Study.

**Figure 2.**
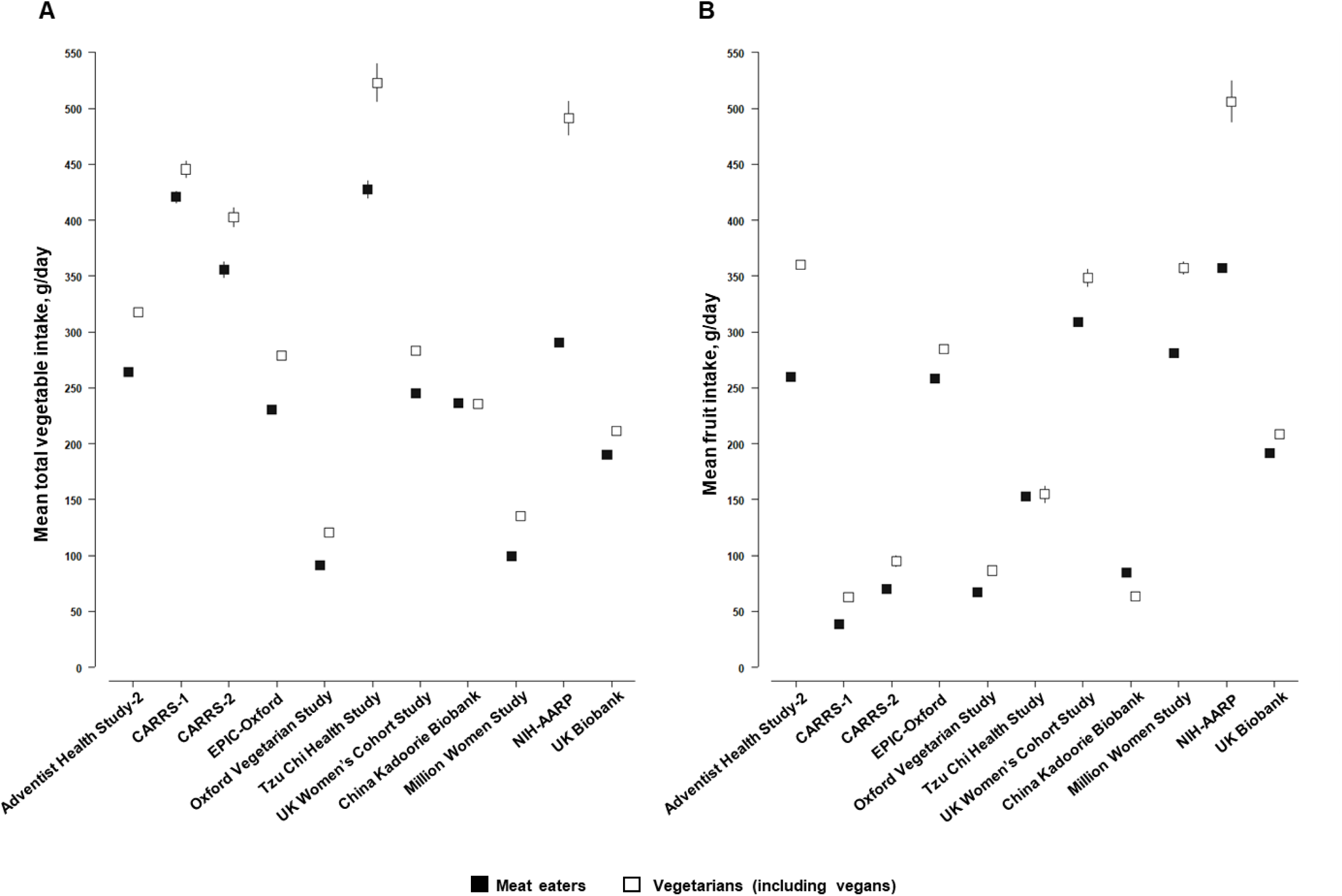
Mean (95% CI) intakes of **(A)** total vegetables and **(B)** fruit in meat eaters versus vegetarians (including vegans) by cohort. Abbreviations: CARRS, Centre for cArdiometabolic Risk Reduction in South Asia; EPIC, European Prospective Investigation into Cancer and Nutrition; NIH-AARP, National Institutes of Health-American Association of Retired Persons Diet and Health Study.

### Estimated nutrient intakes across diet groups and cohorts

In general, the intakes of nutrients such as protein, total fat, saturated fat, monounsaturated fat, vitamin B12, and vitamin D were higher among meat eaters compared to vegetarians and vegans (Table S8). On the other hand, vegans had higher intakes of dietary fibre and vitamin C compared to the other dietary groups, and lower intakes of calcium.

### Diet group consistency at resurvey

In most of the cohorts with resurvey dietary data, ≥90% of baseline meat eaters continued to follow a meat-eating diet at resurvey, with the exceptions being CARRS-1 (72%) and the Oxford Vegetarian Study (87%) (Table S9). In the four UK cohorts with dietary data during the follow-up, 56–83% of the baseline pescatarians remained as pescatarians at resurvey. However, in CARRS-1 and the China Kadoorie Biobank, 46% of people who were pescatarian at baseline were classified as meat eaters at the resurvey. Furthermore, in the UK cohorts and CARRS-1, 68–94% of baseline vegetarians remained as vegetarians at resurvey; however, in the China Kadoorie Biobank, only 19% continued to follow a vegetarian diet, while 69% had incorporated red meat into their diet. Among the baseline vegans in the UK cohorts, most participants either continued to follow a vegan diet or were classified as vegetarian. In contrast, in CARRS-1, the majority of baseline vegans were vegetarians at resurvey (80%), while in the China Kadoorie Biobank the majority of vegans were meat eaters (65%) at resurvey.

### Sex, sociodemographic and lifestyle factors, and body mass index differences by diet group

Overall, a higher proportion of men were meat eaters (93%) compared to women (91%) while a higher proportion of women were vegetarians (4%) compared to men (3%) (Table S4). However, in the Adventist Health Study-2 and EPIC-Oxford, a greater proportion of men compared to women reported that they were vegetarian or vegan. When comparing ethnicity by diet group across the cohorts, we observed that most of the UK and US studies, except for the Adventist Health Study-2, had a higher proportion of Asian participants among lacto vegetarians compared to the other diet groups (Table S10).

The proportion of participants with a university degree or equivalent was generally higher among those who followed any vegetarian diet (**Table 8**). However, in the Tzu Chi Health Study and the China Kadoorie Biobank, the proportion of participants with a university degree or equivalent was higher among the meat eaters. **Table 9** shows the lifestyle factors by diet group and cohort. Except for the UK Women’s Cohort Study and the Million Women Study, a higher proportion of participants who were classified as poultry eaters, pescatarians, vegetarians and vegans were never smokers compared to meat eaters. In general, meat eaters had a higher mean alcohol intake compared to other diet groups, though such differences were not observed in EPIC-Oxford, the UK Women’s Cohort Study and the Million Women Study. In the Adventist Health Study-2, EPIC-Oxford, the Oxford Vegetarian Study, NIH-AARP and UK Biobank, a higher proportion of participants who were poultry eaters, pescatarians, vegetarians or vegans showed a high level of physical activity compared to meat eaters. In CARRS-1, the vegetarians and vegans showed the highest level of physical activity compared to participants in the other diet groups, while in the China Kadoorie Biobank, a smaller proportion of participants who were pescatarians were highly active compared to the other diet groups.

**Table 8:**
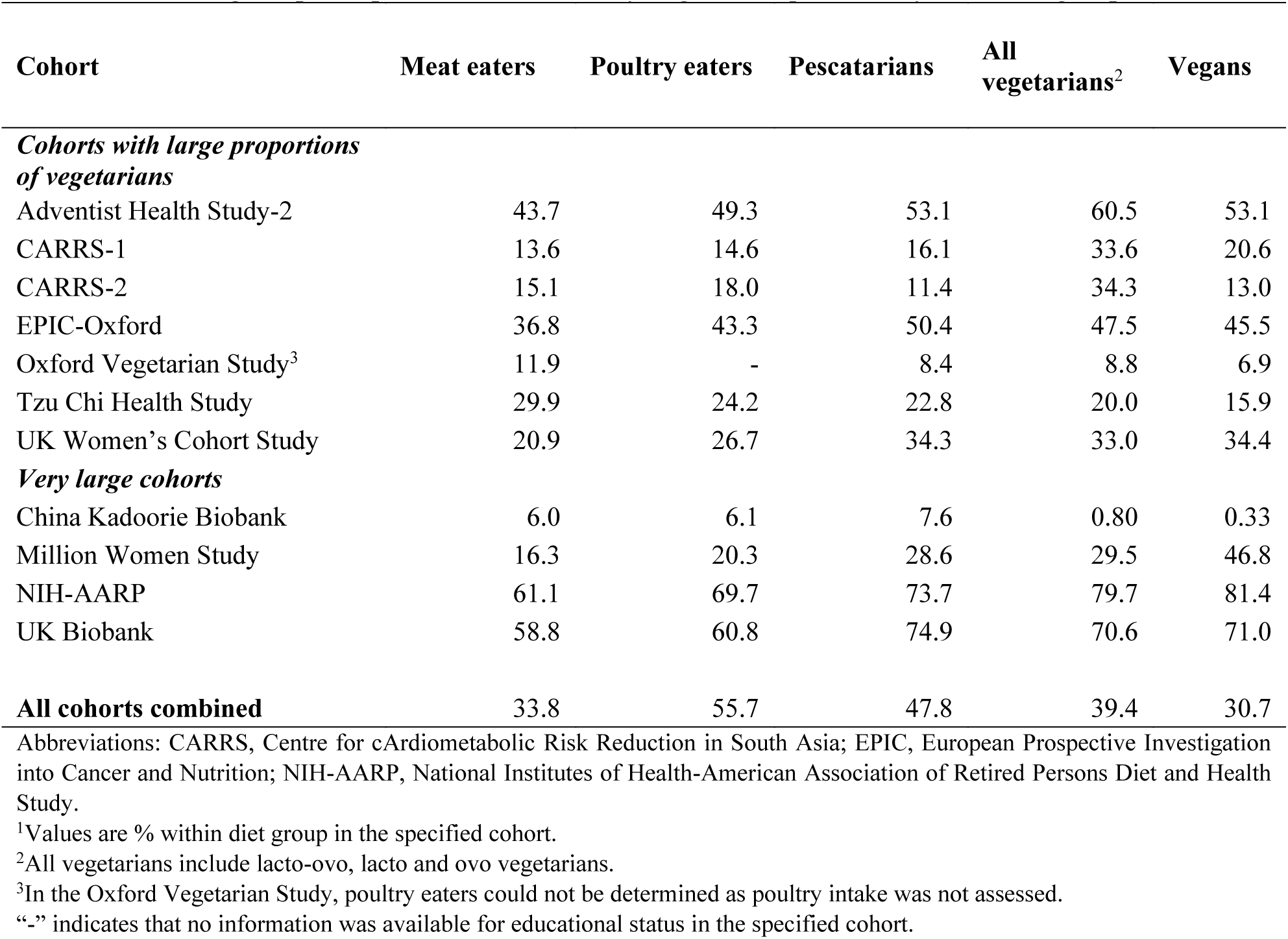
Percentage of participants with a university degree or equivalent by main diet groups and cohort.

| Cohort | Meat eaters | Poultry eaters | Pescatarians | All vegetarians<sup>2</sup> | Vegans |
| --- | --- | --- | --- | --- | --- |
| ***Cohorts with large proportions of vegetarians*** |  |  |  |  |  |
| Adventist Health Study-2 | 43.7 | 49.3 | 53.1 | 60.5 | 53.1 |
| CARRS-1 | 13.6 | 14.6 | 16.1 | 33.6 | 20.6 |
| CARRS-2 | 15.1 | 18.0 | 11.4 | 34.3 | 13.0 |
| EPIC-Oxford | 36.8 | 43.3 | 50.4 | 47.5 | 45.5 |
| Oxford Vegetarian Study<sup>3</sup> | 11.9 | - | 8.4 | 8.8 | 6.9 |
| Tzu Chi Health Study | 29.9 | 24.2 | 22.8 | 20.0 | 15.9 |
| UK Women's Cohort Study | 20.9 | 26.7 | 34.3 | 33.0 | 34.4 |
| ***Very large cohorts*** |  |  |  |  |  |
| China Kadoorie Biobank | 6.0 | 6.1 | 7.6 | 0.80 | 0.33 |
| Million Women Study | 16.3 | 20.3 | 28.6 | 29.5 | 46.8 |
| NIH-AARP | 61.1 | 69.7 | 73.7 | 79.7 | 81.4 |
| UK Biobank | 58.8 | 60.8 | 74.9 | 70.6 | 71.0 |
| **All cohorts combined** | **33.8** | **55.7** | **47.8** | **39.4** | **30.7** |

Abbreviations: CARRS, Centre for cArdiometabolic Risk Reduction in South Asia; EPIC, European Prospective Investigation into Cancer and Nutrition; NIH-AARP, National Institutes of Health-American Association of Retired Persons Diet and Health Study.
<sup>1</sup>Values are % within diet group in the specified cohort.
<sup>2</sup>All vegetarians include lacto-ovo, lacto and ovo vegetarians.
<sup>3</sup>In the Oxford Vegetarian Study, poultry eaters could not be determined as poultry intake was not assessed.
“-” indicates that no information was available for educational status in the specified cohort.

**Table 9.**
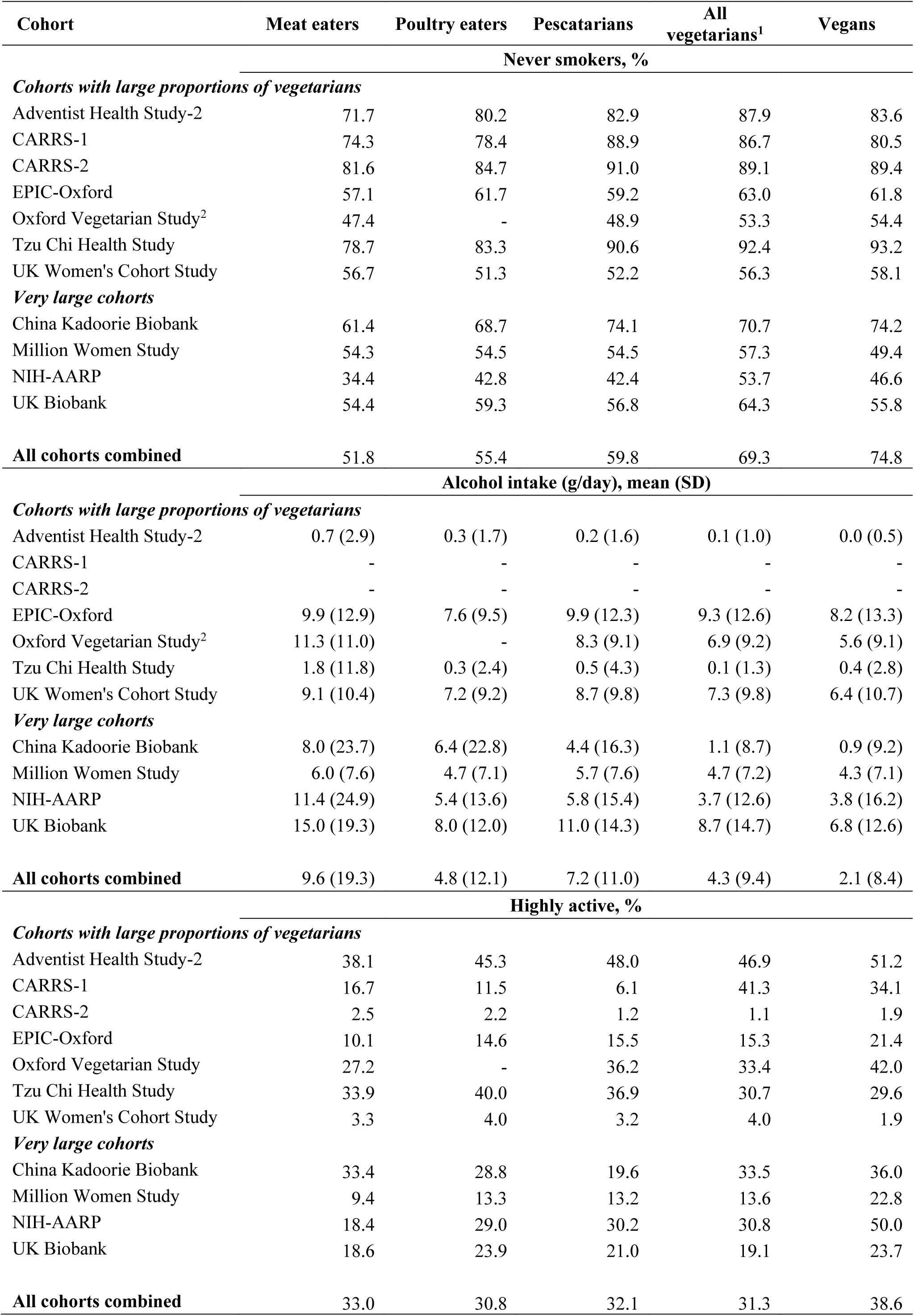
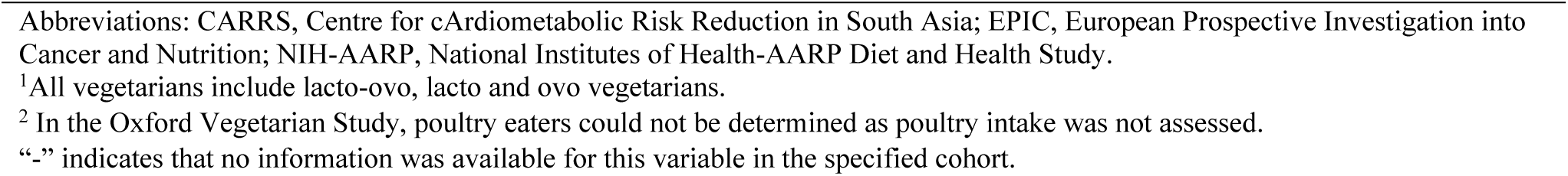
Lifestyle factors by main diet groups and cohort.

| Cohort | Meat eaters | Poultry eaters | Pescatarians | All vegetarians<sup>1</sup> | Vegans |
| --- | --- | --- | --- | --- | --- |
| Never smokers, % |  |  |  |  |  |
| *Cohorts with large proportions of vegetarians* |  |  |  |  |  |
| Adventist Health Study-2 | 71.7 | 80.2 | 82.9 | 87.9 | 83.6 |
| CARRS-1 | 74.3 | 78.4 | 88.9 | 86.7 | 80.5 |
| CARRS-2 | 81.6 | 84.7 | 91.0 | 89.1 | 89.4 |
| EPIC-Oxford | 57.1 | 61.7 | 59.2 | 63.0 | 61.8 |
| Oxford Vegetarian Study<sup>2</sup> | 47.4 | - | 48.9 | 53.3 | 54.4 |
| Tzu Chi Health Study | 78.7 | 83.3 | 90.6 | 92.4 | 93.2 |
| UK Women's Cohort Study | 56.7 | 51.3 | 52.2 | 56.3 | 58.1 |
| *Very large cohorts* |  |  |  |  |  |
| China Kadoorie Biobank | 61.4 | 68.7 | 74.1 | 70.7 | 74.2 |
| Million Women Study | 54.3 | 54.5 | 54.5 | 57.3 | 49.4 |
| NIH-AARP | 34.4 | 42.8 | 42.4 | 53.7 | 46.6 |
| UK Biobank | 54.4 | 59.3 | 56.8 | 64.3 | 55.8 |
| **All cohorts combined** | **51.8** | **55.4** | **59.8** | **69.3** | **74.8** |
| Alcohol intake (g/day), mean (SD) |  |  |  |  |  |
| *Cohorts with large proportions of vegetarians* |  |  |  |  |  |
| Adventist Health Study-2 | 0.7 (2.9) | 0.3 (1.7) | 0.2 (1.6) | 0.1 (1.0) | 0.0 (0.5) |
| CARRS-1 | - | - | - | - | - |
| CARRS-2 | - | - | - | - | - |
| EPIC-Oxford | 9.9 (12.9) | 7.6 (9.5) | 9.9 (12.3) | 9.3 (12.6) | 8.2 (13.3) |
| Oxford Vegetarian Study<sup>2</sup> | 11.3 (11.0) | - | 8.3 (9.1) | 6.9 (9.2) | 5.6 (9.1) |
| Tzu Chi Health Study | 1.8 (11.8) | 0.3 (2.4) | 0.5 (4.3) | 0.1 (1.3) | 0.4 (2.8) |
| UK Women's Cohort Study | 9.1 (10.4) | 7.2 (9.2) | 8.7 (9.8) | 7.3 (9.8) | 6.4 (10.7) |
| *Very large cohorts* |  |  |  |  |  |
| China Kadoorie Biobank | 8.0 (23.7) | 6.4 (22.8) | 4.4 (16.3) | 1.1 (8.7) | 0.9 (9.2) |
| Million Women Study | 6.0 (7.6) | 4.7 (7.1) | 5.7 (7.6) | 4.7 (7.2) | 4.3 (7.1) |
| NIH-AARP | 11.4 (24.9) | 5.4 (13.6) | 5.8 (15.4) | 3.7 (12.6) | 3.8 (16.2) |
| UK Biobank | 15.0 (19.3) | 8.0 (12.0) | 11.0 (14.3) | 8.7 (14.7) | 6.8 (12.6) |
| **All cohorts combined** | **9.6 (19.3)** | **4.8 (12.1)** | **7.2 (11.0)** | **4.3 (9.4)** | **2.1 (8.4)** |
| Highly active, % |  |  |  |  |  |
| *Cohorts with large proportions of vegetarians* |  |  |  |  |  |
| Adventist Health Study-2 | 38.1 | 45.3 | 48.0 | 46.9 | 51.2 |
| CARRS-1 | 16.7 | 11.5 | 6.1 | 41.3 | 34.1 |
| CARRS-2 | 2.5 | 2.2 | 1.2 | 1.1 | 1.9 |
| EPIC-Oxford | 10.1 | 14.6 | 15.5 | 15.3 | 21.4 |
| Oxford Vegetarian Study | 27.2 | - | 36.2 | 33.4 | 42.0 |
| Tzu Chi Health Study | 33.9 | 40.0 | 36.9 | 30.7 | 29.6 |
| UK Women's Cohort Study | 3.3 | 4.0 | 3.2 | 4.0 | 1.9 |
| *Very large cohorts* |  |  |  |  |  |
| China Kadoorie Biobank | 33.4 | 28.8 | 19.6 | 33.5 | 36.0 |
| Million Women Study | 9.4 | 13.3 | 13.2 | 13.6 | 22.8 |
| NIH-AARP | 18.4 | 29.0 | 30.2 | 30.8 | 50.0 |
| UK Biobank | 18.6 | 23.9 | 21.0 | 19.1 | 23.7 |
| **All cohorts combined** | **33.0** | **30.8** | **32.1** | **31.3** | **38.6** |

“-” indicates that no information was available for this variable in the specified cohort.

Overall, participants who were regular meat eaters in the UK and US cohorts and the Tzu Chi Health Study had a higher BMI compared to participants in the other diet groups in the same cohort (**Figure 3**). Regular meat eaters in the Adventist Health Study-2 had the highest mean BMI [31 (7) kg/m^2^] compared to all diet groups in all the studies. Across all the studies, the Indian cohorts had the highest BMI among all vegetarians (**Figure 3**). Moreover, the Oxford Vegetarian Study had the participants with the lowest BMI when comparing all the diet groups. In the China Kadoorie Biobank, the mean BMIs of participants across the diet groups were relatively similar, with pescatarians having the highest BMI (**Figure 3**).

**Figure 3.**
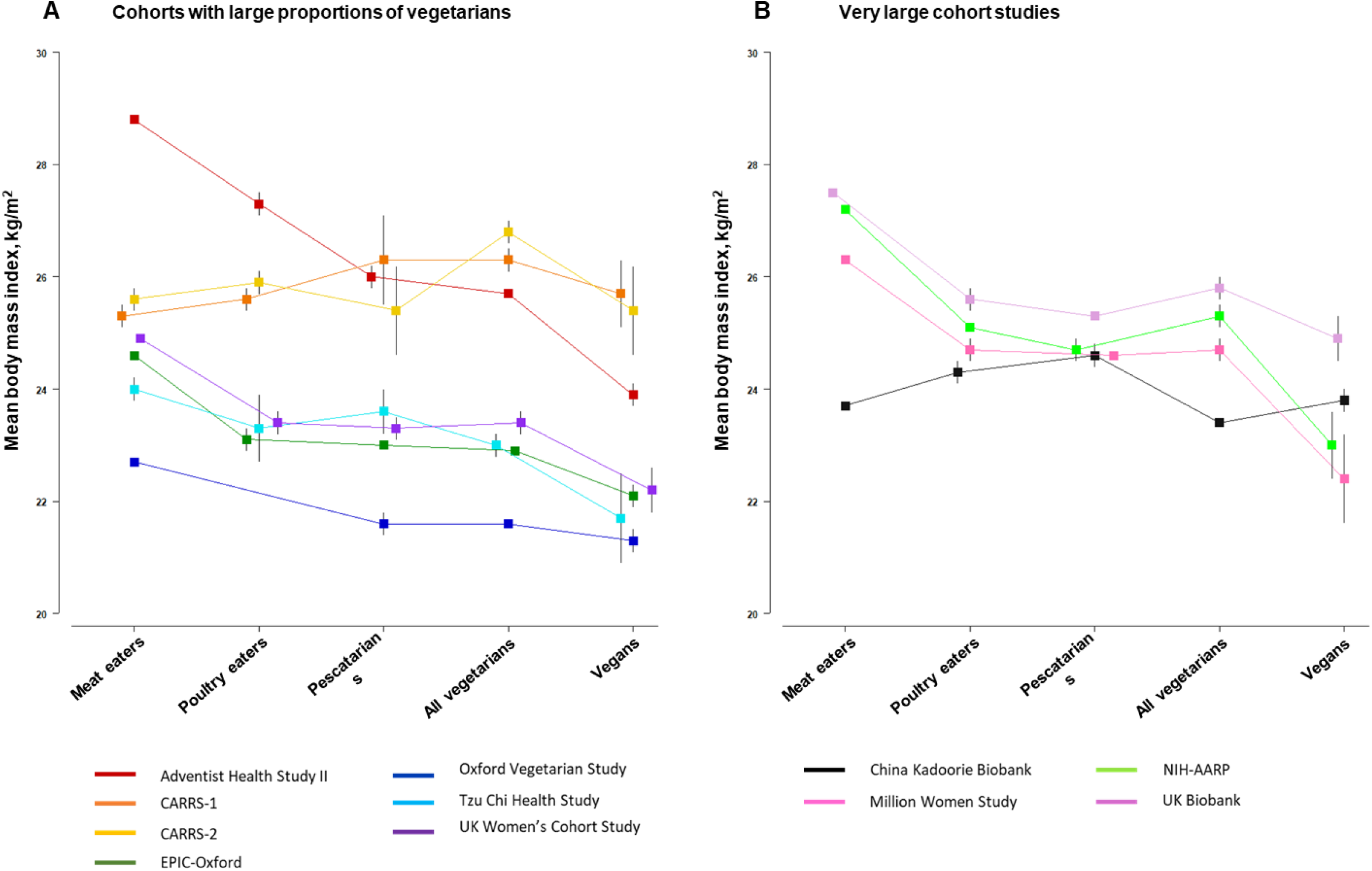
Mean (95% CI) body mass index by diet group in **(A)** cohorts with large proportions of vegetarians and **(B)** very large cohorts. Abbreviations: CARRS, Centre for cArdiometabolic Risk Reduction in South Asia; EPIC, European Prospective Investigation into Cancer and Nutrition; NIH-AARP, National Institutes of Health-AARP Diet and Health Study. All vegetarians include lacto-ovo, lacto and ovo vegetarians. In the Oxford Vegetarian Study, poultry eaters could not be determined as poultry intake was not assessed.

## 4. Discussion

This unique international consortium of prospective cohort studies with large numbers of vegetarians was established to assess the associations between various diet groups and the risks of site-specific cancers, with a particular focus on vegetarian and vegan diets. Our results presented here on the cohort characteristics showed differences between cohorts in food intakes, BMI, and educational status within each diet group. These findings support the interpretation that while vegetarians worldwide share the avoidance of meat, poultry, and fish, their diets, socio-demographics, and lifestyle factors vary substantially.

### Dietary groups and food intakes across the different cohorts

The Adventist Health Study-2, EPIC-Oxford, and the China Kadoorie Biobank contributed the largest numbers of vegetarians and vegans; however, the dietary groups in the China Kadoorie Biobank require careful interpretation (see further discussion below). Among vegetarians, the lacto-ovo vegetarian diet was most prevalent in the UK and US cohorts as well as in the Tzu Chi Health Study, while vegetarians in the two Indian studies mostly followed a lacto vegetarian diet. In recent years, vegetarian and vegan diets have become more popular in Western countries, and this is likely motivated by self-choice due to ethical, environmental and health concerns (28, 29). In India, where vegetarianism has been a cultural tradition for centuries, people tend to follow this diet from birth due to religious and cultural beliefs that encourage the avoidance of meat consumption (30); recent statistics show that 29% of women and 17% of men in India are vegetarian (31). In the US, the estimated proportion of the population following a vegetarian diet was around 3% in 2016 (32, 33), while in the UK, as of 2023, it is estimated that around 5% of the population is vegetarian (34).

We found substantial differences in reported food intakes across cohorts, which may be due to both actual dietary variations and to variations in dietary assessment methods. For example, we observed that intakes of wholegrains, total vegetables, and fresh fruit were markedly lower in vegetarians in the Oxford Vegetarian Study compared to vegetarians in EPIC-Oxford, the UK Women’s Cohort Study and the Million Women Study. This might in part be due to the limited 28 food items-FFQ in the Oxford Vegetarian Study, with only two questions related to fruit consumption, while the three other UK cohorts had more comprehensive FFQs covering over 100 food items, including at least 11 questions on fruit intake. Higher numbers of the food items used to generate food groups could tend to result in higher mean intakes, which may contribute to some of the observed differences between cohorts. However, the observed differences in reported food intakes may also be influenced by time trends in food consumption and/or the assumptions made within different cohorts when allocating portion sizes. Another example of variations in food intakes due to differences in the dietary questionnaires can be observed in the two Indian cohorts. These cohorts recruited participants from the same demographic regions, yet differences in mean food intakes were noted, which could be attributed to the use of a non-quantitative FFQ at baseline in CARRS-1, while a semi-quantitative FFQ was used in CARRS-2.

In the China Kadoorie Biobank, the substantial number of participants classified as vegetarians and vegans might not be attributed to personal choices, such as health or religious beliefs. Instead, this may be related to socioeconomic status, poverty, and affordability (35). For instance, in this cohort, the meat eaters, poultry eaters, and pescatarians were more likely to have a university degree or equivalent (and higher income; findings not shown) compared to the vegetarians and vegans. We also found that vegetarians and vegans in this cohort were mostly from two rural regions of China (Gansu and Henan), while pescatarians were mainly from two coastal regions (Qingdao and Harbin; findings not shown).

### Differences in sex, sociodemographic, and lifestyle factors by diet groups in the different cohorts

Comparisons across the individual cohorts showed that pescatarians, vegetarians, and vegans in studies from the UK, US, and India were more likely to have a university degree or equivalent than the meat eaters within the same cohort. These findings are in alignment with cross-sectional analyses in a random sample of Germans (36) and Finnish adults (37), and in the French NutriNet-Santé study (38).

Differences in sex distribution across the diet groups within each individual study were observed. In most of the cohorts, there was a higher proportion of women in the vegetarian diet group while a higher proportion of men were meat eaters. These findings are in line with previous research in France and Germany (36, 38), which also showed that women typically consume less meat and are more likely to follow a vegetarian or vegan diet.

### BMI differences across diet groups and studies

In the UK and US cohorts, meat eaters had a higher average BMI, while vegans followed by vegetarians had the lowest. The consumption of red and processed meat may be associated with having a greater body weight; a meta-analysis of 18 studies, mainly from Western countries, showed a positive association with obesity (39). The EPIC-Europe study, which included 373,803 men and women, found that higher intakes of total meat, red meat, processed meat, and poultry were associated with greater weight gain over 5 years of follow-up (40). Dietary differences between meat eaters and vegetarians/vegans, such as higher intakes of fibre and lower intakes of protein, may contribute to the latter group’s lower BMI (41, 42).

When comparing the BMI of vegetarians and vegans across the different studies, those in CARRS-1 and CARRS-2 had the highest mean BMI. In these Indian studies, lacto vegetarians had a higher average BMI compared to participants in the other diet groups. These findings are consistent with the cross-sectional National Family Health Survey (2005–06) data in India, which also reported the highest mean BMI in the lacto vegetarians (43). Cross-sectional analyses conducted in CARRS-1 and in adults from the general US population (i.e. including all ethnicities) who completed the National Health and Nutrition Examination Survey (NHANES) showed differences in vegetarian diets between these two countries (32); the diets of vegetarians versus meat eaters were healthier in the NHANES compared to the vegetarians in CARRS-1, where the diets of meat eaters were more similar to those of vegetarians. Differences in cooking practices, meal preparation methods, eating frequencies, and eating out habits between vegetarians in India and those from other countries are also important factors that can be associated with variations in BMI (44). However, the evaluation of these dietary and other lifestyle factors such as physical activity was outside the scope of this consortium.

### Diet groups at resurvey

The agreement of diet groups at baseline and resurvey for meat eaters and vegetarians was generally high (more than ∼70%) in the UK cohorts and CARRS-1. This is similar to previous findings in the Adventist Health Study-2, which found greatest stability among non-vegetarians and lacto-ovo vegetarians when lifetime dietary patterns were assessed using a reliable life-course dietary questionnaire (45). Changes in individuals’ diet groups during follow up were mostly to the adjacent category(ies) of animal product consumption (e.g. poultry eaters to either meat eaters or pescatarians), which may be due to misclassification or true dietary changes over time.

Unlike the patterns observed in the other cohorts, in the China Kadoorie Biobank the great majority of baseline vegetarians and vegans reported consuming meat at follow-up (69% and 65%, respectively). This shift coincided with an improvement in socioeconomic status among these participants, as indicated by their reported income in the follow-up questionnaire (findings not shown). Moreover, there was a substantial increase in meat consumption in China during this period, unlike in most high-income countries where meat consumption has plateaued or decreased (46, 47). This further supports the interpretation that vegetarians and vegans in the China Kadoorie Biobank were more likely to report very low or no meat consumption at baseline due to economic constraints, rather than intentionally adhering to a vegetarian diet. Moreover, as previously discussed, the participants in this cohort classified as vegetarians at baseline may have occasionally consumed both meat and fish because they reported eating these foods less than monthly due to the design of the FFQ. These findings show the importance of carefully considering the stability of diet groups over time when conducting analyses of the associations of diet group with long-term health outcomes.

### Strengths and limitations

These analyses have several strengths. This consortium will provide the largest collection of observational data on vegetarian and vegan diets and the risk of individual cancer sites available. The consortium includes participants from the UK, US, and Asia (India, China, and Taiwan). While CARRS-1, CARRS-2 and the Tzu Chi Health Study do not include large numbers of participants, they offer valuable insights for this research on vegetarian and vegan diets because food intakes in these areas differ markedly from those in studies conducted in the UK and the US. We have also compared key participant characteristics, such as sex, BMI, and educational status, across diet groups within the individual cohorts. Further strengths of this consortium are the availability and standardisation of various potential confounders from the different cohorts, which can be used in prospective analyses, offering a unique advantage when investigating the associations between diet groups and the risk of site-specific cancers, which is not possible in meta-analyses of published data.

It should be noted that the differences reported in this paper are descriptions specific to the participants in the consortium, and therefore some of the differences are likely to be influenced by variations between the studies in factors such as age, gender proportions, body size, social and cultural factors, year of recruitment, recruitment age, and methods of dietary ascertainment. Another potential limitation of this consortium is the possibility of diet group misclassification. In the Adventist Health Study-2, CARRS-1, CARRS-2, Tzu Chi Health Study, and China Kadoorie Biobank, the lowest category in the respective FFQ was either “never/rarely” or “never/less than once a month”. Therefore, participants classified as vegetarians or vegans may occasionally consume the relevant animal foods. Misclassification of the diet groups is also possible depending on food items included or omitted, social desirability, and other factors that may vary between the cohorts.

In this consortium, which harmonised data from 11 prospective cohort studies, food intakes among vegetarians varied between studies. This variation may be attributed to differences in culture, socioeconomic status, and the specific dietary assessment tools used in each cohort. Vegetarians and vegans were found to have a lower BMI and higher educational status in comparison to regular meat eaters, showing a gradient across the other diet groups; however, this pattern was not observed in some of the Asian cohorts. In general, vegetarians had lower alcohol intake, a higher proportion of individuals who had never smoked, and higher physical activity when compared to meat eaters. The data harmonised will be used in future analyses to assess the prospective associations between vegetarian diets and risks of site-specific cancer.

## Data Availability

Data described in the manuscript will not be made available because studies pooled by the Cancer Risk in Vegetarians Consortium are not owned by the writing group and so are not available from this consortium. Individual studies may be contacted to request access to their data.

## Author contributions

Statement of authors’ contributions to manuscript: APC and TJK designed the research; YD, JYL and APC conducted the research; YD, JYL, CZW and APC analysed data; YD wrote the paper; all authors commented on the analysis, made important contributions to interpretation of the findings, reviewed the paper critically for important intellectual content, read and approved the final manuscript.

## References

1. Bray F, Laversanne M, Weiderpass E, Soerjomataram I. The ever-increasing importance of cancer as a leading cause of premature death worldwide. Cancer-Am Cancer Soc. 2021;127(16):3029–30.

2. World Health Organization. Preventing cancer. Available from: https://www.who.int/activities/preventing-cancer

3. World Cancer Research Fund/American Institute for Cancer Research. Diet, Nutrition, Physical Activity and Cancer: a Global Perspective. 2018. Available from: https://www.wcrf.org/diet-activity-and-cancer/global-cancer-update-programme/resources-and-toolkits/

4. Key TJ, Bradbury KE, Perez-Cornago A, Sinha R, Tsilidis KK, Tsugane S. Diet, nutrition, and cancer risk: what do we know and what is the way forward? BMJ. 2020;368:m511.

5. Tantamango-Bartley Y, Jaceldo-Siegl K, Fan J, Fraser G. Vegetarian diets and the incidence of cancer in a low-risk population. Cancer Epidemiol Biomarkers Prev. 2013;22(2):286–94.

6. Key TJ, Appleby PN, Crowe FL, Bradbury KE, Schmidt JA, Travis RC. Cancer in British vegetarians: updated analyses of 4998 incident cancers in a cohort of 32,491 meat eaters, 8612 fish eaters, 18,298 vegetarians, and 2246 vegans. Am J Clin Nutr. 2014;100 Suppl 1(1):378S–85S.

7. Thorogood M, Mann J, Appleby P, McPherson K. Risk of death from cancer and ischaemic heart disease in meat and non-meat eaters. BMJ. 1994;308(6945):1667–70.

8. Cade JE, Taylor EF, Burley VJ, Greenwood DC. Common dietary patterns and risk of breast cancer: analysis from the United Kingdom Women’s Cohort Study. Nutr Cancer. 2010;62(3):300–6.

9. Gilsing AM, Schouten LJ, Goldbohm RA, Dagnelie PC, van den Brandt PA, Weijenberg MP. Vegetarianism, low meat consumption and the risk of colorectal cancer in a population based cohort study. Sci Rep. 2015;5:13484.

10. Watling CZ, Schmidt JA, Dunneram Y, Tong TYN, Kelly RK, Knuppel A, et al. Risk of cancer in regular and low meat-eaters, fish-eaters, and vegetarians: a prospective analysis of UK Biobank participants. BMC Medicine. 2022;20(1):73.

11. Butler TL, Fraser GE, Beeson WL, Knutsen SF, Herring RP, Chan J, et al. Cohort profile: the Adventist Health Study-2 (AHS-2). Int J Epidemiol. 2008;37(2):260–5.

12. Nair M, Ali MK, Ajay VS, Shivashankar R, Mohan V, Pradeepa R, et al. CARRS Surveillance study: design and methods to assess burdens from multiple perspectives. BMC Public Health. 2012;12.

13. Kondal D, Patel SA, Ali MK, Mohan D, Rautela G, Gujral UP, et al. Cohort profile: the Center for cArdiometabolic Risk Reduction in South Asia (CARRS). Int J Epidemiol. 2022;51(6):e358–e71.

14. Davey GK, Spencer EA, Appleby PN, Allen NE, Knox KH, Key TJ. EPIC-Oxford: lifestyle characteristics and nutrient intakes in a cohort of 33 883 meat-eaters and 31 546 non meat-eaters in the UK. Public Health Nutr. 2003;6(3):259–69.

15. Appleby PN, Thorogood M, Mann JI, Key TJ. The Oxford Vegetarian Study: an overview. Am J Clin Nutr. 1999;70(3 Suppl):525s–31s.

16. Chiu THT, Huang HY, Chen KJ, Wu YR, Chiu JPC, Li YH, et al. Relative validity and reproducibility of a quantitative FFQ for assessing nutrient intakes of vegetarians in Taiwan. Public Health Nutr. 2014;17(7):1459–66.

17. Cade JE, Burley VJ, Alwan NA, Hutchinson J, Hancock N, Morris MA, et al. Cohort profile: the UK Women’s Cohort Study (UKWCS). Int J Epidemiol. 2017;46(2):e11.

18. Chen Z, Chen J, Collins R, Guo Y, Peto R, Wu F, et al. China Kadoorie Biobank of 0.5 million people: survey methods, baseline characteristics and long-term follow-up. Int J Epidemiol. 2011;40(6):1652–66.

19. Green J, Reeves GK, Floud S, Barnes I, Cairns BJ, Gathani T, et al. Cohort profile: the Million Women Study. Int J Epidemiol. 2019;48(1):28–9e.

20. Schatzkin A, Subar AF, Thompson FE, Harlan LC, Tangrea J, Hollenbeck AR, et al. Design and serendipity in establishing a large cohort with wide dietary intake distributions : the National Institutes of Health–American Association of Retired Persons Diet and Health Study. Am J Epidemiol. 2001;154(12):1119–25.

21. Sudlow C, Gallacher J, Allen N, Beral V, Burton P, Danesh J, et al. UK Biobank: an open access resource for identifying the causes of a wide range of complex diseases of middle and old age. PLOS Med. 2015;12(3):e1001779.

22. Jaceldo-Siegl K, Fan J, Sabaté J, Knutsen SF, Haddad E, Beeson WL, et al. Race-specific validation of food intake obtained from a comprehensive FFQ: the Adventist Health Study-2. Public Health Nutr. 2011;14(11):1988–97.

23. Bingham SA, Gill C, Welch A, Cassidy A, Runswick SA, Oakes S, et al. Validation of dietary assessment methods in the UK arm of EPIC using weighed records, and 24-hour urinary nitrogen and potassium and serum vitamin C and carotenoids as biomarkers. Int J Epidemiol. 1997;26 Suppl 1:S137–51.

24. Spence M, Cade J, Burley V, Greenwood D. Ability of the UK Women’s Cohort Study Food Frequency Questionnaire to rank dietary intakes: a preliminary validation study. Proc Nutr Soc. 2002;61:117A.

25. Qin CX, Guo Y, Pei P, Du HD, Yang L, Chen YP, et al. The relative validity and reproducibility of food frequency questionnaires in the China Kadoorie Biobank study. Nutrients. 2022;14(4).

26. Thompson FE, Kipnis V, Midthune D, Freedman LS, Carroll RJ, Subar AF, et al. Performance of a food-frequency questionnaire in the US NIH-AARP (National Institutes of Health-American Association of Retired Persons) Diet and Health Study. Public Health Nutr. 2008;11(2):183–95.

27. Willett W. Nutritional Epidemiology: Oxford University Press; 2012. Available from: https://academic.oup.com/book/27443

28. Sexton AE, Garnett T, Lorimer J. Vegan food geographies and the rise of Big Veganism. Prog Hum Geogr. 2022;46(2):605–28.

29. Cramer H, Kessler CS, Sundberg T, Leach MJ, Schumann D, Adams J, Lauche R. Characteristics of Americans choosing vegetarian and vegan diets for health reasons. J Nutr Educ Behav. 2017;49(7):561–7.e1.

30. Ruby MB. Vegetarianism. A blossoming field of study. Appetite. 2012;58(1):141–50.

31. International Institute for Population Sciences (IIPS) and ICF. National Family Health Survey (NFHS-5), 2019-21: India: Volume I.2021. Available from: https://dhsprogram.com/pubs/pdf/FR375/FR375.pdf

32. Jaacks LM, Kapoor D, Singh K, Narayan KMV, Ali MK, Kadir MM, et al. Vegetarianism and cardiometabolic disease risk factors: differences between South Asian and US adults. Nutrition. 2016;32(9):975–84.

33. Stahler C. How often do Americans eat vegetarian meals? And how many adults in the US are vegetarian. The Vegetarian Resource Group. 2015;29. Available from: https://www.vrg.org/blog/2015/05/29/how-often-do-americans-eat-vegetarian-meals-and-how-many-adults-in-the-u-s-are-vegetarian-2/

34. YouGov. Dietary choices of Brits (e.g. vegeterian, flexitarian, meat-eater etc)? 2022 [Internet]. Available from: https://yougov.co.uk/topics/society/trackers/dietery-choices-of-brits-eg-vegeterian-flexitarian-meat-eater-etc/.

35. Papier K, Kakkoura MG, Guo Y, Knuppel A, Pei P, Tong TYN, et al. Intakes of major food groups in China and UK: results from 100,000 adults in the China Kadoorie Biobank and UK biobank. Eur J Nutr. 2023;62(2):819–32.

36. Paslakis G, Richardson C, Nohre M, Brahler E, Holzapfel C, Hilbert A, de Zwaan M. Prevalence and psychopathology of vegetarians and vegans - results from a representative survey in Germany (vol 10, 6840, 2020). Sci Rep. 2020;10(1).

37. Lehto E, Kaartinen NE, Saaksjarvi K, Mannisto S, Jallinoja P. Vegetarians and different types of meat eaters among the Finnish adult population from 2007 to 2017. Br J Nutr. 2022;127(7):1060–72.

38. Allès B, Baudry J, Méjean C, Touvier M, Péneau S, Hercberg S, Kesse-Guyot E. Comparison of sociodemographic and nutritional characteristics between self-reported vegetarians, vegans, and meat-eaters from the NutriNet-Santé Study. Nutrients. 2017;9(9).

39. Rouhani MH, Salehi-Abargouei A, Surkan PJ, Azadbakht L. Is there a relationship between red or processed meat intake and obesity? A systematic review and meta-analysis of observational studies. Obes Rev. 2014;15(9):740–8.

40. Vergnaud A-C, Norat T, Romaguera D, Mouw T, May AM, Travier N, et al. Meat consumption and prospective weight change in participants of the EPIC-PANACEA study. Am J Clin Nutr. 2010;92(2):398–407.

41. Newby P, Tucker KL, Wolk A. Risk of overweight and obesity among semivegetarian, lactovegetarian, and vegan women. Am J Clin Nutr. 2005;81(6):1267–74.

42. Spencer EA, Appleby PN, Davey GK, Key TJ. Diet and body mass index in 38,000 EPIC-Oxford meat-eaters, fish-eaters, vegetarians and vegans. Int J Obes. 2003;27(6):728–34.

43. Agrawal S, Millett CJ, Dhillon PK, Subramanian SV, Ebrahim S. Type of vegetarian diet, obesity and diabetes in adult Indian population. Nutr J. 2014;13:89.

44. Borude S. Which is a good diet-veg or non-veg? Faith-based vegetarianism for protection from obesity-a myth or actuality? Obes Surg. 2019;29(4):1276–80.

45. Martins MCT, Jaceldo-Siegl K, Orlich M, Fan J, Mashchak A, Fraser GE. A new approach to assess lifetime dietary patterns finds lower consumption of animal foods with aging in a longitudinal analysis of a health-oriented Adventist population. Nutrients. 2017;9(10).

46. Godfray HCJ, Aveyard P, Garnett T, Hall JW, Key TJ, Lorimer J, et al. Meat consumption, health, and the environment. Science. 2018;361(6399).

47. Organisation for Economic Cooperation and Development/ Food Agricultural Organisation (OECD/FAO). OECD-FAO Agricultural Outlook 2023-2032, 2023. Available from: https://www.oecd-ilibrary.org/agriculture-and-food/oecd-fao-agricultural-outlook-2023-2032_08801ab7-en.

