## Supplementary Tables and Figures for "Methods and participant characteristics in the Cancer Risk in Vegetarians Consortium: A cross-sectional analysis across 11 prospective studies"

**Table S1**. Variables in the Cancer Risk in Vegetarians Consortium and the corresponding questionnaire information from the individual cohorts.

| **Variable categories** | **Cohorts with large proportions of vegetarians** | | | | | | | **Very large cohorts** | | | |
|---|---|---|---|---|---|---|---|---|---|---|---|
| | **AHS-2** | **CARRS-1** | **CARRS-2** | **EPIC-Oxford** | **OVS** | **TCHS** | **UKWCS** | **CKB** | **MWS** | **NIH-AARP** | **UK Biobank** |
| | **Marital status (living with a partner)** | | | | | | | | | | |
| | **What is your marital status?** | **What is your marital status?** | **What is your marital status?** | **What is your marital status?** | **What is your spouse's occupation? (if applicable)** | **What is your marital status?** | **What is your marital status?** | **What is your current marital status?** | **Are you currently married or living with a partner?** | **What is your current marital status?** | **How are people in household related to participant?** |
| Yes | • First marriage<br>• Remarried<br>• Common law marriage | • Married | • Married | • Married or living as married | • Married | • Married<br>• Remarried | • Married or living as married | • Married | • Yes | • Married or living as married | • Husband, wife or partner |
| No | • Never married<br>• Separated<br>• Divorced<br>• Widowed | •- Single<br>• Widow/ Widower<br>• Separated/ Divorced | • Single<br>• Widow/ Widower<br>• Separated/ Divorced | • Widowed<br>• Separated<br>• Divorced<br>• Single | • Unmarried | • Unmarried<br>• Separated<br>• Divorced<br>• Widowed | • Divorced<br>• Widowed<br>• Single<br>• Separated | • Widowed<br>• Separated/ divorced<br>• Never married | • No | • Widowed<br>• Divorced<br>• Separated<br>• Never married | • Husband, wife or partner<br>• Son and/or daughter (including step-children)<br>• Brother and/or sister<br>• Mother and/or father<br>• Grandparent<br>• Grandchild |
| Not known | • Missing | • Missing | • Missing | • Missing | • Missing | • Missing | • Missing | • Missing | • Missing | • Missing | • Missing/ prefer not to answer |
| | **Educational status** | | | | | | | | | | |
| | **What is the highest grade of school or year of college that you completed?** | **Educational status (highest attained degree)** | **Educational status (highest attained degree)** | **Do you have any of the following qualifications?** | **What is your occupation? (used as a proxy for educational status)** | **What is your educational level?** | **Do you have any of the following qualifications ?** | **What is the highest level of school education you ever received?** | **What qualification(s ) do you have from school, college or the equivalent?** | **What is the highest grade or level of schooling you completed?** | **Which of the following qualifications do you have?** |
| Less than secondary/high school or equivalent | • Grade school<br>• Some high school | • Illiterate<br>• Literate, no formal education<br>• Primary school | • Illiterate<br>• Literate, no formal education<br>• Primary school | • No qualification | | • Illiterate<br>• Elementary school | • No qualification | • No formal school<br>• Primary school | Derived from age left school, compulsory school leaving age and qualifications | • Less than 8 years<br>• 8-11 years | • No qualifications |

| | | | | | | | | | | | |
|---|---|---|---|---|---|---|---|---|---|---|---|
| Secondary/high school or equivalent | • High school diploma<br>• Trade school diploma<br>• Some college | • Secondary school<br>• High school | Secondary school<br>High school | • CSE<br>• GCE "O" level<br>• "A" level, Highers | | • Senior high school<br>• Junior college | • CSE<br>• GCE "O" level<br>• "A" level, Highers | • Middle school<br>• High school | | • 12 years or completed high school | • CSEs or equivalent<br>• O levels/ GCSEs or equivalent<br>• A levels/ AS levels or equivalent |
| University degree or equivalent | • Associate degree<br>• Bachelor's degree<br>• Master's degree<br>• Doctoral degree | • Professional degree/post graduate<br>• Graduate | • Professional degree/post graduate<br>• Graduate | • Teaching, diploma<br>• Degree | | • University<br>• Master or above | • Teaching, diploma<br>• City & Guilds<br>• Degree | • Technical school/college<br>• University | | • Post-high school or some college<br>• College and post graduate | • College or University Degree<br>• NVQ or HND or HNC or equivalent<br>• Other professional qualifications |
| Not known | • Missing | • Missing | • Missing | • Missing | | • Missing | • Missing | • Missing | • Missing | • Missing | • Missing/ prefer not to answer/none of the above |
| | **Ethnic group** | | | | | | | | | | |
| | **In the table below, select the group(s) that best describes the racial origin or ethnicity of yourself:** | | | **To which of these groups do you consider you belong?** | | | **Which of these groups would consider you belong to?** | | **To which ethnic group do you consider you belong?** | **Which of these best describes your race?** | **What is your ethnic group?** |
| White | • White non-Hispanic | | | • White | • White | | • White | | • White | • Non-Hispanic White | • White |
| Black | • African American<br>• West Indian/Caribbe an<br>• African | | | • Black - Caribbean<br>• Black other | | | • Black - Caribbean<br>• Black other | | • Black | • Non-Hispanic Black | •Black or Black British<br>• Mixed |
| Hispanic | • Latino/ Mexican<br>• Puerto Rican<br>• Cuban<br>• Central American<br>• South American<br>• Other Hispanic | | | | | | | | | • Hispanic | |

| | | | | | | | | | | | |
|---|---|---|---|---|---|---|---|---|---|---|---|
| Asian | • Asian Indian<br>• Chinese<br>• Filipino<br>• Japanese<br>• Korean<br>• Vietnamese<br>• Other Asian | | | • Indian<br>• Pakistani<br>• Bangladeshi<br>• Chinese | • Bangladeshi<br>• Indian<br>• Chinese<br>• Pakistan<br>• Black Caribbean | | • Indian<br>• Pakistani<br>• Bangladeshi<br>• Chinese | | • Asian | • Asian, Pacific Islander, or American Indian/Alaskan Native | • Asian or Asian British<br>• Chinese |
| Other | • Native Hawaiian/other pacific Islander<br>• American Indian | | | • Other | • Black-other<br>• Other | | • Other | | • Other | • American Indian/Alaskan Native | • Mixed<br>• Other ethnic group |
| Not known | • Missing | | | • Missing | • Missing | | • Missing | | • Missing | • Missing | • Missing/do not know/prefer not to answer |
| | **Cigarette smoking** | | | | | | | | | | |
| | **Have you ever smoked regularly?/ If you have ever smoked cigarettes regularly, please mark the number you smoke(d) per day** | **Derived from multiple questions** | **Derived from multiple questions** | **Derived from multiple questions** | **Derived from multiple questions** | **What is your smoking habit?** | **Which one of the following best describes you?** | **Have you smoked any tobacco today?/ How often do you smoke tobacco now?** | **Have you ever been a smoker?/ Are you are smoker now?** | **Derived from multiple questions** | **Do you smoke now? In the past, how often have you smoked tobacco?** |
| Never | • No | | | | | • No cigarette smoking habit | • Never smoked | • Never smoker | • Never | • Never smoked | • Never |
| Previous | • Yes, none in past 12 months<br>• 2-5 years ago<br>• 6-10 years ago<br>• 11-15 years ago<br>• 16+ years ago | | | | | • Used to smoke but have already quit | • Used to smoke every day, but do not smoke at all now | • Ex regular smoker | • Past | • Former smoker | • Previous |
| Current | • Yes, cigars<br>• Yes, pipe<br>• Yes, cigarettes | | | | | • Have a smoking habit | • I smoke every day<br>• I smoke occasionally, but not every day | • Occasional smoker<br>• Smoker | • Current | • Current smoker | • Current |
| Not known | • Missing | | | | | • Missing | • Missing | • Missing | • Missing | • Missing | • Missing/prefer not to answer |

| | | | | | | | | | | | |
|---|---|---|---|---|---|---|---|---|---|---|---|
| | **History of diabetes** | | | | | | | | | | |
| | **If you have never been told by a doctor that you had any of the conditions on this page, only fill the last circle for question 9, and go to question 10 below. Otherwise, fill circles for any condition diagnosed by a physician** | **Have you ever been told by a doctor that you have any of the following diseases?** | **Have you ever been told by a doctor that you have any of the following diseases?** | **Have you ever been told by a doctor that you have, or had, any of the following conditions?** | **Have you ever had any of the following complaints?** | **Have you ever had any of the following conditions?** | **Have you ever been told by a doctor that you have, or had, any of the following conditions?** | **Has a doctor ever told that you had had the following disease?** | **In the last 5 years has a doctor told you that you have had any of the following conditions?** | **Have you ever been told by a doctor that you had any of the following conditions?** | **Has a doctor ever told you that you have diabetes?** |
| Yes | • <5 years ago<br>• 5-9 years ago<br>• 10-14 years ago<br>• 15-19 years ago<br>• 20+ years ago | • Yes | • Yes | • Yes | • Yes | • Yes | • Yes | • Yes | • Yes | • Yes | • Yes |
| No | • Never | • No | • No | • No | • No | • No | • No | • No | • Missing | • No | • No |
| Not known | • Missing | • Missing | • Missing | • Missing | • Missing | • Missing | • Missing | • Missing | | • Missing | • Missing/do not know/prefer not to answer |
| | **Menopausal status (women only)** | | | | | | | | | | |
| | **Derived from multiple questions** | **Derived from multiple questions** | **Derived from multiple questions** | **How many natural periods have you had in the last 12 months?** | **Derived using age at recruitment** | **Have you had your menopause?** | **How many natural menstrual periods have you had in the last 12 months?** | **Have you had your menopause?** | **Have you had your menopause?** | **Derived from multiple questions** | **Have you had your menopause (periods stopped)?** |
| Premenopausal | | | | • 1 to 3<br>• 4 to 5<br>• 6 to 9<br>• 10 or more | <50 years | • No | • 1 to 3<br>• 4 to 5<br>• 6 to 9<br>• 10 or more | • No | • No | | |
| Postmenopausal | | | | • None | ≥50 years | • Yes | • None | • Yes, currently<br>• Yes, had menopause | • Not sure<br>• Yes | | • Yes |
| | **Ever used oral contraceptives (women only)** | | | | | | | | | | |

| | In the table below, please mark how long you took birth control pills during each of the stated age? | Hormonal drugs or oral contraceptive pills? | Have you used hormonal drugs or oral contraceptive pills? | Have you ever used contraceptives (the pill)? | Have you ever taken the contraceptive pill? | Have you ever used an oral contraceptive? | Have you ever used contraceptives (the pill)? | Have you ever used oral contraceptive pills? | Have you ever used the pill (oral contraceptive) ? | Have you ever taken oestrogen hormone pills?/ Have you ever taken progesterone or progestin hormone pills? | Have you ever taken the contraceptive pill? |
|---|---|---|---|---|---|---|---|---|---|---|---|
| Yes | • Yes, but only around menopause<br>• Yes, taking now<br>• Yes, but not taking them at this time | • Yes, current use<br>• Yes, past use | • Yes, current use<br>• Yes, past use | • Yes | • Yes | • Ever | • Yes | • Past use<br>• Current use | • Yes | • Yes, oestrogen use<br>• Yes, progesterone use | • Yes |
| No | • No | • No | • No | • No | • No | • Never | • No | • Never | • No | • No, oestrogen use<br>• No, progesterone use | • No |
| Not known | • Missing | • Missing | • Missing | • Missing | • Missing | • Missing | • Missing | • Missing | • Missing | • Missing | • Missing/do not know/prefer not to answer |
| | **Ever used hormone replacement therapy (women only)** | | | | | | | | | | |
| | **Have you ever used an oestrogen pill or patch around the time that your natural periods were stopping or after they stopped?** | | | **Have you ever used hormone replacement therapy?** | | **Have you ever supplemented female hormones?** | **Have you ever used hormone replacement therapy?** | | **Have you ever used hormone replacement therapy?** | **Are you currently taking replacement hormones?/ How many years have you taken replacement hormones?** | **Have you ever used hormone replacement therapy?** |
| Yes | • Yes, I am using oestrogens now<br>• Yes, I used them in the past, but not now | | | • Yes | | • Ever | • Yes | | • Past<br>• Current<br>• Ever | • Less than 5 years<br>• 5 to 9 years<br>• 10 or more years | • Yes |
| No | • No, I have never taken | | | • No | | • Never | • No | | • Never | • Never | • No |
| Not known | • Missing | | | • Missing | | • Missing | • Missing | | • Missing | • Missing | • Missing/do not know/prefer not to answer |

Abbreviations: AHS-2, Adventist Health Study-2; CARRS, Centre for cArdiometabolic Risk Reduction in South Asia; CKB, China Kadoorie Biobank; CSE, Certificate of Secondary Education; EPIC, European Prospective Investigation into Cancer and Nutrition; GCE, General Certificate of Education; GCSE, General Certificate of Secondary Education; HNC, Higher National Certificate; HND, Higher National Diploma; MWS, Million Women Study; NIH-AARP, National Institutes of Health-AARP Diet and Health Study; NVQ, National Vocational Qualification; OVS; Oxford Vegetarian Study; TCHS, Tzu Chi Health Study; UKWCS, UK Women's Cohort Study.

**Table S2.** List of food groups requested from the individual cohorts.

| **Food groups (g/day)** |
|---|
| Refined grains (e.g. white pasta/noodles/couscous, bread and rice) |
| Wholegrains (e.g. wholemeal pasta/noodles/couscous and bread, and brown rice) |
| Meat substitute dishes (e.g. vegetable burgers, textured vegetable proteins, soy-based foods) |
| Total vegetables |
| Potatoes |
| Legumes (green beans, sugar snap peas, pulses, etc.) |
| Fresh fruit |
| Fruit smoothie or fruit juice |
| Nuts and seeds |
| Sweets, preserves, cakes & confectionery (e.g. biscuits, desserts, chocolate) |
| Tea and/or coffee |
| Plant milks |
| Sugar sweetened beverages, including diet versions |
| Red meat |
| Processed meat |
| Poultry |
| Offal |
| Oily fish |
| Non-oily fish |
| Total fish (including shellfish) |
| Dairy milk |
| Cheese |
| Yogurt |
| Total dairy products (includes other dairy products, e.g. butter, dairy ice cream) |
| Egg & egg dishes |

**Table S3.** Food group intake criteria for the classification of diet groups in each cohort.

| | Diet groups | | | | | |
|---|---|---|---|---|---|---|
| **Food group intake** | **Poultry eaters** | **Pescatarians** | **Lacto-ovo vegetarians** | **Lacto vegetarians** | **Ovo vegetarians** | **Vegans** |
| | **Adventist Health Study–2** | | | | | |
| Red and processed meat | Never/rarely | Never/rarely | Never/rarely | Never/rarely | Never/rarely | Never/rarely |
| Poultry | ≥1–3/month | Never/rarely | Never/rarely | Never/rarely | Never/rarely | Never/rarely |
| Fish | | ≥1–3/month | Never/rarely | Never/rarely | Never/rarely | Never/rarely |
| Dairy products | | | ≥1–3/month | ≥1–3/month | Never/rarely | Never/rarely |
| Eggs | | | ≥1–3/month | Never/rarely | ≥1–3/month | Never/rarely |
| | **Centre for cArdiometabolic Risk Reduction in South Asia–1** | | | | | |
| Red and processed meat | Never/<1/month | Never/<1/month | Never/<1/month | Never/<1/month | Never/<1/month | Never/<1/month |
| Poultry | ≥Monthly | Never/<1/month | Never/<1/month | Never/<1/month | Never/<1/month | Never/<1/month |
| Fish | | ≥Monthly | Never/<1/month | Never/<1/month | Never/<1/month | Never/<1/month |
| Dairy products | | | ≥Monthly | ≥Monthly | Never/<1/month | Never/<1/month |
| Eggs | | | ≥Monthly | Never/<1/month | ≥Monthly | Never/<1/month |
| | **Centre for cArdiometabolic Risk Reduction in South Asia–2** | | | | | |
| Red and processed meat | Never/<1/month | Never/<1/month | Never/<1/month | Never/<1/month | Never/<1/month | Never/<1/month |
| Poultry | ≥Monthly | Never/<1/month | Never/<1/month | Never/<1/month | Never/<1/month | Never/<1/month |
| Fish | | ≥Monthly | Never/<1/month | Never/<1/month | Never/<1/month | Never/<1/month |
| Dairy products | | | ≥Monthly | ≥Monthly | Never/<1/month | Never/<1/month |
| Eggs | | | ≥Monthly | Never/<1/month | ≥Monthly | Never/<1/month |
| | **European Prospective Investigation into Cancer and Nutrition–Oxford** | | | | | |
| Red and processed meat | Never | Never | Never | Never | Never | Never |
| Poultry | ≥1–3/month | Never | Never | Never | Never | Never |
| Fish | | Yes in question 1, or >Never | Never | Never | Never | Never |
| Dairy products | | | Yes in question 1, or >Never | Yes in question 1, or >Never | Never | Never |
| Eggs | | | Yes in question 1, or >Never | Never | Yes in question 1, or >Never | Never |
| | **Oxford Vegetarian Study** | | | | | |
| Red and processed meat | – | Never | Never | Never | Never | Never |
| Fish | – | >Never | Never | Never | Never | Never |
| Dairy products | | | >Never | >Never | Never | Never |
| Eggs | | | >Never | Never | >Never | Never |

| | | | | | | |
|---|---|---|---|---|---|---|
| | **Tzu Chi Health Study** | | | | | |
| Red and processed meat | No intake or <1/month | No intake or <1/month | No intake or <1/month | No intake or <1/month | No intake or <1/month | No intake or <1/month |
| Poultry | ≥1/month | No intake or <1/month | No intake or <1/month | No intake or <1/month | No intake or <1/month | No intake or <1/month |
| Fish | | ≥1/month | No intake or <1/month | No intake or <1/month | No intake or <1/month | No intake or <1/month |
| Dairy products | | | ≥1/month | ≥1/month | No intake or <1/month | No intake or <1/month |
| Eggs | | | ≥1/month | No intake or <1/month | ≥1/month | No intake or <1/month |
| | **UK Women's Cohort Study** | | | | | |
| Red and processed meat | Never | Never | Never | Never | Never | Never |
| Poultry | >Never | Never | Never | Never | Never | Never |
| Fish | | >Never | Never | Never | Never | Never |
| Dairy products | | | >Never | >Never | Never | Never |
| Eggs | | | >Never | Never | >Never | Never |
| | **China Kadoorie Biobank** | | | | | |
| Red and processed meat | Never/rarely | Never/rarely | Never/rarely | Never/rarely | Never/rarely | Never/rarely |
| Poultry | ≥Monthly | Never/rarely | Never/rarely | Never/rarely | Never/rarely | Never/rarely |
| Fish | | ≥Monthly | Never/rarely | Never/rarely | Never/rarely | Never/rarely |
| Dairy products | | | ≥Monthly | ≥Monthly | Never/rarely | Never/rarely |
| Eggs | | | ≥Monthly | Never/rarely | ≥Monthly | Never/rarely |
| | **Million Women Study** | | | | | |
| Red and processed meat | Never | Never | Never | Never | Never | Never |
| Poultry | ≥1/week | Never | Never | Never | Never | Never |
| Fish | | ≥1/week | Never | Never | Never | Never |
| Dairy products | | | ≥1/week | ≥1/week | Never | Never |
| Eggs | | | ≥1/week | Never | ≥1/week | Never |
| | **National Institutes of Health-AARP Diet and Health Study** | | | | | |
| Red and processed meat | ≤10 g/day | ≤10 g/day | ≤10 g/day | ≤10 g/day | ≤10 g/day | ≤10 g/day |
| Poultry | >0 g/day | 0 g/day | 0 g/day | 0 g/day | 0 g/day | 0 g/day |
| Fish | | >0 g/day | 0 g/day | 0 g/day | 0 g/day | 0 g/day |
| Dairy products | | | >5 g/day | >5 g/day | ≤5 g/day | ≤5 g/day |
| Eggs | | | >0 g/day | 0 g/day | >0 g/day | 0 g/day |
| | **UK Biobank** | | | | | |
| Red and processed meat | Never | Never | Never | Never | Never | Never |
| Poultry | >Never | Never | Never | Never | Never | Never |

| | | | | | |
|---|---|---|---|---|---|
| Fish | >Never | Never | Never | Never | Never |
| Dairy products | | >Never | >Never | Never | Never |
| Eggs | | >Never | Never | >Never | Never |

**Table S4.** Number of participants in each diet group by sex and cohort (n = 2,337,152).

| Cohort | Meat eaters | Poultry eaters | Pescatarians | Vegetarians | | | | Vegans |
|---|---|---|---|---|---|---|---|---|
| | | | | Lacto-ovo vegetarians | Lacto vegetarians | Ovo vegetarians | All vegetarians | |
| | **Women, N (%)** | | | | | | | |
| ***Cohorts with large proportions of vegetarians*** | | | | | | | | |
| Adventist Health Study-2 | 14,711 (34.9) | 7,451 (17.7) | 4,215 (10.0) | 8,156 (19.3) | 3,826 (9.1) | 446 (1.1) | 12,428 (29.5) | 3,389 (8.0) |
| CARRS-1 | 3,561 (55.9) | 1,036 (16.3) | 123 (1.9) | 199 (3.1) | 1,113 (17.5) | 56 (0.88) | 1,368 (21.5) | 284 (4.5) |
| CARRS-2 | 2,292 (45.3) | 1,077 (21.3) | 111 (2.2) | 159 (3.1) | 1,249 (24.7) | 29 (0.57) | 1,437 (28.4) | 148 (2.9) |
| EPIC-Oxford | 20,156 (48.8) | 1,404 (3.4) | 6,685 (16.2) | 8,708 (21.1) | 3,046 (7.4) | 74 (0.18) | 11,828 (28.6) | 1,262 (3.1) |
| Oxford Vegetarian Study[1] | 2,939 (45.4) | - | 724 (11.2) | 2,367 (36.5) | 232 (3.6) | 38 (0.59) | 2,637 (40.7) | 180 (2.8) |
| Tzu Chi Health Study | 1,827 (54.9) | 84 (2.5) | 162 (4.9) | 967 (29.0) | 183 (5.5) | 73 (2.2) | 1,223 (36.7) | 33 (0.99) |
| UK Women's Cohort Study | 21,180 (70.3) | 929 (3.1) | 3,804 (12.6) | 3,300 (10.9) | 743 (2.5) | 32 (0.11) | 4,075 (13.5) | 160 (0.53) |
| ***Very large cohort studies*** | | | | | | | | |
| China Kadoorie Biobank | 284,030 (94.4) | 1,302 (0.43) | 1,290 (0.43) | 1,536 (0.51) | 563 (0.19) | 8,504 (2.8) | 10,603 (3.5) | 3,684 (1.2) |
| Million Women Study | 618,450 (96.8) | 3,414 (0.53) | 10,947 (1.7) | 5,361 (0.84) | 742 (0.12) | 33 (0.01) | 6,136 (0.96) | 79 (0.01) |
| NIH-AARP | 193,341 (89.5) | 20,664 (9.6) | 981 (0.45) | 483 (0.22) | 370 (0.17) | 10 (0.00) | 863 (0.40) | 56 (0.03) |
| UK Biobank | 237,928 (93.1) | 4,100 (1.6) | 7,692 (3.0) | 4,778 (1.9) | 575 (0.23) | 117 (0.05) | 5,470 (2.1) | 264 (0.10) |
| **Total** | 1,400,415 (90.6) | 41,461 (2.7) | 36,734 (2.4) | 36,014 (2.3) | 12,642 (0.82) | 9,412 (0.61) | 58,068 (3.8) | 9,539 (0.62) |
| | **Men, N (%)** | | | | | | | |
| ***Cohorts with large proportions of vegetarians*** | | | | | | | | |
| Adventist Health Study-2 | 8,534 (38.2) | 3,043 (13.6) | 1,987 (8.9) | 4,642 (20.8) | 2,089 (9.3) | 230 (1.0) | 6,961 (31.1) | 1,836 (8.2) |
| CARRS-1 | 3,653 (62.5) | 922 (15.8) | 57 (0.98) | 166 (2.8) | 807 (13.8) | 73 (1.2) | 1,046 (17.9) | 168 (2.9) |
| CARRS-2 | 2,494 (55.9) | 861 (19.3) | 56 (1.3) | 164 (3.7) | 812 (18.2) | 18 (0.40) | 994 (22.3) | 60 (1.3) |
| EPIC-Oxford | 6,342 (51.1) | 299 (2.4) | 1,443 (11.6) | 2,682 (21.6) | 902 (7.3) | 21 (0.17) | 3,605 (29.0) | 728 (5.9) |
| Oxford Vegetarian Study[1] | 2,241 (55.4) | - | 274 (6.8) | 1,209 (29.9) | 156 (3.9) | 14 (0.35) | 1,379 (34.1) | 153 (3.8) |
| Tzu Chi Health Study | 1,692 (77.2) | 36 (1.6) | 71 (3.2) | 310 (14.1) | 51 (2.3) | 20 (0.91) | 381 (17.3) | 11 (0.50) |
| ***Very large cohort studies*** | | | | | | | | |
| China Kadoorie Biobank | 201,766 (96.4) | 647 (0.31) | 413 (0.20) | 916 (0.44) | 167 (0.08) | 3,901 (1.9) | 4,984 (2.4) | 1,426 (0.68) |
| NIH-AARP | 297,757 (95.5) | 12,296 (3.9) | 709 (0.23) | 486 (0.16) | 472 (0.15) | 4 (0.00) | 962 (0.31) | 62 (0.02) |
| UK Biobank | 211,243 (96.6) | 1,338 (0.61) | 3,036 (1.4) | 2,362 (1.1) | 365 (0.17) | 58 (0.03) | 2,785 (1.3) | 184 (0.08) |
| **Total** | 735,722 (93.0) | 19,442 (2.5) | 8,046 (1.0) | 12,937 (1.6) | 5,821 (0.74) | 4,339 (0.55) | 23,097 (2.9) | 4,628 (0.59) |

Abbreviations: CARRS, Centre for cArdiometabolic Risk Reduction in South Asia; EPIC, European Prospective Investigation into Cancer and Nutrition; NIH-AARP, National Institutes of Health-AARP Diet and Health Study.

[1]In the Oxford Vegetarian Study, poultry eaters could not be determined as poultry intake was not assessed.

**Table S5.** Baseline characteristics of women by cohort[1].

| | Total | AHS-2 | CARRS-1 | CARRS-2 | EPIC-Oxford | OVS | TCHS | UKWCS | CKB | MWS | NIH-AARP | UK Biobank |
|---|---|---|---|---|---|---|---|---|---|---|---|---|
| N | 1,546,217 | 42,194 | 6,372 | 5,065 | 41,335 | 6,480 | 3,329 | 30,148 | 300,909 | 639,026 | 215,905 | 255,454 |
| Living with partner | | | | | | | | | | | | |
| Yes | 1,145,967 (74.1) | 27,503 (65.2) | 5,671 (89.0) | 4,332 (85.5) | 27,614 (66.8) | 3,497 (54.0) | 2824 (84.8) | 22,282 (73.9) | 267,799 (89.0) | 511,789 (80.1) | 95,749 (44.3) | 176,907 (69.3) |
| No | 336,711 (21.8) | 13,937 (33.0) | 701 (11.0) | 733 (14.5) | 13,649 (33.0) | 2,934 (45.3) | 504 (15.1) | 7,415 (24.6) | 33,110 (11.0) | 118,337 (18.5) | 118,140 (54.7) | 27,251 (10.7) |
| Unknown | 63,539 (4.1) | 754 (1.8) | 0 (0.00) | 0 (0.00) | 72 (0.17) | 49 (0.76) | 1 (0.00) | 451 (1.5) | 0 (0.00) | 8,900 (1.4) | 2,016 (0.93) | 51,296 (20.1) |
| Educational status | | | | | | | | | | | | |
| Less than secondary/high school | 514,795 (33.3) | 3,052 (7.2) | 1,568 (24.6) | 1,271 (25.1) | 5,426 (13.1) | 309 (4.8) | 1023 (30.7) | 4,671 (15.5) | 170,715 (56.7) | 217,050 (34.0) | 67,342 (31.2) | 42,368 (16.6) |
| Secondary/high school or equivalent | 572,012 (37.0) | 18,125 (43.0) | 3,859 (60.6) | 2,908 (57.4) | 16,422 (39.7) | 4,573 (70.6) | 1636 (49.1) | 15,539 (51.5) | 116,872 (38.8) | 305,150 (47.8) | 22,927 (10.6) | 64,001 (25.1) |
| University degree or equivalent | 430,226 (27.8) | 20,431 (48.4) | 945 (14.8) | 884 (17.5) | 16,406 (39.7) | 434 (6.7) | 670 (20.1) | 7,372 (24.5) | 13,322 (4.4) | 106,496 (16.7) | 118,637 (54.9) | 144,629 (56.6) |
| Unknown | 29,184 (1.9) | 586 (1.4) | 0 (0.00) | 2 (0.04) | 3,081 (7.5) | 1,164 (18.0) | 0 (0.00) | 2,566 (8.5) | 0 (0.00) | 10,330 (1.6) | 6,999 (3.2) | 4,456 (1.7) |
| Cigarette smoking | | | | | | | | | | | | |
| Never | 974,560 (63.0) | 35,304 (83.7) | 6,094 (95.6) | 4,985 (98.4) | 25,457 (61.6) | 3,595 (55.5) | 3274 (98.3) | 16,851 (55.9) | 285,673 (94.9) | 346,928 (54.3) | 94,000 (43.5) | 152,399 (59.7) |
| Previous | 406,326 (26.3) | 6,166 (14.6) | 5 (0.08) | 21 (0.41) | 11,253 (27.2) | 1,809 (27.9) | 40 (1.20) | 9,148 (30.3) | 2,619 (0.87) | 211,953 (33.2) | 84,058 (38.9) | 79,254 (31.0) |
| Current | 146,379 (9.5) | 410 (0.97) | 58 (0.91) | 59 (1.2) | 4,439 (10.7) | 1,049 (16.2) | 15 (0.45) | 3,224 (10.7) | 12,617 (4.2) | 71,277 (11.2) | 30,343 (14.1) | 22,888 (9.0) |
| Unknown | 18,952 (1.2) | 314 (0.74) | 215 (3.4) | 0 (0.00) | 186 (0.45) | 27 (0.42) | 0 (0.00) | 925 (3.1) | 0 (0.00) | 8,868 (1.4) | 7,504 (3.5) | 913 (0.36) |
| Physical activity | | | | | | | | | | | | |
| Inactive | 357,643 (23.1) | 8,553 (20.3) | 2,681 (42.1) | 3,815 (75.3) | 23,253 (56.3) | 2,596 (40.1) | 1248 (37.5) | 22,033 (73.1) | 99,953 (33.2)[2] | 36,813 (5.8) | 79,280 (36.7) | 77,418 (30.3) |
| Moderately active | 426,785 (27.6) | 13,711 (32.5) | 797 (12.5) | 491 (9.7) | 6,931 (16.8) | 1,871 (28.9) | 1082 (32.5) | 7,074 (23.5) | 100,654 (33.4)[2] | 70,898 (11.1) | 99,164 (45.9) | 124,112 (48.6) |
| Highly active | 570,911 (36.9) | 17,753 (42.1) | 1,494 (23.4) | 40 (0.79) | 4,770 (11.5) | 1,831 (28.3) | 999 (30.0) | 1,032 (3.4) | 100,302 (33.3)[2] | 364,921 (57.1) | 34,746 (16.1) | 43,023 (16.8) |
| Unknown | 190,878 (12.3) | 2,177 (5.2) | 1,400 (22.0) | 719 (14.2) | 6,381 (15.4) | 182 (2.8) | 0 (0.00) | 9 (0.03) | 0 (0.00) | 166,394 (26.0) | 2,715 (1.3) | 10,901 (4.3) |
| History of diabetes | | | | | | | | | | | | |
| Yes | 71,748 (4.6) | 3,195 (7.6) | 777 (12.2) | 830 (16.4) | 459 (1.1) | 33 (0.51) | 155 (4.7) | 544 (1.8) | 18,426 (6.1) | 21,570 (3.4) | 16,204 (7.5) | 9,555 (3.7) |
| No | 1,466,761 (94.9) | 38,512 (91.3) | 5,569 (87.4) | 4,231 (83.5) | 36,684 (88.7) | 6,419 (99.1) | 3174 (95.3) | 27,293 (90.5) | 282,483 (93.9) | 617,456 (96.6) | 199,701 (92.5) | 245,239 (96.0) |

| | | | | | | | | | | | | |
|---|---|---|---|---|---|---|---|---|---|---|---|---|
| Unknown | 7,708 (0.50) | 487 (1.2) | 26 (0.41) | 4 (0.08) | 4,192 (10.1) | 28 (0.43) | 0 (0.00) | 2,311 (7.7) | 0 (0.00) | 0 (0.00) | 0 (0.00) | 660 (0.26) |
| Age at menarche | | | | | | | | | | | | |
| ≤10 years | 60,622 (3.9) | 3,658 (8.7) | 16 (0.25) | 18 (0.36) | 1,800 (4.4) | - | 3 (0.09) | 1,760 (5.8) | 278 (0.09) | 26,924 (4.2) | 14,720 (6.8) | 11,445 (4.5) |
| 11–12 years | 457,022 (29.6) | 18,046 (42.8) | 759 (11.9) | 635 (12.5) | 14,635 (35.4) | - | 282 (8.47) | 10,787 (35.8) | 16,130 (5.4) | 220,840 (34.6) | 90,177 (41.8) | 84,731 (33.2) |
| 13–14 years | 624,908 (40.4) | 15,807 (37.5) | 3,931 (61.7) | 2,817 (55.6) | 18,925 (45.8) | - | 1482 (44.5) | 13,111 (43.5) | 84,506 (28.1) | 285,866 (44.7) | 89,002 (41.2) | 109,461 (42.8) |
| ≥15 years | 375,784 (24.3) | 4,365 (10.3) | 1,664 (26.1) | 1,570 (31.0) | 5,399 (13.1) | - | 1536 (46.1) | 3,877 (12.9) | 199,947 (66.4) | 95,559 (15.0) | 19,860 (9.2) | 42,007 (16.4) |
| Unknown | 27,881 (1.8) | 318 (0.75) | 2 (0.03) | 25 (0.49) | 576 (1.4) | 6,480 (100) | 26 (0.8) | 613 (2.0) | 48 (0.02) | 9,837 (1.5) | 2,146 (0.99) | 7,810 (3.1) |
| Parity | | | | | | | | | | | | |
| None | 187,425 (12.1) | 6,542 (15.5) | 329 (5.2) | 295 (5.8) | 15,748 (38.1) | 3,244 (50.1) | 255 (7.66) | 3,739 (12.4) | 3,956 (1.3) | 72,831 (11.4) | 32,816 (15.2) | 47,670 (18.7) |
| One | 254,901 (16.5) | 5,457 (12.9) | 546 (8.6) | 385 (7.6) | 5,357 (13.0) | 917 (14.2) | 184 (5.53) | 3,748 (12.4) | 103,690 (34.5) | 78,095 (12.2) | 22,460 (10.4) | 34,062 (13.3) |
| Two | 593,107 (38.4) | 13,342 (31.6) | 1,692 (26.6) | 1,287 (25.4) | 12,261 (29.7) | 1,386 (21.4) | 1148 (34.5) | 11,668 (38.7) | 95,596 (31.8) | 287,141 (44.9) | 55,974 (25.9) | 111,612 (43.7) |
| 3–4 | 430,594 (27.8) | 13,045 (30.9) | 2,489 (39.1) | 1,983 (39.2) | 7,062 (17.1) | 714 (11.0) | 1550 (46.6) | 7,255 (24.1) | 77,933 (25.9) | 182,369 (28.5) | 78,618 (36.4) | 57,576 (22.5) |
| 5–9 | 70,558 (4.6) | 3,066 (7.3) | 1,211 (19.0) | 1,042 (20.6) | 526 (1.3) | 63 (0.97) | 173 (5.20) | 603 (2.0) | 19,542 (6.5) | 17,126 (2.7) | 23,014 (10.7) | 4,192 (1.6) |
| ≥10 | 1,305 (0.08) | 133 (0.32) | 59 (0.93) | 52 (1.0) | 2 (0.00) | 1 (0.02) | 0 (0.00) | 12 (0.04) | 192 (0.06) | 63 (0.01) | 746 (0.35) | 45 (0.02) |
| Unknown | 8,327 (0.54) | 609 (1.4) | 46 (0.72) | 21 (0.41) | 379 (0.92) | 155 (2.4) | 19 (0.6) | 3,123 (10.4) | 0 (0.00) | 1,401 (0.22) | 2,277 (1.1) | 297 (0.12) |
| Age at first birth | | | | | | | | | | | | |
| ≤19 years | 157,185 (10.2) | 5,947 (14.1) | - | - | 1,489 (3.6) | 195 (3.0) | - | 1,661 (5.5) | 28,021 (9.3) | 62,803 (9.8) | 37,301 (17.3) | 19,768 (7.7) |
| 20–24 years | 619,652 (40.1) | 13,002 (30.8) | - | - | 7,934 (19.2) | 967 (14.9) | - | 8,101 (26.9) | 170,982 (56.8) | 258,825 (40.5) | 92,737 (43.0) | 67,104 (26.3) |
| 25–29 years | 401,728 (26.0) | 8,627 (20.4) | - | - | 9,889 (23.9) | 1,259 (19.4) | - | 8,883 (29.5) | 86,929 (28.9) | 173,713 (27.2) | 38,202 (17.7) | 74,226 (29.1) |
| 30–34 years | 108,882 (7.0) | 3,479 (8.2) | - | - | 4,444 (10.8) | 490 (7.6) | - | 3,434 (11.4) | 9,222 (3.1) | 45,920 (7.2) | 9,717 (4.5) | 32,176 (12.6) |
| 35–39 years | 30,023 (1.9) | 1,049 (2.5) | - | - | 1,248 (3.0) | 134 (2.1) | - | 1,025 (3.4) | 1,433 (0.48) | 11,223 (1.8) | 2,387 (1.1) | 11,524 (4.5) |
| ≥40 years | 5,511 (0.36) | 236 (0.56) | - | - | 193 (0.47) | 23 (0.35) | - | 157 (0.52) | 218 (0.07) | 2,002 (0.31) | 413 (0.19) | 2,269 (0.89) |
| No children | 187,425 (12.1) | 6,542 (15.5) | 329 (5.2) | 295 (5.8) | 15,748 (38.1) | 3,244 (50.1) | 255 (7.7) | 3,739 (12.4) | 3,956 (1.3) | 72,831 (11.4) | 32,816 (15.2) | 47,670 (18.7) |
| Unknown | 35,811 (2.3) | 3,312 (7.8) | 6,043 (94.8) | 4,770 (94.2) | 390 (0.94) | 168 (2.6) | 3074 (92.3) | 3,148 (10.4) | 148 (0.05) | 11,709 (1.8) | 2,332 (1.1) | 717 (0.28) |

| | | | | | | | | | | | | |
|---|---|---|---|---|---|---|---|---|---|---|---|---|
| Menopausal status | | | | | | | | | | | | |
| Pre-menopausal | 244,562 (15.8) | 9,689 (23.0) | 4,018 (63.1) | 2,941 (58.1) | 24,718 (59.8) | 4,754 (73.4) | 1232 (37.0) | 11,796 (39.1) | 128,741 (42.8) | 1 (0.00) | 0 (0.00) | 56,672 (22.2) |
| Post-menopausal | 1,301,655 (84.2) | 32,505 (77.0) | 2,354 (36.9) | 2,124 (41.9) | 16,617 (40.2) | 1,726 (26.6) | 2097 (63.0) | 18,352 (60.9) | 172,168 (57.2) | 639,025 (100.0) | 215,905 (100) | 198,782 (77.8) |
| Age at menopause (postmenopausal women only) | | | | | | | | | | | | |
| <40 years | 77,087 (5.9) | 2,594 (8.0) | 406 (17.2) | 292 (13.7) | 0 (0.00) | - | 131 (6.2) | 2,004 (10.9) | 6,464 (3.8) | 21,158 (3.3) | 38,407 (17.8) | 5,631 (2.8) |
| 40–44 years | 124,498 (9.6) | 3,005 (9.2) | 639 (27.1) | 518 (24.4) | 1,508 (9.1) | - | 251 (12.0) | 2,008 (10.9) | 18,184 (10.6) | 52,184 (8.2) | 33,393 (15.5) | 12,808 (6.4) |
| 45–49 years | 296,861 (22.8) | 5,383 (16.6) | 757 (32.2) | 653 (30.7) | 3,142 (18.9) | - | 556 (26.5) | 3,953 (21.5) | 65,017 (37.8) | 132,074 (20.7) | 51,357 (23.8) | 33,969 (17.1) |
| 50–54 years | 438,854 (33.7) | 7,762 (23.9) | 340 (14.4) | 375 (17.7) | 4,893 (29.4) | - | 964 (46.0) | 6,294 (34.3) | 60,141 (34.9) | 223,470 (35.0) | 66,050 (30.6) | 68,565 (34.5) |
| ≥55 years | 104,681 (8.0) | 12,232 (37.6) | 103 (4.4) | 73 (3.4) | 739 (4.4) | - | 173 (8.2) | 1,051 (5.7) | 7,779 (4.5) | 46,200 (7.2) | 14,617 (6.8) | 21,714 (10.9) |
| Unknown | 259,674 (19.9) | 1,529 (4.7) | 109 (4.6) | 213 (10.0) | 6,335 (38.1) | 1,726 (100) | 22 (1.0) | 3,042 (16.6) | 14,583 (8.5) | 163,939 (25.7) | 12,081 (5.6) | 56,095 (28.2) |
| Ever used oral contraceptives | | | | | | | | | | | | |
| Yes | 782,773 (50.6) | 24,802 (58.8) | 223 (3.5) | 349 (6.9) | 30,475 (73.7) | 3,582 (55.3) | 523 (15.7) | 20,167 (66.9) | 29,611 (9.8) | 391,488 (61.3) | 74,501 (34.5) | 207,052 (81.1) |
| No | 621,155 (40.2) | 17,392 (41.2) | 6,148 (96.5) | 4,688 (92.6) | 10,503 (25.4) | 2,848 (44.0) | 2778 (83.4) | 9,521 (31.6) | 271,252 (90.1) | 243,805 (38.2) | 4,671 (2.2) | 47,549 (18.6) |
| Unknown | 142,289 (9.2) | - | 1 (0.02) | 28 (0.55) | 357 (0.86) | 50 (0.77) | 28 (0.01) | 460 (1.5) | 46 (0.02) | 3,733 (0.58) | 136,733 (63.3) | 853 (0.33) |
| Ever used hormone replacement therapy | | | | | | | | | | | | |
| Yes | 584,677 (37.8) | 16,619 (39.4) | - | - | 7,506 (18.2) | - | 654 (19.6) | 8,198 (27.2) | - | 341,137 (53.4) | 114,186 (52.9) | 96,377 (37.7) |
| No | 628,851 (40.7) | 24,294 (57.6) | - | - | 33,239 (80.4) | - | 1432 (43.0) | 20,997 (69.6) | - | 289,096 (45.2) | 101,719 (47.1) | 158,074 (61.9) |
| Unknown | 332,689 (21.5) | 1,281 (3.0) | 6,372 (100) | 5,065 (100) | 590 (1.4) | 6,480 (100) | 1243 (37.3) | 953 (3.2) | 300,909 (100) | 8,793 (1.4) | 0 (0.00) | 1,003 (0.39) |

Abbreviations: AHS-2, Adventist Health Study-2; CARRS, Centre for cArdiometabolic Risk Reduction in South Asia; CKB, China Kadoorie Biobank; EPIC, European Prospective Investigation into Cancer and Nutrition; MWS, Million Women Study; NIH-AARP, National Institutes of Health-AARP Diet and Health Study; OVS, Oxford Vegetarian Study; TCHS, Tzu Chi Health Study; UKWCS, UK Women's Cohort Study.

[1]All values are N (%).

[2]Sex-specific tertiles of metabolic equivalents.

c indicates that no information was available for this variable in the specified cohort.

**Table S6.** Baseline characteristics of men by cohort.

| | Total | AHS-2 | CARRS-1 | CARRS-2 | EPIC-Oxford | OVS | TCHS | CKB | NIH-AARP | UK Biobank |
|---|---|---|---|---|---|---|---|---|---|---|
| N | 790,935 | 22,361 | 5,846 | 4,465 | 12,417 | 4,047 | 2,191 | 209,236 | 311,786 | 218,586 |
| Living with partner | | | | | | | | | | |
| Yes | 670,475 (84.8) | 19,077 (85.3) | 5,225 (89.4) | 3,924 (87.9) | 8,885 (71.6) | 2,449 (60.5) | 2,012 (91.8) | 194,417 (92.9) | 267,667 (85.8) | 166,819 (76.3) |
| No | 79,323 (10.0) | 2,946 (13.2) | 621 (10.6) | 540 (12.1) | 3,505 (28.2) | 1,582 (39.1) | 178 (8.1) | 14,819 (7.1) | 42,262 (13.6) | 12,870 (5.9) |
| Unknown | 41,137 (5.2) | 338 (1.5) | 0 (0.00) | 1 (0.02) | 27 (0.22) | 16 (0.40) | 1 (0.00) | 0 (0.00) | 1,857 (0.60) | 38,897 (17.8) |
| Educational status | | | | | | | | | | |
| Less than secondary/high school | 197,732 (25.0) | 1,553 (6.9) | 744 (12.7) | 504 (11.3) | 1,529 (12.3) | 34 (0.84) | 354 (16.2) | 88,290 (42.2) | 67,682 (21.7) | 37,042 (16.9) |
| Secondary/high school or equivalent | 195,929 (24.8) | 7,867 (35.2) | 3,850 (65.9) | 2,892 (64.8) | 3,598 (29.0) | 2,604 (64.3) | 1,045 (47.7) | 104,506 (49.9) | 29,002 (9.3) | 40,565 (18.6) |
| University degree or equivalent | 383,487 (48.5) | 12,713 (56.9) | 1,252 (21.4) | 1,069 (23.9) | 6,409 (51.6) | 640 (15.8) | 792 (36.2) | 16,440 (7.9) | 207,254 (66.5) | 136,918 (62.6) |
| Unknown | 13,787 (1.7) | 228 (1.0) | 0 (0.00) | 0 (0.00) | 881 (7.1) | 769 (19.0) | 0 (0.00) | 0 (0.00) | 7,848 (2.5) | 0 (0.00) |
| Cigarette smoking | | | | | | | | | | |
| Never | 260,434 (32.9) | 16,316 (73.0) | 3,420 (58.5) | 3,065 (68.6) | 6,501 (52.4) | 1,668 (41.2) | 1,328 (60.6) | 30,156 (14.4) | 90,924 (29.2) | 107,056 (49.0) |
| Previous | 299,946 (37.9) | 5,172 (23.1) | 209 (3.6) | 311 (7.0) | 4,225 (34.0) | 1,364 (33.7) | 647 (29.5) | 27,486 (13.1) | 177,428 (56.9) | 83,104 (38.0) |
| Current | 216,513 (27.4) | 258 (1.2) | 1,628 (27.8) | 1,086 (24.3) | 1,648 (13.3) | 1,006 (24.9) | 216 (9.9) | 151,594 (72.5) | 31,497 (10.1) | 27,580 (12.6) |
| Unknown | 14,042 (1.8) | 615 (2.8) | 589 (10.1) | 3 (0.07) | 43 (0.35) | 9 (0.22) | 0 (0.00) | 0 (0.00) | 11,937 (3.8) | 0 (0.00) |
| Physical activity | | | | | | | | | | |
| Inactive | 235,008 (29.7) | 3,588 (16.0) | 1,990 (34.0) | 3,164 (70.9) | 6,736 (54.2) | 1,380 (34.1) | 672 (30.7) | 69,414 (33.2)[2] | 87,726 (28.1) | 60,338 (27.6) |
| Moderately active | 343,204 (43.4) | 7,281 (32.6) | 1,362 (23.3) | 475 (10.6) | 2,383 (19.2) | 1,213 (30.0) | 685 (31.3) | 69,869 (33.4)[2] | 155,271 (49.8) | 104,665 (47.9) |
| Highly active | 198,300 (25.1) | 10,605 (47.4) | 1,097 (18.8) | 156 (3.5) | 2,201 (17.7) | 1,417 (35.0) | 834 (38.1) | 69,953 (33.4)[2] | 66,132 (21.2) | 45,905 (21.0) |
| Unknown | 14,423 (1.8) | 887 (4.0) | 1,397 (23.9) | 670 (15.0) | 1,097 (8.8) | 37 (0.91) | 0 (0.00) | 0 (0.00) | 2,657 (0.85) | 0 (0.00) |
| History of diabetes | | | | | | | | | | |
| Yes | 62,379 (7.9) | 1,880 (8.4) | 696 (11.9) | 696 (15.6) | 263 (2.1) | 29 (0.72) | 139 (6.3) | 11,579 (5.5) | 31,927 (10.2) | 15,170 (6.9) |
| No | 726,764 (91.9) | 20,189 (90.3) | 5,111 (87.4) | 3,762 (84.3) | 11,645 (93.8) | 4,009 (99.1) | 2,052 (97.7) | 197,657 (94.5) | 279,859 (89.8) | 202,480 (92.6) |
| Unknown | 1,792 (0.23) | 292 (1.3) | 39 (0.67) | 7 (0.16) | 509 (4.1) | 9 (0.22) | 0 (0.00) | 0 (0.00) | 0 (0.00) | 936 (0.43) |

Abbreviations: AHS-2, Adventist Health Study-2; CARRS, Centre for cArdiometabolic Risk Reduction in South Asia; CKB, China Kadoorie Biobank; EPIC, European Prospective Investigation into Cancer and Nutrition; NIH-AARP, National Institutes of Health-AARP Diet and Health Study; OVS, Oxford Vegetarian Study; TCHS, Tzu Chi Health Study.

[1]All values are N (%).

[2]Sex-specific tertiles of metabolic equivalents.

**Table S7.** Baseline food intakes by diet group and cohort (n = 2,337,152)[1].

| Cohort | Meat eaters | Poultry eaters | Pescatarians | Vegetarians | | | | Vegans |
|---|---|---|---|---|---|---|---|---|
| | | | | Lacto-ovo vegetarians | Lacto vegetarians | Ovo vegetarians | All vegetarians[2] | |
| | **Refined grains, g/day** | | | | | | | |
| ***Cohorts with large proportions of vegetarians*** | | | | | | | | |
| Adventist Health Study-2 | 107.2 (100.0) | 93.6 (91.7) | 93.6 (98.3) | 86.9 (78.2) | 76.5 (75.0) | 75.0 (80.8) | 83.3 (77.5) | 67.8 (78.7) |
| CARRS-1 | 403.7 (201.0) | 432.0 (180.6) | 452.3 (161.8) | 259.4 (237.2) | 209.7 (228.0) | 249.3 (240.8) | 219.4 (230.8) | 244.3 (236.8) |
| CARRS-2 | 737.0 (654.1) | 605.4 (592.7) | 718.8 (529.5) | 248.7 (388.5) | 192.9 (330.8) | 267.0 (323.1) | 201.7 (339.4) | 246.2 (336.7) |
| EPIC-Oxford | 91.1 (62.2) | 83.1 (70.3) | 95.1 (67.0) | 102.1 (70.0) | 93.5 (71.1) | 79.9 (71.9) | 99.8 (70.4) | 78.6 (76.0) |
| Oxford Vegetarian Study[3] | 110.4 (66.2) | - | 114.3 (65.9) | 116.9 (66.2) | 113.8 (72.5) | 132.8 (74.9) | 116.8 (66.9) | 134.5 (76.5) |
| Tzu Chi Health Study | 369.1 (247.8) | 304.4 (235.6) | 303.6 (239.5) | 349.0 (240.2) | 341.1 (265.6) | 345.4 (257.1) | 347.7 (244.9) | 374.8 (289.5) |
| UK Women’s Cohort Study | 91.6 (65.6) | 81.6 (69.3) | 94.1 (68.2) | 95.7 (69.6) | 83.5 (70.1) | 83.1 (74.9) | 93.4 (69.9) | 71.1 (72.1) |
| ***Very large cohorts*** | | | | | | | | |
| China Kadoorie Biobank[4] | 307.2 (69.4) | 301.5 (89.1) | 319.4 (101.5) | 252.8 (62.5) | 267.5 (80.8) | 238.8 (33.6) | 242.3 (43.3) | 237.9 (36.8) |
| Million Women Study | 97.3 (58.6) | 97.9 (63.6) | 103.9 (62.9) | 112.4 (64.1) | 108.7 (75.3) | 109.1 (68.4) | 111.9 (65.6) | 102.6 (63.5) |
| NIH-AARP | 91.6 (66.7) | 97.3 (83.9) | 133.5 (110.1) | 122.1 (88.5) | 144.2 (109.1) | 143.7 (117.1) | 132.4 (99.3) | 169.3 (109.3) |
| UK Biobank[4] | 147.4 (19.5) | 140.0 (14.9) | 139.9 (15.2) | 141.7 (16.5) | 145.5 (17.2) | 141.9 (14.3) | 142.2 (16.6) | 138.8 (14.4) |
| | **Wholemeal grains, g/day** | | | | | | | |
| ***Cohorts with large proportions of vegetarians*** | | | | | | | | |
| Adventist Health Study-2 | 126.2 (113.6) | 158.2 (127.9) | 180.8 (143.1) | 164.9 (122.0) | 196.0 (139.7) | 214.3 (143.2) | 176.1 (129.4) | 242.5 (157.3) |
| CARRS-1 | 81.9 (82.2) | 74.8 (78.6) | 62.1 (71.5) | 121.4 (88.4) | 129.7 (89.7) | 100.6 (90.0) | 126.9 (89.7) | 117.7 (92.3) |
| CARRS-2 | 80.2 (106.4) | 95.5 (111.7) | 64.7 (101.3) | 170.5 (113.2) | 179.4 (123.4) | 166.8 (118.1) | 178.0 (122.0) | 163.9 (113.1) |
| EPIC-Oxford | 82.7 (63.5) | 111.1 (73.5) | 112.5 (77.0) | 113.0 (76.0) | 116.2 (81.7) | 117.9 (83.0) | 113.8 (77.5) | 152.1 (95.3) |
| Oxford Vegetarian Study[3] | 24.4 (24.7) | - | 28.6 (24.3) | 30.0 (24.7) | 25.7 (25.4) | 20.8 (24.4) | 29.5 (24.8) | 26.8 (25.4) |
| Tzu Chi Health Study | 46.0 (60.3) | 54.6 (75.4) | 60.4 (60.8) | 65.2 (71.4) | 72.2 (84.4) | 68.2 (89.8) | 66.4 (74.6) | 101.7 (116.2) |
| UK Women’s Cohort Study | 75.8 (64.0) | 100.4 (72.0) | 100.7 (71.3) | 100.8 (71.4) | 104.5 (88.0) | 113.5 (84.1) | 101.5 (74.8) | 119.9 (79.4) |
| ***Very large cohorts*** | | | | | | | | |
| China Kadoorie Biobank[4] | 35.6 (43.7) | 51.1 (54.8) | 36.2 (38.8) | 73.2 (64.8) | 57.4 (58.7) | 95.1 (66.5) | 89.9 (66.7) | 86.1 (66.2) |
| Million Women Study | 81.6 (57.7) | 98.4 (63.2) | 102.3 (63.3) | 100.2 (63.5) | 90.7 (66.0) | 146.8 (97.3) | 99.3 (64.2) | 111.6 (77.1) |
| NIH-AARP | 48.2 (58.0) | 67.2 (80.3) | 79.6 (93.6) | 86.9 (92.0) | 92.9 (105.1) | 147.3 (296.1) | 90.1 (101.2) | 137.5 (143.1) |
| UK Biobank[4] | 85.9 (28.4) | 89.9 (25.8) | 94.6 (24.8) | 92.6 (26.8) | 76.5 (29.5) | 81.2 (30.5) | 90.6 (27.7) | 95.0 (28.2) |

| | | | | | | | | |
|---|---|---|---|---|---|---|---|---|
| | **Vegetable intake, g/day** | | | | | | | |
| ***Cohorts with large proportions of vegetarians*** | | | | | | | | |
| Adventist Health Study-2 | 264.9 (190.7) | 301.5 (213.5) | 342.6 (235.2) | 296.9 (183.9) | 320.4 (212.7) | 322.5 (215.9) | 304.9 (194.6) | 370.6 (253.8) |
| CARRS-1 | 420.7 (220.5) | 435.4 (212.0) | 433.0 (214.9) | 477.7 (179.0) | 443.6 (206.6) | 448.4 (204.1) | 449.0 (202.8) | 427.0 (218.6) |
| CARRS-2 | 355.5 (245.5) | 376.7 (233.5) | 359.3 (235.1) | 412.7 (219.3) | 406.5 (226.5) | 359.9 (175.7) | 406.4 (224.7) | 361.0 (225.6) |
| EPIC-Oxford | 230.6 (122.0) | 268.3 (144.9) | 273.9 (141.3) | 270.8 (141.9) | 277.5 (166.5) | 338.8 (174.5) | 272.9 (148.9) | 322.5 (176.9) |
| Oxford Vegetarian Study[3] | 91.5 (49.3) | - | 118.1 (55.3) | 119.7 (56.7) | 121.8 (62.3) | 129.4 (56.1) | 120.1 (57.3) | 122.6 (59.8) |
| Tzu Chi Health Study | 427.5 (291.9) | 491.2 (366.2) | 510.1 (300.5) | 519.6 (343.9) | 529.0 (374.0) | 502.1 (321.7) | 519.9 (347.1) | 633.4 (447.4) |
| UK Women's Cohort Study | 245.3 (127.3) | 305.0 (175.8) | 287.4 (150.6) | 277.4 (133.0) | 294.9 (173.1) | 317.0 (158.4) | 280.9 (141.5) | 342.3 (185.4) |
| ***Very large cohorts*** | | | | | | | | |
| China Kadoorie Biobank[4] | 236.6 (8.5) | 237.3 (6.0) | 237.5 (5.6) | 236.1 (8.8) | 236.0 (8.6) | 235.4 (10.4) | 235.6 (10.1) | 234.8 (12.0) |
| Million Women Study | 99.0 (67.6) | 108.2 (77.0) | 124.8 (84.6) | 134.6 (88.2) | 131.9 (103.9) | 198.7 (125.4) | 134.7 (90.6) | 175.9 (114.5) |
| NIH-AARP | 290.9 (183.0) | 321.2 (248.4) | 462.2 (361.0) | 445.8 (302.7) | 500.3 (331.1) | 509.1 (337.6) | 471.4 (317.4) | 801.7 (439.0) |
| UK Biobank[4] | 190.3 (46.5) | 204.7 (48.5) | 208.4 (47.5) | 209.2 (48.1) | 218.5 (50.3) | 219.5 (52.8) | 210.4 (48.6) | 224.6 (47.8) |
| | **Legumes, g/day** | | | | | | | |
| ***Cohorts with large proportions of vegetarians*** | | | | | | | | |
| Adventist Health Study-2 | 59.9 (68.0) | 77.2 (81.5) | 98.4 (100.7) | 89.4 (78.7) | 100.7 (90.1) | 95.5 (87.0) | 93.1 (82.8) | 109.5 (105.2) |
| CARRS-1 | 71.7 (85.5) | 66.0 (80.4) | 65.6 (80.9) | 134.9 (111.5) | 137.0 (111.6) | 125.1 (110.3) | 136.1 (111.5) | 117.3 (109.4) |
| CARRS-2 | 104.5 (112.0) | 106.2 (103.8) | 95.6 (96.2) | 136.1 (110.1) | 138.5 (108.0) | 92.4 (74.6) | 137.2 (107.9) | 143.9 (112.7) |
| EPIC-Oxford | 55.1 (37.2) | 58.8 (41.2) | 67.9 (46.1) | 74.6 (48.9) | 75.4 (60.5) | 89.2 (51.8) | 74.9 (52.2) | 97.8 (66.5) |
| Oxford Vegetarian Study[3] | 63.6 (34.4) | - | 83.8 (43.9) | 81.0 (42.2) | 86.9 (58.7) | 94.1 (57.4) | 81.7 (44.3) | 110.0 (55.6) |
| Tzu Chi Health Study | 15.0 (25.3) | 15.4 (25.8) | 18.1 (33.8) | 21.0 (37.3) | 17.7 (29.5) | 15.9 (31.6) | 20.2 (36.0) | 17.2 (20.6) |
| UK Women's Cohort Study | 65.9 (39.9) | 81.3 (50.8) | 86.4 (51.6) | 94.2 (55.3) | 90.7 (59.4) | 122.6 (62.4) | 93.8 (56.2) | 110.8 (64.6) |
| ***Very large cohorts*** | | | | | | | | |
| China Kadoorie Biobank[4] | NA | NA | NA | NA | NA | NA | NA | NA |
| Million Women Study | 18.0 (15.2) | 18.8 (17.7) | 21.4 (18.4) | 23.7 (20.6) | 23.5 (22.2) | 33.8 (25.3) | 23.8 (20.8) | 28.9 (24.9) |
| NIH-AARP | 25.5 (31.5) | 28.6 (45.5) | 62.5 (78.3) | 69.0 (80.7) | 79.6 (87.4) | 112.5 (114.4) | 74.2 (84.4) | 132.0 (135.4) |
| UK Biobank[4] | NA | NA | NA | NA | NA | NA | NA | NA |
| | **Fruit intake, g/day** | | | | | | | |
| ***Cohorts with large proportions of vegetarians*** | | | | | | | | |
| Adventist Health Study-2 | 258.7 (246.6) | 328.8 (285.3) | 377.1 (311.1) | 310.3 (235.6) | 372.6 (286.5) | 406.9 (305.1) | 332.7 (256.8) | 454.5 (321.3) |
| CARRS-1 | 38.4 (71.6) | 39.5 (73.0) | 38.7 (72.4) | 67.0 (89.2) | 67.8 (89.3) | 39.1 (73.3) | 66.1 (88.7) | 44.8 (77.0) |

| | | | | | | | | |
|---|---|---|---|---|---|---|---|---|
| CARRS-2 | 70.0 (93.7) | 61.6 (86.0) | 46.7 (70.6) | 104.8 (122.9) | 100.7 (130.3) | 34.6 (44.5) | 100.0 (128.5) | 32.4 (77.6) |
| EPIC-Oxford | 258.5 (202.5) | 315.0 (229.3) | 292.1 (228.7) | 268.6 (212.5) | 298.2 (242.9) | 315.2 (236.5) | 276.4 (221.2) | 351.0 (349.1) |
| Oxford Vegetarian Study[3] | 67.0 (37.5) | - | 83.5 (34.2) | 86.1 (33.6) | 82.9 (38.0) | 86.0 (31.8) | 85.8 (34.0) | 94.0 (32.2) |
| Tzu Chi Health Study | 153.3 (153.9) | 173.6 (146.2) | 175.8 (175.5) | 155.4 (136.0) | 157.9 (159.6) | 139.6 (236.8) | 154.9 (147.2) | 146.7 (118.1) |
| UK Women's Cohort Study | 308.7 (217.9) | 364.6 (260.1) | 351.7 (252.0) | 339.5 (243.3) | 369.4 (279.2) | 403.1 (236.7) | 345.4 (250.4) | 432.3 (369.6) |
| ***Very large cohorts*** | | | | | | | | |
| China Kadoorie Biobank[4] | 84.8 (35.1) | 91.6 (39.7) | 102.9 (41.6) | 74.2 (33.3) | 77.7 (36.9) | 62.5 (24.1) | 65.0 (27.0) | 59.0 (25.8) |
| Million Women Study | 281.2 (175.2) | 330.9 (208.1) | 345.7 (201.4) | 354.8 (214.8) | 363.1 (253.8) | 475.2 (252.4) | 356.5 (220.2) | 439.6 (220.9) |
| NIH-AARP | 356.9 (307.4) | 456.4 (376.5) | 465.4 (385.5) | 481.7 (364.0) | 508.9 (416.8) | 474.1 (446.1) | 494.2 (389.9) | 691.8 (634.7) |
| UK Biobank[4] | 191.5 (84.0) | 225.9 (88.8) | 214.7 (86.1) | 206.4 (87.8) | 209.1 (92.1) | 226.0 (94.0) | 207.2 (88.4) | 228.3 (92.6) |
| | **Nuts and seeds, g/day** | | | | | | | |
| ***Cohorts with large proportions of vegetarians*** | | | | | | | | |
| Adventist Health Study-2 | 17.6 (20.8) | 20.2 (23.3) | 23.6 (24.3) | 25.4 (24.2) | 27.8 (25.7) | 29.3 (27.9) | 26.2 (24.8) | 34.4 (31.1) |
| CARRS-1 | 4.5 (7.4) | 3.8 (7.1) | 3.1 (6.1) | 6.8 (9.7) | 7.2 (9.7) | 4.0 (7.7) | 6.9 (9.6) | 5.0 (8.7) |
| CARRS-2 | 2.9 (14.9) | 2.9 (16.4) | 1.2 (3.6) | 4.8 (17.0) | 4.1 (25.2) | 1.0 (3.2) | 4.1 (24.1) | 0.9 (3.1) |
| EPIC-Oxford | 4.4 (8.8) | 7.0 (12.2) | 7.9 (11.6) | 9.2 (13.7) | 9.9 (15.1) | 15.4 (23.6) | 9.5 (14.2) | 19.8 (26.0) |
| Oxford Vegetarian Study[3] | 8.5 (10.8) | - | 18.4 (15.2) | 19.3 (16.0) | 21.2 (17.8) | 29.1 (21.7) | 19.6 (16.3) | 29.7 (19.4) |
| Tzu Chi Health Study | 4.0 (9.7) | 2.7 (5.5) | 4.3 (11.5) | 5.8 (11.7) | 5.3 (10.3) | 4.1 (7.9) | 5.6 (11.4) | 6.4 (13.3) |
| UK Women's Cohort Study | 6.2 (9.0) | 9.5 (12.3) | 11.1 (14.3) | 11.7 (14.2) | 13.1 (18.7) | 17.4 (13.1) | 12.0 (15.1) | 21.4 (27.2) |
| ***Very large cohorts*** | | | | | | | | |
| China Kadoorie Biobank[4] | NA | NA | NA | NA | NA | NA | NA | NA |
| Million Women Study | 4.0 (8.9) | 6.9 (12.5) | 9.3 (14.0) | 10.9 (15.0) | 12.1 (17.9) | 19.0 (24.9) | 11.0 (15.5) | 20.7 (18.3) |
| NIH-AARP | 5.7 (10.8) | 4.0 (10.4) | 7.8 (16.1) | 11.6 (19.8) | 9.5 (17.3) | 10.0 (15.2) | 10.6 (18.7) | 14.6 (28.3) |
| UK Biobank[4] | NA | NA | NA | NA | NA | NA | NA | NA |
| | **Sweets, preserves, cakes & confectionery, g/day** | | | | | | | |
| ***Cohorts with large proportions of vegetarians*** | | | | | | | | |
| Adventist Health Study-2 | 23.2 (32.4) | 16.6 (24.8) | 14.8 (24.1) | 18.9 (24.8) | 14.8 (21.9) | 10.3 (17.7) | 17.4 (23.8) | 7.5 (15.0) |
| CARRS-1 | 32.9 (44.2) | 29.0 (45.0) | 35.2 (50.2) | 46.6 (49.6) | 50.7 (49.5) | 31.2 (42.0) | 49.0 (49.4) | 39.1 (47.8) |
| CARRS-2 | 19.8 (47.1) | 15.9 (35.9) | 13.6 (30.5) | 22.1 (43.7) | 21.8 (35.8) | 9.7 (20.0) | 21.6 (36.7) | 17.4 (173.3) |
| EPIC-Oxford | 79.0 (61.8) | 61.6 (54.2) | 65.1 (51.8) | 70.6 (55.0) | 63.2 (51.3) | 50.9 (53.8) | 68.6 (54.2) | 46.8 (43.0) |
| Oxford Vegetarian Study[3] | 33.9 (25.7) | - | 31.6 (24.7) | 32.4 (25.7) | 26.7 (24.9) | 29.3 (26.3) | 31.8 (25.7) | 23.4 (23.4) |
| Tzu Chi Health Study | 10.2 (18.4) | 6.1 (10.7) | 7.1 (14.2) | 12.2 (19.6) | 9.1 (17.1) | 0.0 (0.0) | 11.1 (18.9) | 0.0 (0.0) |

| | | | | | | | | |
|---|---|---|---|---|---|---|---|---|
| UK Women's Cohort Study | 77.0 (57.0) | 61.7 (52.5) | 64.3 (49.7) | 67.6 (51.4) | 54.8 (51.9) | 49.8 (62.6) | 65.1 (51.9) | 41.1 (45.6) |
| ***Very large cohorts*** | | | | | | | | |
| China Kadoorie Biobank[4] | NA | NA | NA | NA | NA | NA | NA | NA |
| Million Women Study | 43.2 (42.1) | 34.5 (38.5) | 36.8 (39.5) | 40.4 (40.8) | 30.5 (41.6) | 32.5 (43.2) | 39.1 (41.1) | 19.4 (41.6) |
| NIH-AARP | 44.1 (41.5) | 25.1 (31.6) | 29.8 (39.4) | 35.2 (40.1) | 30.0 (35.9) | 11.5 (15.1) | 32.6 (38.2) | 17.2 (27.1) |
| UK Biobank[4] | NA | NA | NA | NA | NA | NA | NA | NA |
| | **Plant milk, g/day** | | | | | | | |
| ***Cohorts with large proportions of vegetarians*** | | | | | | | | |
| Adventist Health Study-2 | 42.8 (108.9) | 87.6 (152.7) | 116.0 (162.4) | 100.2 (149.7) | 144.5 (177.8) | 159.0 (177.1) | 115.8 (161.2) | 162.1 (176.2) |
| CARRS-1 | NA | NA | NA | NA | NA | NA | NA | NA |
| CARRS-2 | NA | NA | NA | NA | NA | NA | NA | NA |
| EPIC-Oxford | 4.6 (38.2) | 18.4 (73.2) | 21.1 (78.7) | 25.5 (86.5) | 48.8 (119.3) | 184.9 (163.7) | 32.4 (97.8) | 221.7 (187.7) |
| Oxford Vegetarian Study[3] | NA | - | NA | NA | NA | NA | NA | NA |
| Tzu Chi Health Study | 79.6 (112.8) | 80.2 (103.4) | 73.9 (95.8) | 88.0 (126.8) | 77.2 (122.1) | 76.6 (139.8) | 85.7 (126.9) | 51.2 (83.2) |
| UK Women's Cohort Study | 2.6 (30.0) | 9.9 (58.6) | 15.2 (71.1) | 19.0 (76.0) | 44.1 (122.3) | 155.4 (206.7) | 24.7 (89.2) | 192.9 (166.9) |
| ***Very large cohorts*** | | | | | | | | |
| China Kadoorie Biobank[4] | NA | NA | NA | NA | NA | NA | NA | NA |
| Million Women Study | 4.3 (26.0) | 15.1 (47.5) | 18.1 (50.3) | 22.7 (57.8) | 29.8 (70.5) | 119.6 (102.8) | 24.1 (60.2) | 80.6 (94.5) |
| NIH-AARP | NA | NA | NA | NA | NA | NA | NA | NA |
| UK Biobank[4] | NA | NA | NA | NA | NA | NA | NA | NA |
| | **Sugar-sweetened beverages, g/day** | | | | | | | |
| ***Cohorts with large proportions of vegetarians*** | | | | | | | | |
| Adventist Health Study-2 | 232.2 (425.5) | 110.2 (292.6) | 58.5 (183.1) | 88.3 (236.4) | 56.3 (197.0) | 24.1 (122.5) | 76.3 (222.6) | 12.6 (100.1) |
| CARRS-1 | 51.9 (61.8) | 38.9 (56.8) | 46.3 (61.5) | 52.8 (64.0) | 54.0 (59.9) | 47.4 (59.4) | 53.5 (60.5) | 45.1 (56.2) |
| CARRS-2 | 24.5 (54.5) | 17.8 (37.7) | 18.1 (39.5) | 19.7 (37.9) | 19.1 (42.9) | 10.3 (20.3) | 19.0 (41.9) | 9.3 (20.9) |
| EPIC-Oxford | 88.6 (166.7) | 74.1 (157.3) | 74.3 (145.4) | 94.8 (166.1) | 95.4 (173.9) | 52.5 (146.5) | 94.7 (168.1) | 70.1 (149.2) |
| Oxford Vegetarian Study[3] | NA | - | NA | NA | NA | NA | NA | NA |
| Tzu Chi Health Study | 24.7 (103.7) | 7.7 (32.5) | 7.3 (44.3) | 14.5 (79.0) | 5.8 (28.0) | 18.8 (112.4) | 13.5 (76.3) | 0.4 (2.5) |
| UK Women's Cohort Study | 63.6 (123.6) | 50.6 (115.3) | 48.7 (105.6) | 54.0 (119.0) | 54.6 (129.0) | 32.9 (62.0) | 54.0 (120.6) | 55.1 (182.4) |
| ***Very large cohorts*** | | | | | | | | |
| China Kadoorie Biobank[4] | NA | NA | NA | NA | NA | NA | NA | NA |
| Million Women Study | 114.9 (207.8) | 104.1 (216.4) | 89.3 (184.3) | 97.9 (202.1) | 122.0 (279.3) | 169.7 (594.6) | 101.2 (216.9) | 77.0 (191.5) |

| | | | | | | | | |
|---|---|---|---|---|---|---|---|---|
| NIH-AARP | 895.7 (1,093.9) | 630.5 (905.3) | 588.0 (922.8) | 585.6 (881.6) | 639.5 (1,079.6) | 472.7 (804.5) | 609.6 (977.4) | 446.9 (660.6) |
| UK Biobank[4] | NA | NA | NA | NA | NA | NA | NA | NA |
| | **Total red and processed meat, g/day (includes red meat, processed meat, and offal)** | | | | | | | |
| ***Cohorts with large proportions of vegetarians*** | | | | | | | | |
| Adventist Health Study-2 | 23.6 (27.2) | - | - | - | - | - | - | - |
| CARRS-1 | 44.0 (38.9) | - | - | - | - | - | - | - |
| CARRS-2 | 44.9 (71.4) | - | - | - | - | - | - | - |
| EPIC-Oxford | 49.7 (34.6) | - | - | - | - | - | - | - |
| Oxford Vegetarian Study[3] | 76.2 (39.3) | - | - | - | - | - | - | - |
| Tzu Chi Health Study | 29.5 (41.3) | - | - | - | - | - | - | - |
| UK Women's Cohort Study | 70.8 (45.2) | - | - | - | - | - | - | - |
| ***Very large cohorts*** | | | | | | | | |
| China Kadoorie Biobank[4] | 54.6 (14.2) | - | - | - | - | - | - | - |
| Million Women Study | 37.9 (27.5) | - | - | - | - | - | - | - |
| NIH-AARP | 68.0 (50.7) | - | - | - | - | - | - | - |
| UK Biobank[4] | 58.9 (14.1) | - | - | - | - | - | - | - |
| | **Red meat, g/day** | | | | | | | |
| ***Cohorts with large proportions of vegetarians*** | | | | | | | | |
| Adventist Health Study-2 | 20.6 (24.2) | - | - | - | - | - | - | - |
| CARRS-1 | 38.1 (33.7) | - | - | - | - | - | - | - |
| CARRS-2 | 38.9 (64.8) | - | - | - | - | - | - | - |
| EPIC-Oxford | 27.7 (24.7) | - | - | - | - | - | - | - |
| Oxford Vegetarian Study[3] | 69.1 (38.0) | - | - | - | - | - | - | - |
| Tzu Chi Health Study | 26.1 (38.4) | - | - | - | - | - | - | - |
| UK Women's Cohort Study | 48.1 (36.6) | - | - | - | - | - | - | - |
| ***Very large cohorts*** | | | | | | | | |
| China Kadoorie Biobank[4] | NA | - | - | - | - | - | - | - |
| Million Women Study | 26.8 (23.2) | - | - | - | - | - | - | - |
| NIH-AARP | 50.6 (39.7) | - | - | - | - | - | - | - |
| UK Biobank[4] | 40.2 (11.5) | - | - | - | - | - | - | - |

| | | | | | | | | |
|---|---|---|---|---|---|---|---|---|
| | **Processed meat, g/day** | | | | | | | |
| ***Cohorts with large proportions of vegetarians*** | | | | | | | | |
| Adventist Health Study-2 | 3.0 (5.4) | - | - | - | - | - | - | - |
| CARRS-1 | NA | - | - | - | - | - | - | - |
| CARRS-2 | NA | - | - | - | - | - | - | - |
| EPIC-Oxford | 20.0 (16.4) | - | - | - | - | - | - | - |
| Oxford Vegetarian Study[3] | NA | - | - | - | - | - | - | - |
| Tzu Chi Health Study | 2.3 (5.5) | - | - | - | - | - | - | - |
| UK Women's Cohort Study | 18.9 (14.2) | - | - | - | - | - | - | - |
| ***Very large cohorts*** | | | | | | | | |
| China Kadoorie Biobank[4] | NA | - | - | - | - | - | - | - |
| Million Women Study | 10.0 (10.7) | - | - | - | - | - | - | - |
| NIH-AARP | 16.2 (18.4) | - | - | - | - | - | - | - |
| UK Biobank[4] | 18.6 (6.4) | - | - | - | - | - | - | - |
| | **Offal, g/day** | | | | | | | |
| ***Cohorts with large proportions of vegetarians*** | | | | | | | | |
| Adventist Health Study-2 | NA | - | - | - | - | - | - | - |
| CARRS-1 | 5.9 (13.6) | - | - | - | - | - | - | - |
| CARRS-2 | 5.9 (20.1) | - | - | - | - | - | - | - |
| EPIC-Oxford | 2.0 (3.3) | - | - | - | - | - | - | - |
| Oxford Vegetarian Study[3] | 7.1 (7.6) | - | - | - | - | - | - | - |
| Tzu Chi Health Study | 1.1 (4.1) | - | - | - | - | - | - | - |
| UK Women's Cohort Study | 3.9 (4.5) | - | - | - | - | - | - | - |
| ***Very large cohorts*** | | | | | | | | |
| China Kadoorie Biobank[4] | NA | - | - | - | - | - | - | - |
| Million Women Study | 1.1 (4.4) | - | - | - | - | - | - | - |
| NIH-AARP | 1.2 (3.2) | - | - | - | - | - | - | - |
| UK Biobank[4] | NA | - | - | - | - | - | - | - |
| | **Poultry, g/day** | | | | | | | |
| ***Cohorts with large proportions of vegetarians*** | | | | | | | | |
| Adventist Health Study-2 | 20.6 (22.7) | 16.6 (23.0) | - | - | - | - | - | - |
| CARRS-1 | 45.8 (47.1) | 46.2 (33.3) | - | - | - | - | - | - |

| | | | | | | | | |
|---|---|---|---|---|---|---|---|---|
| CARRS-2 | 78.5 (114.0) | 69.9 (104.7) | - | - | - | - | - | - |
| EPIC-Oxford | 26.7 (22.0) | 22.3 (24.2) | - | - | - | - | - | - |
| Oxford Vegetarian Study[3] | NA | NA | - | - | - | - | - | - |
| Tzu Chi Health Study | 13.2 (21.6) | 6.6 (11.8) | - | - | - | - | - | - |
| UK Women's Cohort Study | 23.1 (18.5) | 19.0 (25.6) | - | - | - | - | - | - |
| ***Very large cohorts*** | | | | | | | | |
| China Kadoorie Biobank[4] | 14.5 (6.8) | 17.0 (5.4) | - | - | - | - | - | - |
| Million Women Study | 21.1 (16.1) | 13.8 (20.6) | - | - | - | - | - | - |
| NIH-AARP | 39.6 (39.4) | 38.0 (48.4) | - | - | - | - | - | - |
| UK Biobank[4] | 32.0 (9.4) | 27.6 (11.8) | - | - | - | - | - | - |
| | **Total fish, g/day (includes oily fish, non-oily fish, and shellfish if available)** | | | | | | | |
| ***Cohorts with large proportions of vegetarians*** | | | | | | | | |
| Adventist Health Study-2 | 16.9 (21.0) | 16.6 (23.3) | 19.0 (24.6) | - | - | - | - | - |
| CARRS-1 | 33.6 (39.8) | 30.2 (29.6) | 41.0 (42.3) | - | - | - | - | - |
| CARRS-2 | 45.0 (71.1) | 30.1 (59.7) | 56.3 (113.9) | - | - | - | - | - |
| EPIC-Oxford | 41.8 (28.8) | 49.1 (35.0) | 39.0 (33.6) | - | - | - | - | - |
| Oxford Vegetarian Study[3] | 28.4 (20.3) | - | 21.7 (21.9) | - | - | - | - | - |
| Tzu Chi Health Study | 31.8 (41.3) | 24.2 (40.4) | 25.3 (57.6) | - | - | - | - | - |
| UK Women's Cohort Study | 32.5 (22.5) | 39.7 (30.9) | 32.4 (28.6) | - | - | - | - | - |
| ***Very large cohorts*** | | | | | | | | |
| China Kadoorie Biobank[4] | 23.6 (19.7) | 23.2 (22.1) | 33.7 (22.2) | - | - | - | - | - |
| Million Women Study | 33.7 (22.6) | 41.4 (29.7) | 43.8 (29.3) | - | - | - | - | - |
| NIH-AARP | 19.4 (20.7) | 19.0 (25.8) | 23.4 (34.9) | - | - | - | - | - |
| UK Biobank[4] | 23.1 (8.7) | 27.6 (10.5) | 27.5 (10.4) | - | - | - | - | - |
| | **Oily fish, g/day** | | | | | | | |
| ***Cohorts with large proportions of vegetarians*** | | | | | | | | |
| Adventist Health Study-2 | 5.0 (8.8) | 5.6 (10.1) | 7.0 (11.8) | - | - | - | - | - |
| CARRS-1 | NA | NA | NA | - | - | - | - | - |
| CARRS-2 | NA | NA | NA | - | - | - | - | - |
| EPIC-Oxford | 14.8 (17.1) | 19.7 (20.2) | 15.9 (19.5) | - | - | - | - | - |
| Oxford Vegetarian Study[3] | NA | - | NA | - | - | - | - | - |
| Tzu Chi Health Study | NA | NA | NA | - | - | - | - | - |

| | | | | | | | | |
|---|---|---|---|---|---|---|---|---|
| UK Women's Cohort Study | 10.1 (11.3) | 14.2 (16.6) | 12.2 (17.1) | - | - | - | - | - |
| ***Very large cohorts*** | | | | | | | | |
| China Kadoorie Biobank[4] | NA | NA | NA | - | - | - | - | - |
| Million Women Study | 18.1 (18.5) | 25.4 (23.9) | 26.7 (23.2) | - | - | - | - | - |
| NIH-AARP | 5.6 (8.9) | 5.6 (11.5) | 6.0 (12.7) | - | - | - | - | - |
| UK Biobank[4] | 11.2 (6.5) | 14.4 (7.6) | 14.2 (7.4) | - | - | - | - | - |
| | **Non-oily fish, g/day** | | | | | | | |
| ***Cohorts with large proportions of vegetarians*** | | | | | | | | |
| Adventist Health Study-2 | 12.0 (16.3) | 11.0 (17.9) | 12.1 (18.5) | - | - | - | - | - |
| CARRS-1 | NA | NA | NA | - | - | - | - | - |
| CARRS-2 | 37.1 (61.1) | 24.8 (42.6) | 49.6 (111.3) | - | - | - | - | - |
| EPIC-Oxford | 18.6 (15.0) | 20.9 (18.6) | 14.7 (15.3) | - | - | - | - | - |
| Oxford Vegetarian Study[3] | NA | - | NA | - | - | - | - | - |
| Tzu Chi Health Study | NA | NA | NA | - | - | - | - | - |
| UK Women's Cohort Study | 17.3 (14.5) | 19.8 (19.1) | 14.8 (14.9) | - | - | - | - | - |
| ***Very large cohorts*** | | | | | | | | |
| China Kadoorie Biobank[4] | NA | NA | NA | - | - | - | - | - |
| Million Women Study | 13.5 (13.0) | 13.5 (13.9) | 14.1 (14.3) | - | - | - | - | - |
| NIH-AARP | 13.8 (17.7) | 13.4 (21.6) | 17.4 (30.2) | - | - | - | - | - |
| UK Biobank[4] | 12.0 (4.2) | 13.2 (5.0) | 13.3 (4.9) | - | - | - | - | - |
| | **Total dairy, g/day** | | | | | | | |
| ***Cohorts with large proportions of vegetarians*** | | | | | | | | |
| Adventist Health Study-2 | 186.5 (218.0) | 128.2 (185.3) | 96.0 (167.0) | 133.2 (193.9) | 54.5 (133.9) | - | 104.6 (178.8) | - |
| CARRS-1 | 124.2 (141.8) | 132.3 (145.8) | 128.0 (146.3) | 227.4 (130.2) | 229.9 (128.4) | - | 217.3 (135.2) | - |
| CARRS-2 | 109.9 (148.1) | 107.1 (138.7) | 112.1 (148.3) | 235.6 (217.4) | 233.1 (185.4) | - | 229.0 (190.8) | - |
| EPIC-Oxford | 439.8 (215.4) | 404.9 (240.7) | 396.0 (230.5) | 396.9 (235.3) | 324.4 (244.4) | - | 376.0 (240.8) | - |
| Oxford Vegetarian Study[3] | 170.7 (104.6) | - | 162.9 (102.4) | 165.5 (107.6) | 130.9 (114.3) | - | 160.0 (109.6) | - |
| Tzu Chi Health Study | 50.5 (79.4) | 43.2 (59.7) | 45.2 (68.8) | 46.7 (69.3) | 34.7 (59.2) | - | 42.3 (66.8) | - |
| UK Women's Cohort Study | 424.3 (202.5) | 385.8 (224.0) | 384.3 (220.8) | 385.2 (219.2) | 298.9 (239.8) | - | 366.4 (227.0) | - |
| ***Very large cohorts*** | | | | | | - | | - |
| China Kadoorie Biobank[4] | 46.4 (37.6) | 58.1 (47.2) | 62.6 (48.5) | 91.3 (40.6) | 92.0 (40.3) | - | 38.9 (32.3) | - |
| Million Women Study | 377.4 (194.8) | 344.5 (207.2) | 357.2 (212.0) | 355.9 (210.8) | 287.9 (220.8) | - | 345.9 (214.0) | - |

| | | | | | | | | |
|---|---|---|---|---|---|---|---|---|
| NIH-AARP | 341.9 (316.5) | 300.1 (308.8) | 269.7 (300.2) | 320.1 (395.8) | 287.6 (384.5) | - | 302.7 (390.2) | - |
| UK Biobank[4] | NA | NA | NA | NA | NA | - | NA | - |
| | **Dairy milk, g/day** | | | | | | | |
| ***Cohorts with large proportions of vegetarians*** | | | | | | | | |
| Adventist Health Study-2 | 137.4 (200.3) | 83.1 (161.3) | 58.0 (144.2) | 87.6 (173.2) | 29.7 (114.7) | - | 66.9 (157.0) | - |
| CARRS-1 | NA | NA | NA | NA | NA | - | NA | - |
| CARRS-2 | 66.1 (118.7) | 60.7 (112.3) | 63.7 (111.9) | 147.7 (186.4) | 146.4 (152.9) | - | 143.8 (157.5) | - |
| EPIC-Oxford | 325.7 (186.7) | 275.7 (195.2) | 269.1 (191.4) | 274.1 (201.2) | 212.2 (202.4) | - | 256.6 (203.7) | - |
| Oxford Vegetarian Study[3] | 137.4 (102.3) | - | 114.3 (96.0) | 117.9 (102.4) | 90.9 (106.3) | - | 113.8 (103.2) | - |
| Tzu Chi Health Study | 41.0 (72.6) | 35.6 (53.4) | 35.4 (63.2) | 36.2 (57.9) | 28.2 (55.7) | - | 33.0 (56.5) | - |
| UK Women's Cohort Study | 316.5 (184.4) | 267.6 (199.7) | 266.9 (195.9) | 268.0 (195.6) | 198.3 (204.5) | - | 253.1 (199.6) | - |
| ***Very large cohorts*** | | | | | | - | | - |
| China Kadoorie Biobank[4] | NA | NA | NA | NA | NA | - | NA | - |
| Million Women Study | 268.9 (176.1) | 232.8 (179.4) | 236.1 (186.2) | 229.9 (184.9) | 188.3 (190.7) | - | 223.8 (186.3) | - |
| NIH-AARP | 242.1 (302.2) | 213.6 (291.5) | 166.1 (271.7) | 207.2 (368.4) | 185.6 (353.5) | - | 195.6 (360.6) | - |
| UK Biobank[4] | NA | NA | NA | NA | NA | - | NA | - |
| | **Cheese, g/day** | | | | | | | |
| ***Cohorts with large proportions of vegetarians*** | | | | | | | | |
| Adventist Health Study-2 | 19.3 (25.7) | 14.8 (23.9) | 12.2 (21.3) | 19.9 (26.3) | 8.0 (17.4) | - | 15.6 (24.2) | - |
| CARRS-1 | NA | NA | NA | NA | NA | - | NA | - |
| CARRS-2 | NA | NA | NA | NA | NA | - | NA | - |
| EPIC-Oxford | 20.5 (18.3) | 22.3 (21.6) | 27.3 (23.9) | 31.4 (24.7) | 27.1 (26.1) | - | 30.1 (25.2) | - |
| Oxford Vegetarian Study[3] | 33.3 (21.5) | - | 48.6 (27.5) | 47.6 (26.1) | 40.0 (31.2) | - | 46.2 (27.1) | - |
| Tzu Chi Health Study | 0.7 (2.4) | 0.5 (2.2) | 0.8 (2.5) | 1.2 (3.8) | 1.2 (3.5) | - | 1.1 (3.6) | - |
| UK Women's Cohort Study | 22.9 (21.9) | 30.3 (27.7) | 32.8 (28.6) | 36.4 (30.8) | 29.2 (31.5) | - | 34.8 (31.1) | - |
| ***Very large cohorts*** | | | | | | - | | - |
| China Kadoorie Biobank[4] | NA | NA | NA | NA | NA | - | NA | - |
| Million Women Study | 19.5 (16.3) | 20.1 (18.1) | 26.6 (19.1) | 32.5 (20.8) | 22.4 (23.9) | - | 31.1 (21.5) | - |
| NIH-AARP | 8.7 (9.6) | 4.6 (7.7) | 8.2 (13.1) | 10.9 (15.6) | 7.2 (11.1) | - | 9.1 (13.7) | - |
| UK Biobank[4] | 16.4 (6.5) | 14.4 (7.5) | 18.3 (6.2) | 19.2 (5.2) | 14.9 (6.6) | - | 18.3 (6.1) | - |

| | | | | | | | | |
|---|---|---|---|---|---|---|---|---|
| | Yogurt, g/day | | | | | | | |
| ***Cohorts with large proportions of vegetarians*** | | | | | | | | |
| Adventist Health Study-2 | 29.7 (58.7) | 30.3 (61.5) | 25.9 (57.1) | 25.7 (51.2) | 16.9 (45.9) | - | 22.1 (49.1) | - |
| CARRS-1 | NA | NA | NA | NA | NA | - | NA | - |
| CARRS-2 | 43.9 (74.1) | 46.4 (69.5) | 48.4 (73.6) | 87.9 (94.4) | 86.6 (89.7) | - | 85.2 (90.3) | - |
| EPIC-Oxford | 34.3 (40.0) | 41.1 (46.8) | 37.7 (42.5) | 34.6 (40.2) | 32.6 (44.2) | - | 33.8 (41.3) | - |
| Oxford Vegetarian Study[3] | NA | - | NA | NA | NA | - | NA | - |
| Tzu Chi Health Study | 9.2 (26.4) | 6.5 (16.4) | 9.1 (27.3) | 9.4 (30.8) | 8.5 (45.3) | - | 8.7 (32.5) | - |
| UK Women's Cohort Study | 58.0 (62.4) | 67.4 (73.2) | 62.9 (68.4) | 58.5 (64.4) | 53.4 (86.9) | - | 57.1 (69.0) | - |
| ***Very large cohorts*** | | | | | | - | | - |
| China Kadoorie Biobank[4] | NA | NA | NA | NA | NA | - | NA | - |
| Million Women Study | 75.0 (68.7) | 82.9 (75.9) | 84.3 (75.4) | 83.1 (77.6) | 69.3 (84.6) | - | 81.0 (78.6) | - |
| NIH-AARP | 15.4 (38.4) | 32.9 (60.5) | 35.9 (67.2) | 32.3 (60.7) | 35.0 (68.9) | - | 33.3 (64.5) | - |
| UK Biobank[4] | NA | NA | NA | NA | NA | - | NA | - |
| | Eggs, g/day | | | | | | | |
| ***Cohorts with large proportions of vegetarians*** | | | | | | | | |
| Adventist Health Study-2 | 15.4 (20.8) | 10.7 (16.0) | 8.4 (12.7) | 11.3 (15.0) | - | 8.5 (12.0) | 7.8 (13.5) | - |
| CARRS-1 | 23.4 (13.3) | 21.9 (13.0) | 19.2 (14.0) | 18.2 (14.0) | - | 21.0 (13.9) | 4.3 (9.8) | - |
| CARRS-2 | 22.2 (31.5) | 18.4 (23.2) | 13.5 (27.3) | 18.8 (39.4) | - | 20.8 (25.2) | 3.9 (16.1) | - |
| EPIC-Oxford | 11.5 (11.1) | 9.8 (10.9) | 11.5 (11.1) | 13.8 (12.5) | - | 15.0 (13.1) | 10.6 (12.1) | - |
| Oxford Vegetarian Study[3] | 29.2 (20.8) | - | 29.4 (22.2) | 28.4 (20.9) | - | 28.6 (25.2) | 25.7 (21.6) | - |
| Tzu Chi Health Study | 20.7 (19.1) | 13.8 (12.6) | 16.4 (16.2) | 19.2 (16.4) | - | 18.9 (17.5) | 16.4 (16.7) | - |
| UK Women's Cohort Study | 18.7 (15.5) | 15.3 (14.2) | 17.5 (15.6) | 17.8 (14.9) | - | 24.2 (41.1) | 14.7 (15.4) | - |
| ***Very large cohorts*** | | | | | - | | | - |
| China Kadoorie Biobank[4] | 33.3 (9.1) | 35.0 (11.4) | 36.5 (11.6) | 36.4 (10.3) | - | 33.1 (8.9) | 33.0 (9.5) | - |
| Million Women Study | 17.4 (12.9) | 14.4 (12.9) | 17.3 (14.0) | 18.1 (14.4) | - | 18.4 (20.8) | 15.9 (14.8) | - |
| NIH-AARP | 11.1 (13.5) | 5.1 (10.0) | 7.5 (13.2) | 9.8 (13.7) | - | 10.2 (26.1) | 5.3 (11.3) | - |
| UK Biobank[4] | NA | NA | NA | NA | NA | - | NA | - |

Abbreviations: CARRS, Centre for cArdiometabolic Risk Reduction in South Asia; CKB, EPIC, European Prospective Investigation into Cancer and Nutrition; NIH-AARP, National Institutes of Health-AARP Diet and Health Study.

[1]All values are mean (SD).

[2]All vegetarians include lacto-ovo, lacto and ovo vegetarians.

[3]Poultry eaters could not be determined as poultry intake was not assessed.

[4]Mean intakes assessed at resurvey have been assigned to baseline categories based on quintiles of intakes to estimate the baseline intakes.

NA indicates that no information was available; – indicates that this food was not applicable for the corresponding diet group.

**Table S8.** Baseline nutrient intakes by diet group and cohort (where nutrient data were available)[1].

| Cohort | Meat eaters | Poultry eaters | Pescatarians | Vegetarians | | | | Vegans |
|---|---|---|---|---|---|---|---|---|
| | | | | Lacto-ovo vegetarians | Lacto vegetarians | Ovo vegetarians | All vegetarians[2] | |
| | **Energy, kcal/day** | | | | | | | |
| ***Cohorts with large proportions of vegetarians*** | | | | | | | | |
| Adventist Health Study-2 | 1,799 (681) | 1,720 (653) | 1,790 (668) | 1,797 (635) | 1,745 (636) | 1,711 (653) | 1,778 (637) | 1,760 (646) |
| EPIC-Oxford | 1,992 (536) | 1,825 (530) | 1,896 (528) | 1,923 (529) | 1,769 (515) | 1,586 (459) | 1,881 (530) | 1,752 (555) |
| UK Women's Cohort Study | 2,226 (549) | 2,160 (561) | 2,228 (563) | 2,223 (555) | 2,048 (607) | 2,195 (639) | 2,191 (569) | 2,100 (613) |
| ***Very large cohorts*** | | | | | | | | |
| Million Women Study | 1,663 (420) | 1,492 (410) | 1,585 (412) | 1,567 (413) | 1,387 (442) | 1,394 (420) | 1,544 (421) | 1,178 (308) |
| NIH-AARP | 1,822 (669) | 1,370 (533) | 1,555 (629) | 1,629 (622) | 1,619 (618) | 1,446 (751) | 1,623 (621) | 1,604 (637) |
| | **Protein, g/day** | | | | | | | |
| ***Cohorts with large proportions of vegetarians*** | | | | | | | | |
| Adventist Health Study-2 | 68.9 (28.4) | 64.8 (27.4) | 65.9 (28.3) | 63.8 (25.7) | 62.1 (26.4) | 57.8 (24.5) | 63.1 (25.9) | 62.0 (26.1) |
| EPIC-Oxford | 82.3 (21.6) | 72.5 (20.6) | 67.7 (19.2) | 64.1 (17.7) | 58.2 (17.8) | 50.6 (14.6) | 62.5 (17.9) | 56.7 (19.0) |
| UK Women's Cohort Study | 89.9 (23.3) | 80.1 (22.1) | 77.3 (21.1) | 73.6 (19.6) | 65.8 (20.8) | 68.9 (20.5) | 72.1 (20.0) | 65.1 (21.2) |
| ***Very large cohorts*** | | | | | | | | |
| Million Women Study | 67.0 (17.2) | 56.0 (16.0) | 57.4 (15.0) | 50.8 (13.8) | 42.1 (14.3) | 40.3 (12.0) | 49.7 (14.2) | 33.7 (10.8) |
| NIH-AARP | 70.0 (28.4) | 51.0 (23.2) | 50.9 (23.6) | 49.4 (22.4) | 48.6 (22.0) | 41.9 (25.3) | 48.9 (22.3) | 45.8 (20.3) |
| | **Carbohydrates, g/day** | | | | | | | |
| ***Cohorts with large proportions of vegetarians*** | | | | | | | | |
| Adventist Health Study-2 | 238 (101) | 243 (102) | 258 (105) | 251 (95) | 260 (102) | 256 (103) | 254 (97) | 274 (106) |
| EPIC-Oxford | 248 (74) | 244 (76) | 252 (74) | 260 (74) | 249 (75) | 231 (74) | 257 (74) | 252 (79) |
| UK Women's Cohort Study | 290 (81) | 303 (85) | 306 (85) | 309 (86) | 296 (93) | 316 (87) | 307 (87) | 308 (96) |
| ***Very large cohorts*** | | | | | | | | |
| Million Women Study | 207 (58) | 201 (60) | 208 (58) | 216 (60) | 203 (63) | 214 (73) | 215 (61) | 178 (52) |
| NIH-AARP | 233 (91) | 217 (93) | 249 (105) | 258 (99) | 271 (108) | 260 (150) | 264 (104) | 293 (120) |
| | **Total fat, g/day** | | | | | | | |
| ***Cohorts with large proportions of vegetarians*** | | | | | | | | |

| | | | | | | | | |
|---|---|---|---|---|---|---|---|---|
| Adventist Health Study-2 | 69.2 (32.1) | 61.3 (29.5) | 62.9 (30.6) | 67.5 (29.5) | 59.1 (27.3) | 59.3 (32.2) | 64.7 (29.2) | 56.0 (28.1) |
| EPIC-Oxford | 72.4 (26.4) | 61.7 (25.3) | 66.8 (25.7) | 68.1 (26.0) | 59.8 (25.5) | 51.8 (20.8) | 65.9 (26.1) | 56.8 (26.9) |
| UK Women's Cohort Study | 80.2 (25.7) | 73.2 (25.9) | 79.4 (26.7) | 80.0 (26.4) | 71.1 (28.4) | 78.1 (31.7) | 78.4 (27.0) | 70.9 (29.4) |
| ***Very large cohorts*** | | | | | | | | |
| Million Women Study | 63.2 (21.7) | 52.5 (20.5) | 58.2 (21.5) | 56.4 (21.1) | 47.2 (21.8) | 44.0 (19.8) | 55.2 (21.4) | 37.3 (15.3) |
| NIH-AARP | 63.0 (29.5) | 33.8 (17.6) | 41.2 (23.7) | 48.0 (26.5) | 42.0 (23.1) | 33.0 (16.3) | 45.2 (25.1) | 33.4 (21.9) |
| | **Saturated fat, g/day** | | | | | | | |
| ***Cohorts with large proportions of vegetarians*** | | | | | | | | |
| Adventist Health Study-2 | 19.4 (10.0) | 15.0 (7.9) | 14.3 (7.9) | 16.3 (8.0) | 12.5 (6.3) | 10.7 (5.4) | 15.0 (7.7) | 9.8 (4.9) |
| EPIC-Oxford | 26.7 (11.6) | 20.9 (10.4) | 23.1 (10.9) | 24.1 (10.9) | 20.5 (10.7) | 12.1 (5.4) | 23.1 (11.0) | 13.2 (6.9) |
| UK Women's Cohort Study | 28.4 (11.0) | 23.6 (10.3) | 25.8 (10.7) | 26.6 (10.9) | 21.8 (11.5) | 16.2 (7.3) | 25.7 (11.2) | 13.4 (6.2) |
| ***Very large cohorts*** | | | | | | | | |
| Million Women Study | 22.1 (9.9) | 17.6 (8.9) | 20.0 (9.4) | 20.8 (9.5) | 16.5 (9.9) | 10.0 (4.0) | 20.2 (9.7) | 8.2 (3.8) |
| NIH-AARP | 19.8 (10.2) | 9.7 (5.6) | 11.8 (7.7) | 14.3 (9.0) | 12.2 (8.0) | 7.3 (3.8) | 13.3 (8.6) | 7.2 (4.2) |
| | **Monounsaturated fat, g/day** | | | | | | | |
| ***Cohorts with large proportions of vegetarians*** | | | | | | | | |
| Adventist Health Study-2 | 27.8 (14.4) | 25.4 (14.2) | 26.7 (15.3) | 27.9 (13.9) | 25.1 (13.4) | 26.7 (18.4) | 27.0 (14.0) | 24.9 (14.9) |
| EPIC-Oxford | 24.5 (9.5) | 20.4 (9.0) | 21.6 (8.9) | 21.8 (9.2) | 19.0 (9.1) | 17.7 (9.0) | 21.0 (9.2) | 19.0 (10.6) |
| UK Women's Cohort Study | 26.5 (8.8) | 23.6 (9.1) | 25.7 (9.4) | 25.6 (9.2) | 22.4 (9.8) | 24.2 (10.2) | 25.0 (9.4) | 22.2 (10.8) |
| ***Very large cohorts*** | | | | | | | | |
| Million Women Study | 23.1 (8.4) | 19.2 (8.2) | 21.2 (8.5) | 19.9 (8.3) | 17.1 (8.6) | 18.1 (10.0) | 19.5 (8.4) | 15.9 (7.4) |
| NIH-AARP | 23.8 (11.5) | 12.2 (6.8) | 15.1 (9.2) | 17.9 (10.5) | 15.5 (9.0) | 12.4 (6.8) | 16.7 (9.9) | 12.6 (9.5) |
| | **Polyunsaturated fat, g/day** | | | | | | | |
| ***Cohorts with large proportions of vegetarians*** | | | | | | | | |
| Adventist Health Study-2 | 16.5 (8.0) | 16.1 (8.0) | 17.2 (8.4) | 18.3 (8.3) | 17.1 (8.2) | 17.6 (8.8) | 17.9 (8.3) | 17.0 (8.5) |
| EPIC-Oxford | 14.3 (6.3) | 13.8 (6.5) | 15.0 (6.8) | 15.1 (7.0) | 13.8 (7.0) | 16.3 (6.6) | 14.8 (7.0) | 18.4 (9.2) |
| UK Women's Cohort Study | 14.8 (5.3) | 15.8 (6.5) | 16.9 (6.4) | 16.7 (6.5) | 15.9 (7.2) | 22.5 (9.2) | 16.6 (6.7) | 21.5 (9.5) |
| ***Very large cohorts*** | | | | | | | | |
| Million Women Study | 11.8 (4.7) | 10.0 (4.4) | 10.9 (4.6) | 10.0 (4.3) | 8.8 (4.6) | 12.3 (5.5) | 9.8 (4.4) | 10.1 (4.1) |
| NIH-AARP | 14.3 (7.0) | 9.0 (5.1) | 11.0 (6.6) | 12.2 (7.2) | 11.1 (6.7) | 10.3 (5.1) | 11.7 (6.9) | 10.5 (7.5) |

| | | | | | | | | |
|---|---|---|---|---|---|---|---|---|
| | **Dietary fibre, g/day** | | | | | | | |
| ***Cohorts with large proportions of vegetarians*** | | | | | | | | |
| Adventist Health Study-2 | 26.5 (13.2) | 31.1 (14.8) | 35.6 (16.5) | 33.1 (14.1) | 37.1 (15.8) | 38.5 (16.1) | 34.5 (14.8) | 43.2 (17.3) |
| EPIC-Oxford | 18.6 (6.7) | 20.7 (7.5) | 21.2 (7.6) | 21.5 (7.5) | 21.8 (8.3) | 23.7 (8.5) | 21.6 (7.7) | 26.1 (9.3) |
| UK Women's Cohort Study | 23.0 (8.3) | 27.4 (9.7) | 27.3 (9.2) | 27.3 (9.1) | 26.9 (10.1) | 32.0 (9.3) | 27.3 (9.3) | 31.6 (10.8) |
| ***Very large cohorts*** | | | | | | | | |
| Million Women Study | 13.8 (4.7) | 14.6 (5.1) | 15.7 (5.2) | 16.3 (5.4) | 15.6 (6.5) | 19.9 (6.8) | 16.3 (5.6) | 17.9 (5.6) |
| NIH-AARP | 18.9 (8.7) | 20.7 (11.0) | 26.6 (14.6) | 27.2 (13.3) | 29.4 (14.6) | 34.2 (22.9) | 28.3 (14.1) | 40.7 (17.8) |
| | **Vitamin C, mg/day** | | | | | | | |
| ***Cohorts with large proportions of vegetarians*** | | | | | | | | |
| Adventist Health Study-2 | 152 (109) | 181 (122) | 201 (133) | 170 (103) | 188 (121) | 187 (129) | 176 (110) | 206 (129) |
| EPIC-Oxford | 132 (63) | 146 (73) | 144 (69) | 142 (69) | 146 (77) | 151 (74) | 143 (72) | 163 (93) |
| UK Women's Cohort Study | 162 (74) | 178 (81) | 175 (81) | 173 (80) | 169 (89) | 189 (88) | 172 (82) | 203 (116) |
| ***Very large cohorts*** | | | | | | | | |
| Million Women Study | 96 (47) | 104 (54) | 112 (54) | 116 (57) | 114 (67) | 146 (74) | 116 (59) | 138 (59) |
| NIH-AARP | 156 (107) | 182 (130) | 203 (145) | 203 (130) | 212 (138) | 178 (149) | 207 (134) | 305 (197) |
| | **Vitamin B12, µg/day** | | | | | | | |
| ***Cohorts with large proportions of vegetarians*** | | | | | | | | |
| Adventist Health Study-2 | 4.0 (2.6) | 3.6 (3.0) | 3.6 (2.9) | 3.4 (2.7) | 2.8 (2.8) | 2.1 (2.6) | 3.2 (2.7) | 1.9 (2.6) |
| EPIC-Oxford | 7.1 (3.4) | 5.7 (2.7) | 4.9 (2.7) | 2.7 (1.3) | 2.0 (1.2) | 0.7 (0.4) | 2.5 (1.3) | 0.4 (0.6) |
| UK Women's Cohort Study | 6.5 (2.8) | 4.4 (2.1) | 4.1 (2.1) | 2.6 (1.1) | 1.9 (1.1) | 1.1 (1.1) | 2.5 (1.2) | 0.8 (0.6) |
| ***Very large cohorts*** | | | | | | | | |
| Million Women Study | 4.6 (2.3) | 3.7 (1.9) | 3.9 (1.9) | 2.0 (0.9) | 1.4 (0.9) | 0.6 (0.4) | 1.9 (0.9) | 0.2 (0.5) |
| NIH-AARP | 4.8 (3.1) | 2.8 (1.9) | 2.6 (2.0) | 2.1 (1.8) | 1.9 (1.9) | 0.7 (0.6) | 2.0 (1.8) | 0.5 (0.6) |
| | **Folate, µg/day** | | | | | | | |
| ***Cohorts with large proportions of vegetarians*** | | | | | | | | |
| Adventist Health Study-2 | 468 (229) | 514 (248) | 573 (274) | 549 (250) | 591 (291) | 591 (321) | 564 (267) | 667 (341) |
| EPIC-Oxford | 323 (100) | 329 (107) | 346 (112) | 357 (117) | 344 (128) | 383 (129) | 354 (120) | 419 (159) |
| UK Women's Cohort Study | 379 (112) | 405 (123) | 404 (122) | 405 (121) | 383 (137) | 433 (124) | 401 (125) | 453 (159) |
| ***Very large cohorts*** | | | | | | | | |

| | | | | | | | | |
|---|---|---|---|---|---|---|---|---|
| Million Women Study | 251 (70) | 245 (74) | 262 (78) | 269 (81) | 243 (94) | 289 (86) | 266 (84) | 250 (84) |
| NIH-AARP | 408 (170) | 405 (198) | 506 (259) | 508 (232) | 546 (250) | 549 (327) | 526 (242) | 705 (309) |
| | **Vitamin D, µg/day** | | | | | | | |
| ***Cohorts with large proportions of vegetarians*** | | | | | | | | |
| Adventist Health Study-2 | 4.0 (3.2) | 3.2 (3.2) | 2.9 (2.9) | 2.2 (2.7) | 1.4 (2.5) | 0.7 (1.3) | 1.9 (2.6) | 0.6 (1.9) |
| EPIC-Oxford | 3.3 (1.9) | 3.1 (2.0) | 2.8 (2.0) | 1.6 (1.2) | 1.2 (1.1) | 1.2 (1.1) | 1.5 (1.2) | 0.9 (1.0) |
| UK Women's Cohort Study | 3.2 (1.5) | 3.1 (1.9) | 3.0 (1.9) | 1.9 (1.1) | 1.4 (0.9) | 2.1 (1.6) | 1.8 (1.1) | 1.4 (1.2) |
| ***Very large cohorts*** | | | | | | | | |
| Million Women Study | 3.1 (1.7) | 3.1 (2.0) | 3.1 (2.0) | 1.4 (1.0) | 1.0 (1.1) | 1.0 (0.6) | 1.4 (1.0) | 0.6 (0.5) |
| NIH-AARP | 4.6 (3.1) | 3.6 (3.0) | 3.3 (3.1) | 2.8 (3.5) | 2.5 (3.3) | 0.6 (0.5) | 2.6 (3.4) | 0.5 (0.5) |
| | **Calcium, mg/day** | | | | | | | |
| ***Cohorts with large proportions of vegetarians*** | | | | | | | | |
| Adventist Health Study-2 | 776 (401) | 734 (385) | 730 (365) | 752 (361) | 675 (340) | 552 (239) | 722 (354) | 609 (273) |
| EPIC-Oxford | 1,007 (312) | 969 (348) | 1,031 (348) | 1,069 (363) | 932 (364) | 523 (190) | 1,031 (369) | 590 (237) |
| UK Women's Cohort Study | 1,095 (328) | 1,109 (354) | 1,149 (372) | 1,153 (357) | 997 (379) | 712 (253) | 1,121 (367) | 686 (240) |
| ***Very large cohorts*** | | | | | | | | |
| Million Women Study | 836 (284) | 799 (304) | 876 (315) | 893 (316) | 723 (331) | 378 (136) | 870 (324) | 328 (155) |
| NIH-AARP | 767 (420) | 700 (409) | 745 (425) | 801 (507) | 776 (507) | 423 (291) | 786 (507) | 551 (252) |
| | **Iron, mg/day** | | | | | | | |
| ***Cohorts with large proportions of vegetarians*** | | | | | | | | |
| Adventist Health Study-2 | 16.1 (7.7) | 16.9 (8.0) | 18.4 (8.6) | 18.0 (7.9) | 19.0 (8.6) | 18.4 (8.7) | 18.3 (8.1) | 20.1 (9.0) |
| EPIC-Oxford | 12.4 (3.5) | 12.2 (3.7) | 12.6 (3.8) | 12.7 (3.8) | 12.2 (4.1) | 13.1 (4.0) | 12.6 (3.9) | 14.3 (4.6) |
| UK Women's Cohort Study | 17.2 (6.5) | 18.0 (7.0) | 17.8 (6.6) | 17.6 (6.4) | 16.7 (7.0) | 18.8 (5.5) | 17.4 (6.5) | 19.6 (6.5) |
| ***Very large cohorts*** | | | | | | | | |
| Million Women Study | - | - | - | - | - | - | - | - |
| NIH-AARP | 14.9 (6.0) | 13.3 (6.3) | 15.1 (7.1) | 15.3 (6.8) | 16.4 (7.6) | 17.1 (13.1) | 15.8 (7.3) | 19.2 (8.7) |

Abbreviations: EPIC, European Prospective Investigation into Cancer and Nutrition; NIH-AARP, National Institutes of Health-AARP Diet and Health Study.
[1]All values are mean (SD).
"-" indicates that no information was available.

**Table S9.** Number and percentage of participants in each diet group at resurvey categorized by their baseline diet group (n = 104,944)[1].

| | | CARRS-1 | EPIC-Oxford | Oxford Vegetarian Study[2] | UK Women's Cohort Study | China Kadoorie Biobank | Million Women Study[3] | UK Biobank |
|---|---|---|---|---|---|---|---|---|
| N with resurvey dietary data (% of baseline) | | 7,389 (60.5) | 27,529 (51.2) | 2,837 (26.9) | 1,652 (5.5) | 24,586 (4.8) | 23,525 (3.7) | 17,426 (3.7) |
| Median (IQR) time between baseline and resurvey (years) | | 5.9 (5.4–6.2) | 14.3 (13.4–15.2) | 13.6 (12.3–14.0) | 4.8 (4.6–5.1) | 8.0 (7.4–8.6) | 13.0 (11.1–14.2) | 4.4 (3.7–5.0) |
| **Baseline diet group** | **Resurvey diet group** | **N (%) in the same diet group as at baseline** | | | | | | |
| **Meat eater** | **Meat eater** | **3,072 (71.7)** | **12,730 (96.3)** | **1,075 (87.2)** | **728 (96.4)** | **21,286 (90.4)** | **19,975 (90.8)** | **17,426 (99.2)** |
| | Poultry eater | 969 (22.6) | 137 (1.0) | 28 (2.3) | 11 (1.5) | 904 (3.8) | 1,265 (5.8) | 83 (0.47) |
| | Pescatarian | 71 (1.7) | 251 (1.9) | 82 (6.7) | 11 (1.5) | 314 (1.3) | 626 (2.8) | 51 (0.29) |
| | Vegetarian[5] | 164 (3.8) | 100 (0.76) | 46 (3.7) | 5 (0.66) | 817 (3.5) | 129 (0.59) | 13 (0.07) |
| | Vegan | 9 (0.21) | 6 (0.05) | 2 (0.2) | 0 (0.00) | 229 (0.97) | 2 (0.01) | 2 (0.01) |
| **Poultry eater** | Meat eater | 630 (51.5) | 644 (74.3) | - | 30 (35.3) | 64 (78.0) | 54 (38.3) | 67 (38.3) |
| | **Poultry eater** | **517 (42.2)** | **114 (13.1)** | - | **46 (54.1)** | **10 (12.2)** | **58 (41.1)** | **97 (55.4)** |
| | Pescatarian | 19 (1.6) | 89 (10.3) | - | 8 (9.4) | 4 (4.9) | 19 (13.5) | 8 (4.6) |
| | Vegetarian[5] | 51 (4.2) | 19 (2.2) | - | 1 (1.2) | 4 (4.9) | 10 (7.1) | 3 (1.7) |
| | Vegan | 7 (0.57) | 1 (0.12) | - | 0 (0.00) | 0 (0.00) | 0 (0.00) | 0 (0.00) |
| **Pescatarian** | Meat eater | 46 (45.5) | 1,394 (31.4) | 66 (21.6) | 35 (10.5) | 27 (45.8) | 139 (16.6) | 44 (7.3) |
| | Poultry eater | 20 (19.8) | 151 (3.4) | 14 (4.6) | 5 (1.5) | 0 (0.00) | 53 (6.3) | 14 (2.3) |
| | **Pescatarian** | **23 (22.8)** | **2,506 (56.4)** | **145 (47.4)** | **276 (82.6)** | **25 (42.4)** | **525 (62.6)** | **522 (87.1)** |
| | Vegetarian[5] | 10 (9.9) | 378 (8.5) | 77 (25.2) | 16 (4.8) | 5 (8.5) | 122 (14.5) | 18 (3.0) |
| | Vegan | 2 (2.0) | 16 (0.36) | 4 (1.3) | 2 (0.60) | 2 (3.4) | 0 (0.00) | 1 (0.17) |
| **Vegetarian**[5] | Meat eater | 98 (6.5) | 991 (12.2) | 72 (5.9) | 14 (3.1) | 427 (68.5) | 22 (4.1) | 4 (1.0) |
| | Poultry eater | 81 (5.3) | 85 (1.0) | 15 (1.2) | 3 (0.66) | 34 (5.5) | 10 (1.9) | 4 (1.0) |
| | Pescatarian | 7 (0.46) | 1,252 (15.4) | 125 (10.3) | 49 (10.8) | 5 (0.80) | 82 (15.2) | 27 (6.9) |
| | **Vegetarian**[5] | **1,303 (86.0)** | **5,557 (68.3)** | **950 (78.2)** | **353 (77.9)** | **119 (19.1)** | **420 (77.8)** | **350 (89.3)** |
| | Vegan | 26 (1.7) | 247 (3.0) | 53 (4.4) | 34 (7.5) | 38 (6.1) | 6 (1.1) | 7 (1.8) |
| **Vegan** | Meat eater | 17 (6.4) | 79 (9.2) | 2 (2.4) | 2 (8.0) | 178 (65.4) | 0 (0.00) | 0 (0.00) |
| | Poultry eater | 17 (6.4) | 8 (0.93) | 0 (0.0) | 0 (0.00) | 15 (5.5) | 0 (0.00) | 0 (0.00) |
| | Pescatarian | 2 (0.76) | 73 (8.5) | 3 (3.6) | 1 (4.0) | 0 (0.00) | 0 (0.00) | 0 (0.00) |
| | Vegetarian[5] | 210 (79.5) | 224 (26.0) | 17 (20.5) | 6 (24.0) | 46 (16.9) | 6 (75.0) | 7 (18.4) |
| | **Vegan** | **18 (6.8)** | **477 (55.4)** | **61 (73.5)** | **16 (64.0)** | **33 (12.1)** | **2 (25.0)** | **31 (81.6)** |

[1]Values are N in baseline diet group or N (% of baseline diet group).

[2]There are 2,837 participants in the Oxford Vegetarian Study who also took part in EPIC-Oxford, therefore the baseline survey in EPIC-Oxford is considered as a resurvey for this subsample of Oxford Vegetarian Study participants. In the Oxford vegetarian Study, poultry eaters at baseline could not be determined as poultry intake was not assessed.

[3]In the Million Women Study, diet groups at baseline ("baseline" in this study is taken as the date of completion of the questionnaire on diet, which was ~3 years after the recruitment questionnaire) were determined using a food frequency questionnaire while diet groups at resurvey were determined using a minimum of three 24-hour dietary assessments.

[4]All vegetarians include lacto-ovo, lacto, and ovo vegetarians.

**Table S10.** Number of participants of each ethnicity by diet group and cohort[1].

| Ethnicity | Meat eaters | Poultry eaters | Pescatarians | Vegetarians | | | | Vegans |
|---|---|---|---|---|---|---|---|---|
| | | | | Lacto-ovo vegetarians | Lacto vegetarians | Ovo vegetarians | All vegetarians[2] | |
| **Adventist Health Study-2** | | | | | | | | |
| White | 14,421 (62.0) | 5,409 (51.5) | 3,070 (49.5) | 10,779 (84.2) | 4,593 (77.7) | 491 (72.6) | 15,863 (81.8) | 3,727 (71.3) |
| Black | 6,384 (27.5) | 3,978 (37.9) | 2,381 (38.4) | 1,341 (10.5) | 966 (16.3) | 126 (18.6) | 2,433 (12.5) | 1,127 (21.6) |
| Hispanic | 1,151 (5.0) | 575 (5.5) | 318 (5.1) | 323 (2.5) | 204 (3.4) | 22 (3.3) | 549 (2.8) | 173 (3.3) |
| Asian | 56 (0.24) | 38 (0.36) | 28 (0.45) | 23 (0.18) | 7 (0.12) | 1 (0.15) | 31 (0.16) | 9 (0.17) |
| Other | 1,233 (5.3) | 494 (4.7) | 405 (6.5) | 332 (2.6) | 145 (2.5) | 36 (5.3) | 513 (2.6) | 189 (3.6) |
| **European Prospective Investigation into Cancer and Nutrition-Oxford** | | | | | | | | |
| White | 25,560 (96.5) | 1,635 (96.0) | 7,799 (96.0) | 10,949 (96.1) | 3,663 (92.8) | 88 (92.6) | 14,700 (95.3) | 1,852 (93.1) |
| Black | 84 (0.32) | 13 (0.76) | 31 (0.38) | 22 (0.19) | 19 (0.48) | 1 (1.1) | 42 (0.27) | 11 (0.55) |
| Asian | 92 (0.35) | 8 (0.47) | 41 (0.50) | 64 (0.56) | 112 (2.8) | 0 (0.00) | 176 (1.1) | 6 (0.30) |
| Other | 185 (0.70) | 16 (0.94) | 104 (1.3) | 116 (1.0) | 62 (1.6) | 2 (2.1) | 180 (1.2) | 43 (2.2) |
| Not known | 577 (2.2) | 31 (1.8) | 153 (1.9) | 239 (2.1) | 92 (2.3) | 4 (4.2) | 335 (2.2) | 78 (3.9) |
| **UK Women's Cohort Study** | | | | | | | | |
| White | 20,466 (96.6) | 877 (94.4) | 3,635 (95.6) | 3,147 (95.4) | 646 (86.9) | 29 (90.6) | 3,822 (93.8) | 153 (95.6) |
| Black | 26 (0.12) | 2 (0.22) | 10 (0.26) | 1 (0.03) | 1 (0.13) | 0 (0.00) | 2 (0.05) | 1 (0.62) |
| Asian | 53 (0.25) | 5 (0.54) | 12 (0.32) | 35 (1.1) | 59 (7.9) | 1 (3.1) | 95 (2.3) | 0 (0.00) |
| Other | 99 (0.47) | 6 (0.65) | 35 (0.92) | 24 (0.73) | 13 (1.7) | 0 (0.00) | 37 (0.91) | 2 (1.2) |
| Not known | 536 (2.5) | 39 (4.2) | 112 (2.9) | 93 (2.8) | 24 (3.2) | 2 (6.2) | 119 (2.9) | 4 (2.5) |
| **Million Women Study** | | | | | | | | |
| White | 607,129 (98.2) | 3,330 (97.5) | 10,741 (98.1) | 5,206 (97.1) | 606 (81.7) | 33 (100) | 5,845 (95.3) | 76 (96.2) |
| Black | 1,084 (0.18) | 9 (0.26) | 17 (0.16) | 1 (0.02) | 1 (0.13) | 0 (0.00) | 2 (0.03) | 0 (0.00) |
| Asian | 1,531 (0.25) | 23 (0.67) | 41 (0.37) | 97 (1.8) | 122 (16.4) | 0 (0.00) | 219 (3.6) | 0 (0.00) |
| Other | 1,914 (0.31) | 18 (0.53) | 47 (0.43) | 19 (0.35) | 5 (0.67) | 0 (0.00) | 24 (0.39) | 1 (1.3) |
| Not known | 6,792 (1.1) | 34 (1.00) | 101 (0.92) | 38 (0.71) | 8 (1.1) | 0 (0.00) | 46 (0.75) | 2 (2.5) |
| **National Institutes of Health-AARP Diet and Health Study** | | | | | | | | |
| White | 452,695 (92.2) | 28,546 (86.6) | 1,457 (86.2) | 839 (86.6) | 661 (78.5) | 10 (71.4) | 1,510 (82.7) | 107 (90.7) |
| Black | 16,893 (3.4) | 2,017 (6.1) | 81 (4.8) | 35 (3.6) | 35 (4.2) | 0 (0.00) | 70 (3.8) | 5 (4.2) |

| | | | | | | | | |
|---|---|---|---|---|---|---|---|---|
| Hispanic | 8,622 (1.8) | 846 (2.6) | 34 (2.0) | 21 (2.2) | 9 (1.1) | 1 (7.1) | 31 (1.7) | 2 (1.7) |
| Asian | 5,277 (1.1) | 780 (2.4) | 47 (2.8) | 53 (5.5) | 112 (13.3) | 0 (0.00) | 165 (9.0) | 1 (0.85) |
| Other | 1,871 (0.38) | 149 (0.45) | 4 (0.24) | 1 (0.10) | 4 (0.48) | 0 (0.00) | 5 (0.27) | 0 (0.00) |
| Not known | 5,740 (1.2) | 622 (1.9) | 67 (4.0) | 20 (2.1) | 21 (2.5) | 3 (21.4) | 44 (2.4) | 3 (2.5) |
| | **UK Biobank** | | | | | | | |
| White | 424,669 (94.5) | 4,819 (88.6) | 9,961 (92.9) | 6,126 (85.8) | 282 (30.0) | 105 (60.0) | 6,513 (78.9) | 399 (89.1) |
| Black | 7,352 (1.6) | 174 (3.2) | 168 (1.6) | 24 (0.34) | 10 (1.1) | 4 (2.3) | 38 (0.46) | 11 (2.5) |
| Asian | 8,865 (2.0) | 289 (5.3) | 360 (3.4) | 825 (11.6) | 626 (66.6) | 65 (37.1) | 1,516 (18.4) | 19 (4.2) |
| Other | 6,752 (1.5) | 133 (2.4) | 173 (1.6) | 117 (1.6) | 18 (1.9) | 0 (0.00) | 135 (1.6) | 13 (2.9) |
| Not known | 1,533 (0.34) | 23 (0.42) | 66 (0.62) | 48 (0.67) | 4 (0.43) | 1 (0.57) | 53 (0.64) | 6 (1.3) |

[1]This table only presents cohort that have collected data on ethnicity. All values are N (%).
[2]All vegetarians include lacto-ovo, lacto, and ovo vegetarians.

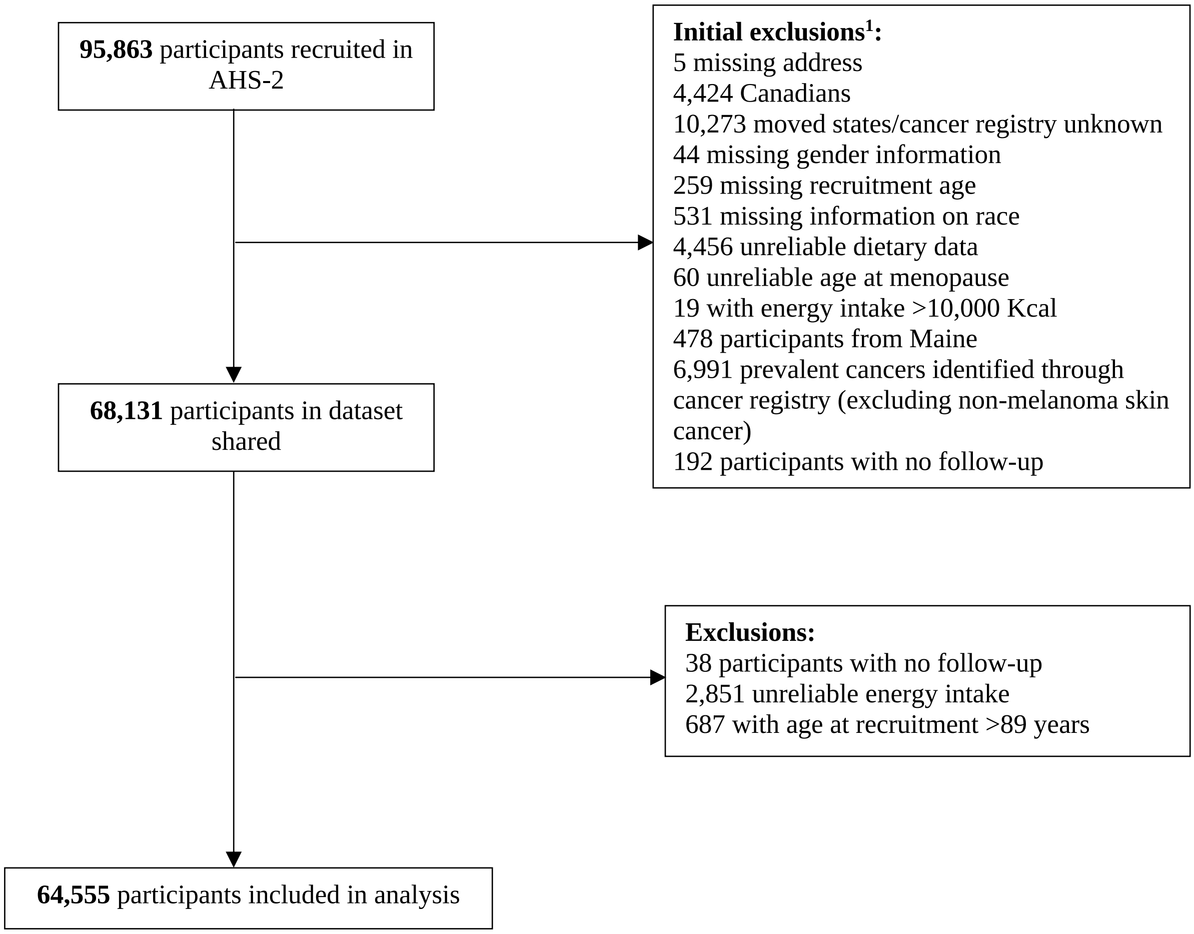

**Figure S1.** Flow diagram of exclusion criteria in Adventist Health Study-2.
[1]Initial exclusions refer to those made by collaborators prior to sharing the data.

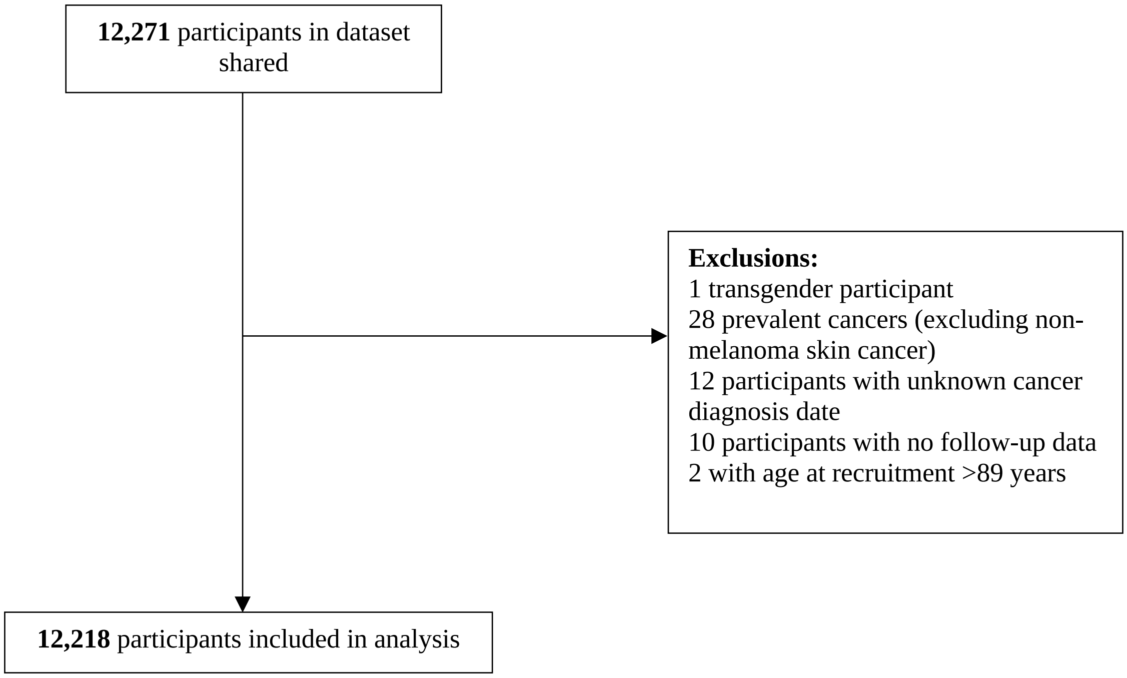

**Figure S2.** Flow diagram of exclusion criteria in Centre for cArdiometabolic Risk Reduction in South Asia 1.

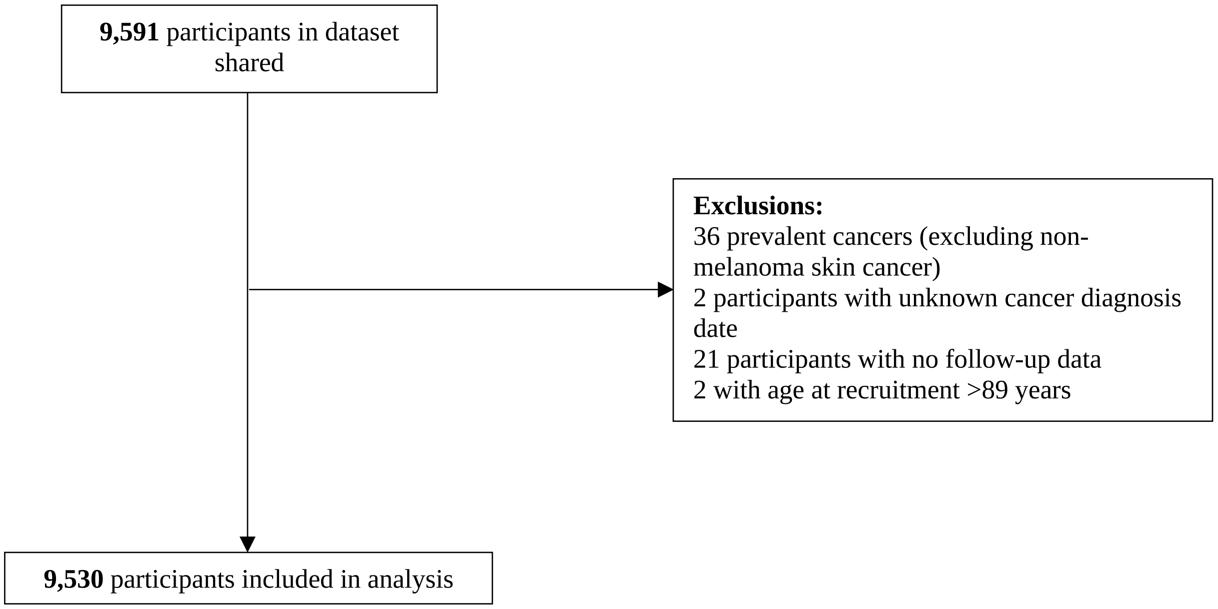

**Figure S3.** Flow diagram of exclusion criteria in Centre for cArdiometabolic Risk Reduction in South Asia 2.

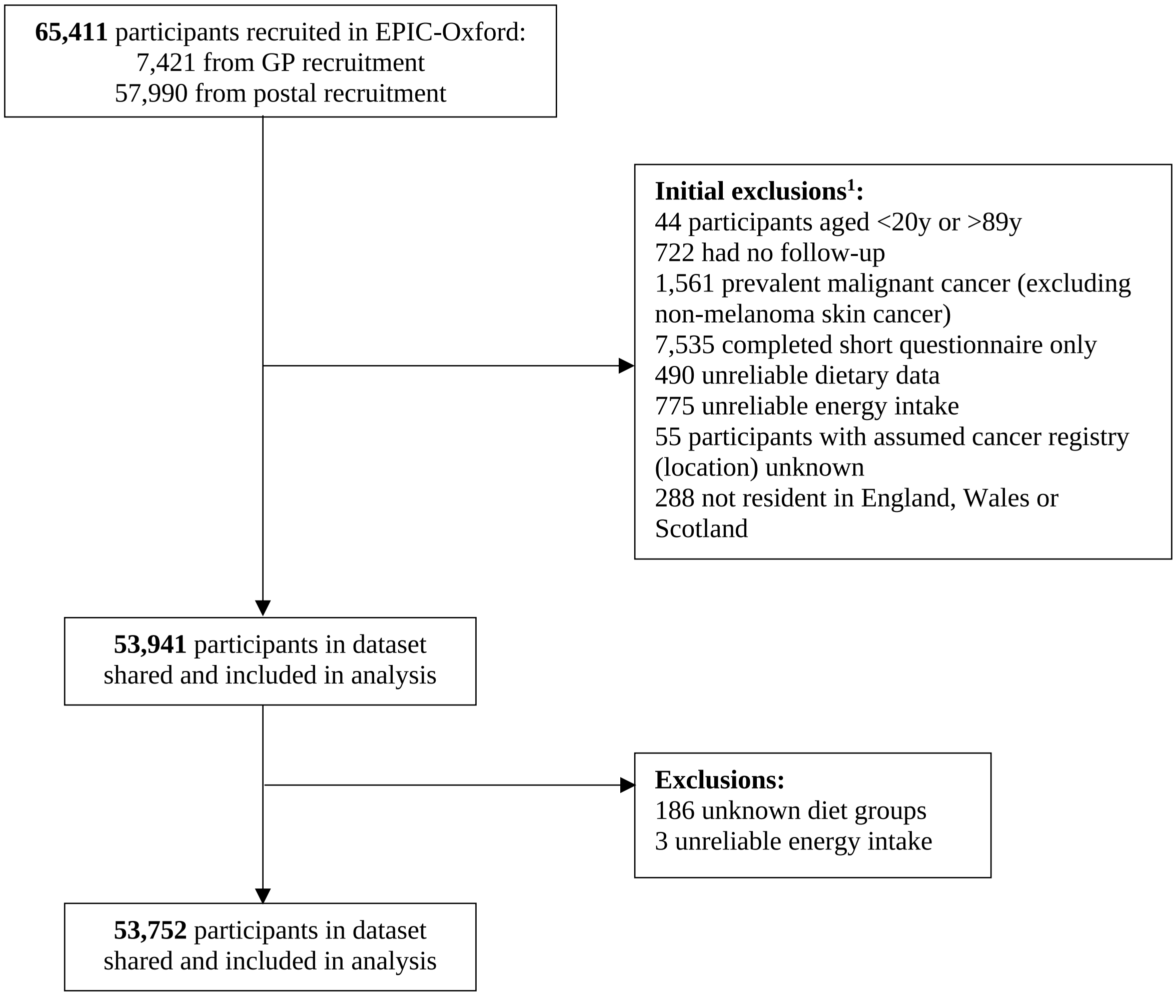

**Figure S4.** Flow diagram of exclusion criteria in EPIC-Oxford.
[1]Initial exclusions refer to those made by collaborators prior to sharing the data.

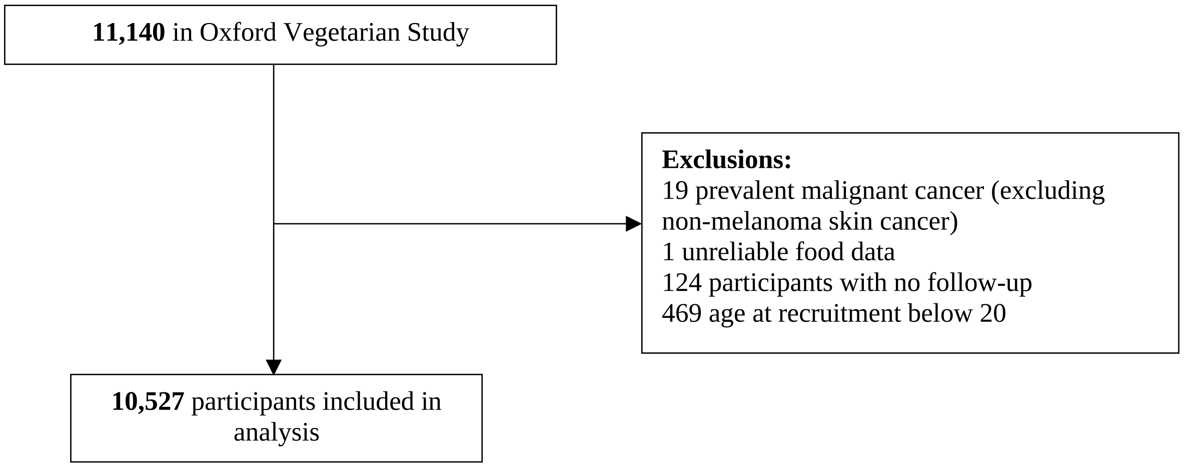

**Figure S5.** Flow diagram of exclusion criteria in Oxford Vegetarian Study.

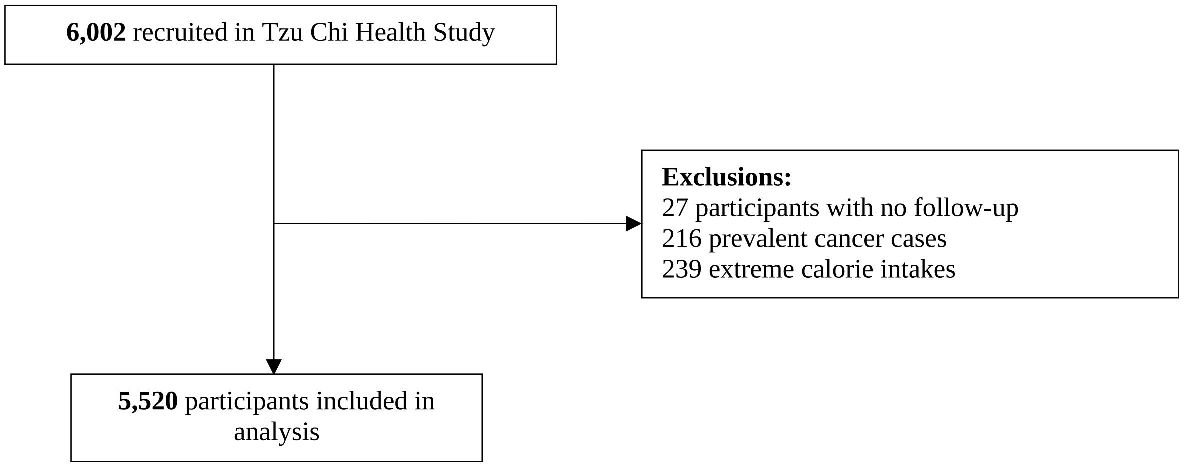

**Figure S6.** Flow diagram of exclusion criteria in Tzu Chi Health Study.

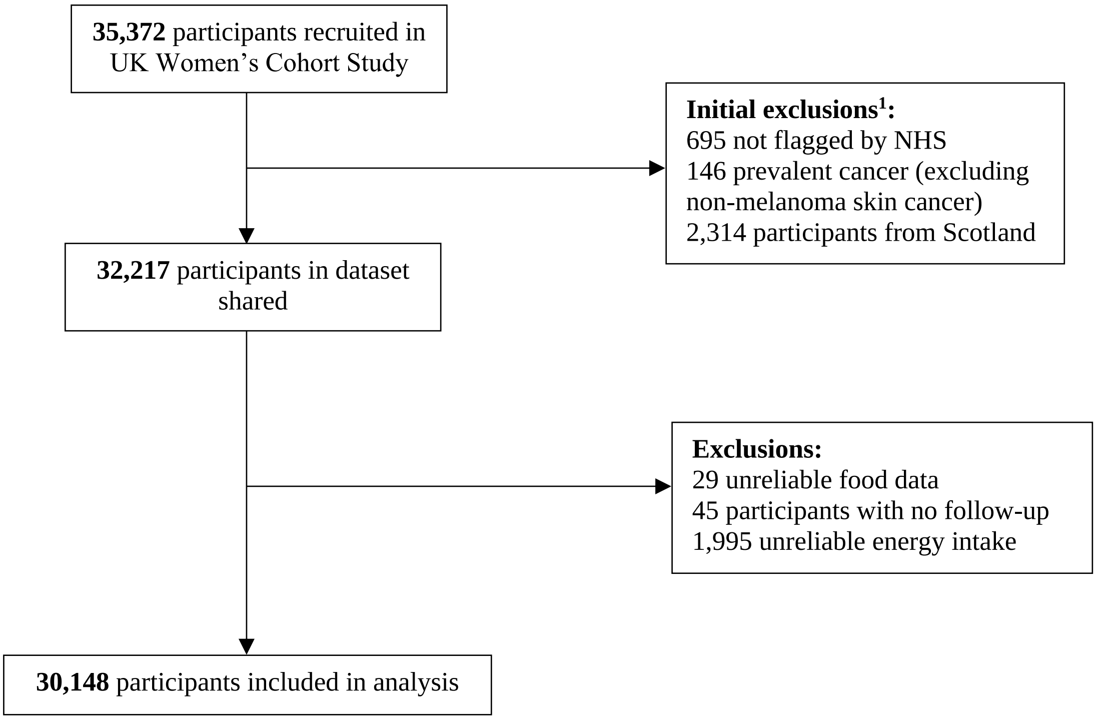

**Figure S7.** Flow diagram of exclusion criteria in UK Women’s Cohort Study.
[1]Initial exclusions refer to those made by collaborators prior to sharing the data.

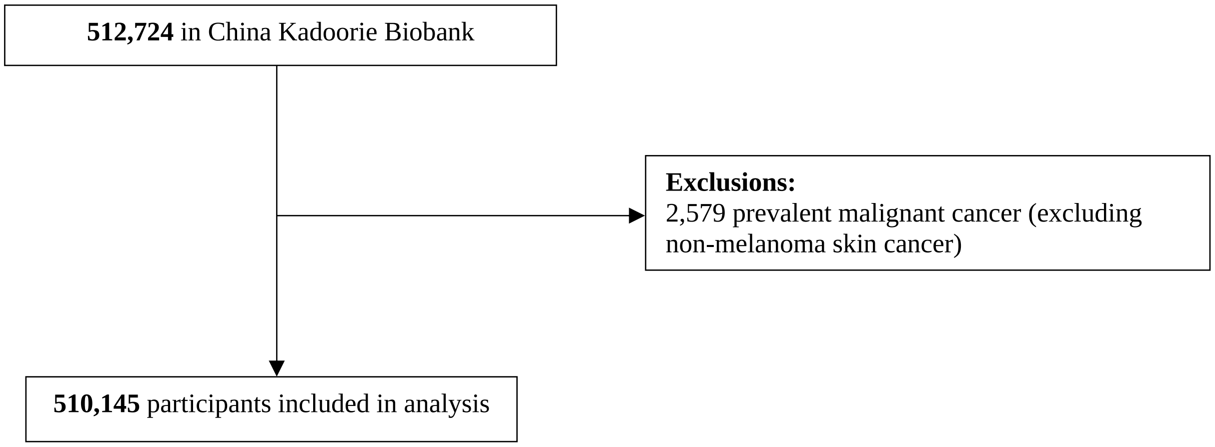

**Figure S8.** Flow diagram of exclusion criteria in China Kadoorie Biobank.

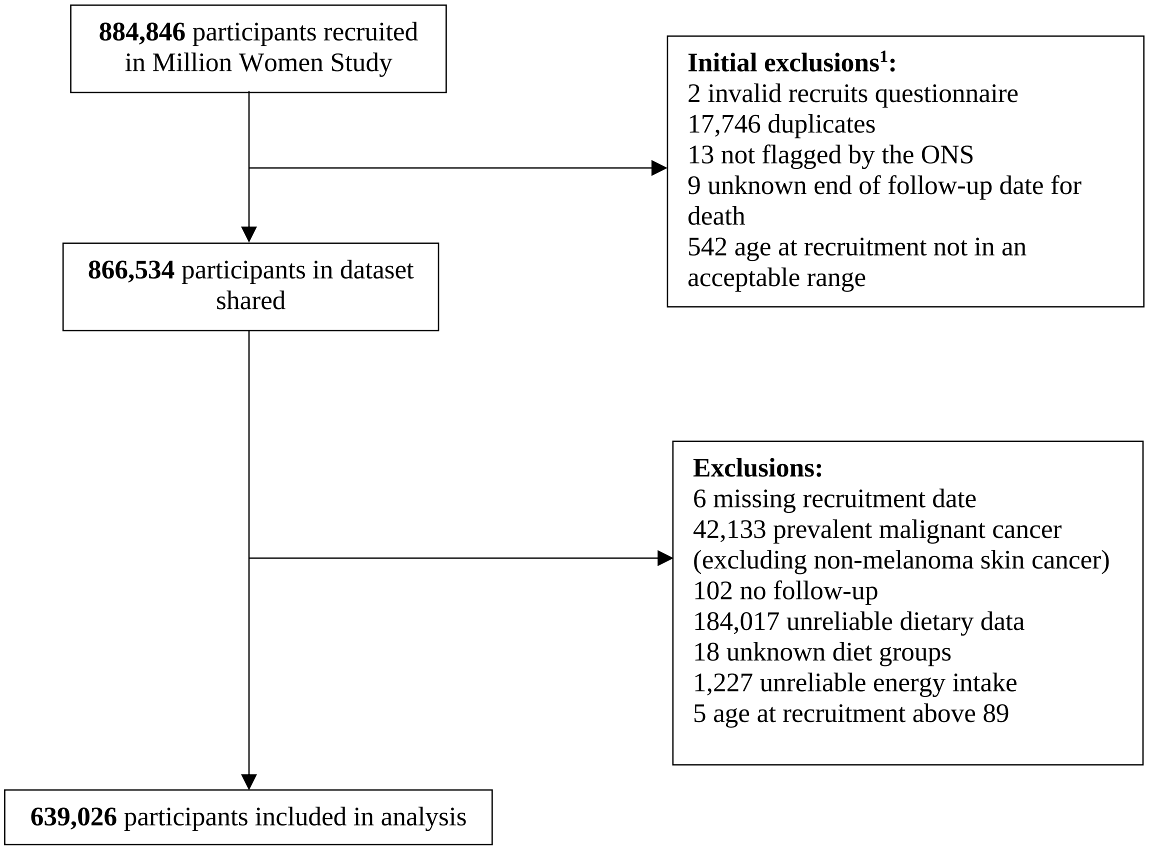

**Figure S9.** Flow diagram of exclusion criteria in Million Women Study.
[1]Initial exclusions refer to those made by collaborators prior to sharing the data.

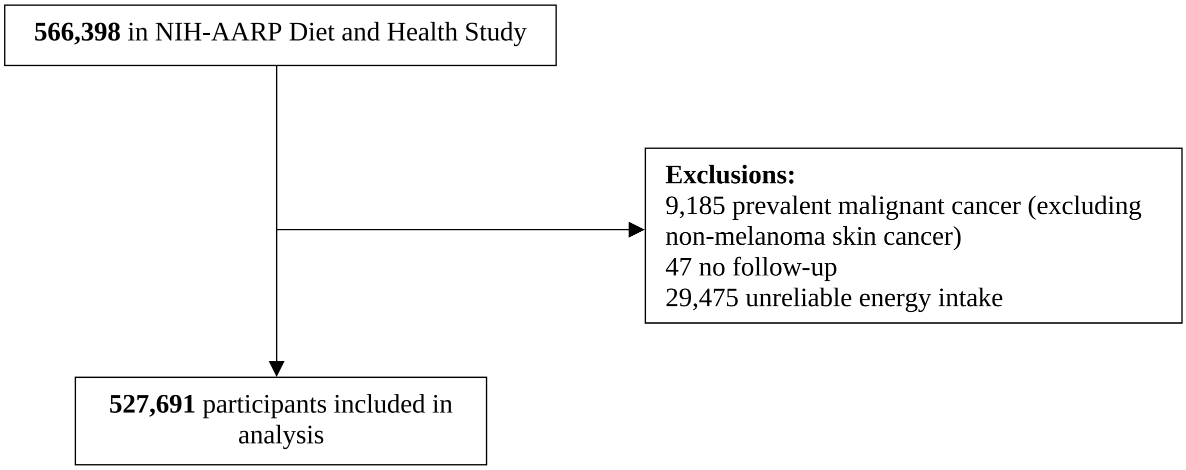

**Figure S10.** Flow diagram of exclusion criteria in NIH-AARP Diet and Health Study.

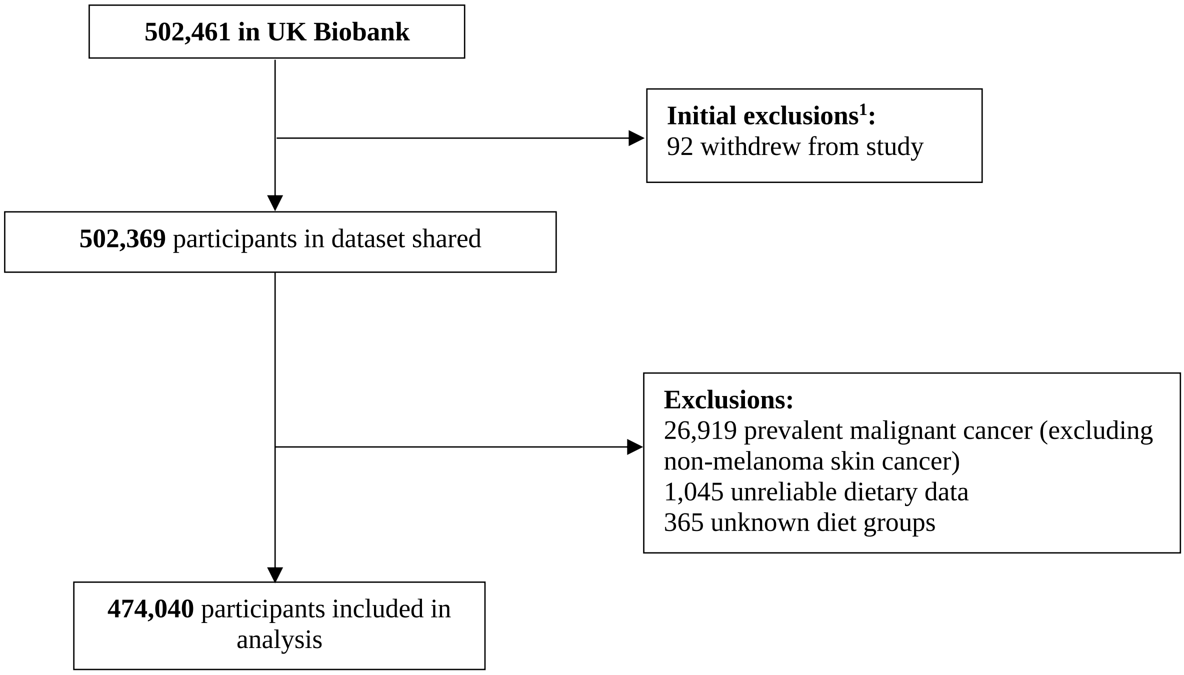

**Figure S11.** Flow diagram of exclusion criteria in UK Biobank.
[1]Initial exclusions refer to those made by collaborators prior to sharing the data.
