## Supplementary Methods for "Methods and participant characteristics in the Cancer Risk in Vegetarians Consortium: A cross-sectional analysis across 11 prospective studies"

### Data collection

After Data Transfer Agreements between the University of Oxford and the collaborating institutions were signed, a data dictionary including details of exposures, outcomes and potential confounders of interest was shared with the collaborators. The datasets with all the variables of interest were then shared by the collaborators using a secure platform managed by the University of Oxford. Due to the regulations governing personal data held by the Health and Welfare Data Science Centre (HWDC) in Taiwan, the collaborators from the Tzu Chi Health Study ran all the analyses at HWDC and shared the summary statistics only with the Cancer Risk in Vegetarians Consortium.

### Profile of studies included in the consortium

#### Cohorts with large proportions of vegetarians

*Adventist Health Study-2*

The Adventist Health Study-2 is a prospective cohort initiated in 2002 with the aim to study the associations between diet and health outcomes among Seventh-day Adventists, a unique health-oriented population with diverse dietary habits. Over a period of five years, around 96,000 Seventh-day Adventist adults (≥30 years old) from the United States and Canada were recruited. In this Consortium, only participants from the United States were included. Approval for the Adventist Health Study-2 study was obtained from the Loma Linda University Human Subjects Committee Institutional Review Board, and written informed consent was acquired from all participants upon enrolment (1). The baseline questionnaire consisted of sections on diet and vegetarian-specific food consumption, physical activity, medical and female reproductive history, and supplement use (1).

*Centre for cArdiometabolic Risk Reduction in South Asia 1*

The Centre for cArdiometabolic Risk Reduction in South Asia 1 (CARRS-1) study started in 2010 and recruited adult participants (20–69 years old) until 2011 (2, 3). This study aimed to capture the prevalence, trends, and risk factors of cardio-metabolic diseases. Participants were recruited using a multi-stage cluster sampling method to obtain a representative cohort from three major cities in South Asia: Delhi (North India), Chennai (South India), and Karachi (Pakistan). In this analysis, only data from the India sites were included. The study followed the Declaration of Helsinki and all procedures involving human participants were approved by the Emory University Institutional Review Board, the Public Health Foundation of India Ethics Review Committee, the Aga Khan University Ethics Review Committee, and the Madras Diabetes Research Foundation Ethics Review Committee. Written informed consent was obtained from all participants.

A baseline questionnaire was used to collect data on participants' socio-demographic and lifestyle characteristics as well as to assess their dietary intake. Five follow-up questionnaires were subsequently administered, with dietary information re-evaluated during the fifth follow-up (2017–2018).

*Centre for cArdiometabolic Risk Reduction in South Asia 2*

In CARRS-2 (2), participants were recruited through independent probability sampling in the same cities as in CARRS-1. This recruitment process took place between 2014 and 2016 and followed methods similar to those used in CARRS-1. There was no overlap in participants between the two studies. One follow-up assessment was completed 4 years after the baseline survey, with a participation rate of 90% similar to CARRS-1.

*EPIC-Oxford*

The EPIC-Oxford cohort was established to recruit participants with a wide range of diets by targeting vegetarians as well as the general UK population. Participants aged over 20 years across the UK were recruited in the EPIC-Oxford study between 1993 and 1999 through general practitioners and postal questionnaires. Ethical clearance was obtained from a multicentre research ethics committee (Scotland A Research Ethics Committee) and all participants provided written informed consent (4). The baseline questionnaire recorded information on participants' diet as well as socio-demographic, lifestyle and health factors. Three follow-up surveys were conducted and diet was reassessed at the third follow-up which was ~15 years (i.e., around 2010) after recruitment.

*Oxford Vegetarian Study*

The Oxford Vegetarian Study is a prospective cohort study which recruited participants across the UK between 1980 and 1984 (5). This study mainly targeted recruitment of vegetarians through the Vegetarian Society of the UK, publicity in the national and local media, and word of mouth via participants already recruited. Non-vegetarian participants were recruited as friends and relatives of the vegetarian participants. Upon entry into the study, participants were required to complete a questionnaire that assessed their diet and other lifestyle factors.

*Tzu Chi Health Study*

The Tzu Chi Health study was established to investigate diet and health outcomes in vegetarians in Taiwan (6). The study recruited 6,002 participants from 2007 to 2009, of which 77% were certified Tzu

Chi volunteers and 23% were their families or individuals who came to the Buddhist Dalin Tzu Chi Hospital for routine health examination. The study was approved by the Institutional Review Board of Dalin Tzu Chi Hospital, and all participants gave written informed consent. All participants underwent a thorough health examination at baseline and were interviewed using a questionnaire to collect demographics, medical history, lifestyle characteristics (e.g. smoking, alcohol, leisure time physical activities), and dietary intake (a questionnaire on vegetarian dietary practice [type and duration] and a quantitative food frequency questionnaire [FFQ]).

*UK Women's Cohort Study*

The UK Women's Cohort Study is a prospective cohort study established in the early 1990s to study the association between diet and cancer incidence and mortality in UK women. Middle-aged women were taken from responders to the World Cancer Research Fund's direct mail survey from across the UK. This cohort aimed to include substantial numbers of vegetarians, fish eaters and meat eaters. Ethical clearance was obtained from 174 local research ethics committees and participants' consent was obtained (7). A self-administered questionnaire was posted to the participants between 1995 and 1998, which assessed the diet, socioeconomic, health and lifestyle factors of the participants.

A repeat FFQ was posted to a random subsample of 2,200 participants in November 2000 and completed by 1,918 participants.

<u>Very large cohort studies</u>

*China Kadoorie Biobank*

The China Kadoorie Biobank is a prospective cohort study of 512,715 participants recruited from 5 urban and 5 rural areas in China. The recruitment process took place between 2004 and 2008. A baseline interviewer-administered electronic questionnaire was used to assess the participants' socio-demographics, medical history, diet and lifestyle factors. Physical measurements (e.g. anthropometrics) were measured using standard protocols. Two follow-up surveys were subsequently conducted on subsamples of participants in late 2008 and 2013–2014, respectively. Ethics clearance for this study was obtained from the Oxford University Tropical Research Ethics Committee, the Chinese Centre for Disease Control and Prevention (CDC) Ethical Review Committee and the local CDC of each study area. Participants also provided written informed consent at recruitment (8).

*Million Women Study*

Invitations to participate in the Million Women Study were sent out to women who were also invited to the National Health Service (NHS) Breast Screening Programme in England and Scotland between 1996 and 2001. Ethical approval was obtained from the Oxford and Anglia Multi-Centre Research Ethics Committee, and all women provided written consent at study recruitment (9). The baseline questionnaire asked about participants' social, demographic and lifestyle factors. Follow-up surveys were conducted every 3–5 years (9). The first follow-up questionnaire was sent out an average of 3.3 years after recruitment and included dietary questions for the first time, therefore, this time point was considered as the baseline for use in this consortium (10). The Oxford WebQ, a validated 24-hour dietary assessment (11), was administered in a sub-sample of women since 2010 to assess dietary intake over time (9).

*NIH-AARP Diet and Health Study*

The NIH-AARP Diet and Health Study (NIH-AARP) is a prospective cohort study which recruited men and women aged 50–71 years from six US states (California, Florida, Pennsylvania, New Jersey, North Carolina and Louisiana) and two US metropolitan areas (Atlanta, Georgia and Detroit, Michigan) (12). The self-administered baseline questionnaire was sent out in 1995–1996 to 3.5 million adults and 18% returned the questionnaire, out of which 566,398 who successfully completed the questionnaire formed part of this consortium. This baseline questionnaire collected information on participants' demographics, anthropometrics (self-reported), lifestyle, usual dietary intake, family history of cancer and reproductive/gynaecologic history. Six months after the completion of the baseline questionnaire, a second questionnaire was sent out to collect additional information on risk factors for cancer; 332,913 returned the completed questionnaire. Ethical approval for this study was obtained by the Special Studies Institutional Review Board of the National Cancer Institute and all participants provided informed consent.

*UK Biobank*

A total of 9.2 million adults aged 40–70 years and registered to the UK National Health Service (NHS) records were invited to participate in the UK Biobank (13). Invited individuals lived within a radius of 25 miles of one of the 22 assessment centres in the UK. Over 500,000 who successfully completed the baseline questionnaire (5.5% response rate) and consented to participate were included in this cohort. The recruitment process took place between 2006 and 2010. This study obtained ethical approval from the NHS North West Multicentre Research Ethics Committee (21/NW/0157).

A touchscreen questionnaire was used at baseline to collect information on participants' diet, socio-demographic, reproductive, and lifestyle factors. Anthropometric measurements, including height and weight, were also taken by trained professionals. A subsample of participants completed up to five Oxford WebQ 24-hour dietary assessments between April 2009 and April 2012, while another subsample completed the baseline touchscreen questionnaire up to three times during the follow-up period.

**Data harmonization**

Harmonization algorithms were established *a priori* for each variable. However, amendments and adjustments to the recoding instructions were implemented upon receiving all individual datasets to ensure the consistent creation of variables across all studies, as elaborated in the following sections.

<u>Socio-demographic variables</u>

In this section of the supplementary methods, we provide additional details on the harmonization process for each covariate. Table S1 can be referred to for an overview of how the original variables were recoded to achieve harmonization.

*Marital status*

In all the studies, participants were asked about their marital status. In the Adventist Health Study-2, participants had to choose from one of the following options: never married, first marriage, remarried, common law marriage, separated, divorced and widowed, while in the Million Women Study and Oxford Vegetarian Study participants only had the option to indicate whether they were living with a partner or not, or if they were married or unmarried, respectively. Therefore, in studies where marital status included more details, these were collapsed into a binary variable, "living with a partner". In the UK Biobank, unlike in the other studies, living with a partner was derived from a question asked at recruitment in which participants answered if they lived with anyone else in their household. If the participants reported to be living with a husband, wife, or partner in their household they were classified as "living with a partner", whereas if the participant reported to be living with any other person, or if they indicated to be living alone, they were categorised as "not living with a partner".

*Educational status*

The responses for educational status were specific to each study and aligned with the education system in the country where the cohort was established. For this consortium, educational status was categorised into “less than secondary/high school or equivalent”, “secondary/high school or equivalent”, and “university degree or equivalent”. Being illiterate, having less than 11 years of education, primary school, no formal school, and UK Certificate of Secondary Education (CSE) corresponds to “less than secondary/high school”. In the “secondary/high school” category, secondary school, high school, junior college, UK General Certificate of Education Ordinary Level (GCE O-level), GCE Advanced Level (A-level) or 12 years of school were included. The “university degree” category corresponds to undergraduate, masters, or doctoral degrees, or equivalent professional qualifications.

In the Oxford Vegetarian Study, participants’ educational status was not assessed. Instead, the questionnaire asked about the occupation of participants, which was used to determine their social class. Participants’ social class was subsequently used as a proxy to estimate the educational status of participants in this study. Individuals who reported their social class as Grade 5 or identified themselves as housewife were categorised under the “less than secondary/high school” category. Those who reported grades 2, 3 or 4 were categorised into the “secondary/high school” category, while those who reported grade 1 were categorised into the “university degree or equivalent” category.

*Ethnic group*

The ethnicity of participants was recorded at baseline in most of the studies. The US-based cohorts included the broadest range of options for ethnicity, with, for example, twenty options listed in the Adventist Health Study-2, while less detailed options were available in the UK-based cohorts. In CARRS-1, CARRS-2, China Kadoorie Biobank, the Oxford Vegetarian Study, and the Tzu Chi Health Study, the ethnicity of participants was not assessed. For these cohorts, the country where the study was conducted was used as a proxy to determine the ethnic group. All participants in CARRS-1 and CARRS-2 were assumed to be South Asians, while those from the China Kadoorie Biobank and the Tzu Chi Health Study were assumed to be East Asians. In the Oxford Vegetarian Study, all participants were assumed to be of White European ancestry. Finally, all participants were categorised into one of the following categories: “White”, “Black”, “Hispanic”, “Asian”, and “Other or not known”.

*Cigarette smoking*

Smoking status was assessed at baseline in all the studies and participants were categorised as “never”, “previous” or “current” smokers. Five studies had information on smoking status in this same format, whereas in the Adventist Health Study-2, CARRS-1, CARRS-2, EPIC-Oxford, Million Women Study and Oxford Vegetarian Study, the smoking status of participants was derived using responses from multiple questions. In the Adventist Health Study-2, participants were asked “Have you ever smoked regularly?” and “if you have ever smoked cigarettes regularly, please mark the usual number you smoke(d) per day or week during each of the given time periods" to determine their smoking status. If participants reported that they ever smoked, the number of cigarettes smoked question was used to determine whether they were previous or current smokers.

In CARRS-1 and CARRS-2, smoking status was determined using questions on cigarette and beedi smoking (a common local form of tobacco). Participants were asked "have you ever used tobacco in any form (smoking, chewing, snuff, etc.)?", "in what forms have you consumed tobacco?", and "do you currently consume tobacco?". Responses from these three questions were used to classify participants as "never", "previous" or "current" smokers.

*Body Mass Index (BMI)*

In some of the studies, height and weight were measured by trained staff at baseline and this was used to determine a participant's BMI, which was calculated by taking the participant's weight in kilograms and dividing it by their squared standing height in metres. However, in the Adventist Health Study-2, EPIC-Oxford, Oxford Vegetarian Study, UK Women's Cohort Study, Million Women Study and the NIH-AARP, height and weight were self-reported. In the Million Women Study, weight was missing for a large number of women at the first resurvey, the time point when dietary intake was collected; for these women, their weight from the recruitment survey was used if available.

*Physical activity*

In all the cohorts, physical activity was assessed at recruitment using a series of questions such as the types of activity, the number of days per week that they were performed, and for how many minutes or hours. The physical activity questionnaire used in most of the studies had been previously validated and was found appropriate for use in the respective study populations. In the Adventist Health Study-2, physical activity over the past 12 months was assessed using a series of questions, which were examined for validity and reliability against 7-day recall activity levels and pedometer readings in a subsample of 165 black Seventh-day Adventists from Southern California (14). In CARRS-1, the International Physical Activity Questionnaire (IPAQ) – short version was used to assess physical activity (3), while in CARRS-2, the global physical activity questionnaire (GPAQ) was used. In the Tzu Chi Health Study, questions were asked about leisure time physical activities. In the China Kadoorie Biobank, questions on physical activity were adapted from validated questionnaires used in several other studies, including the EPIC study and the Shanghai Women's Health Study. The questions were adapted to the study population after a pilot study was conducted in a subsample of participants from the China Kadoorie Biobank (15).

Using responses from these questions, the metabolic equivalent of task per day (MET) was calculated in each individual cohort, and study-specific cut-off points were used to categorise participants as "inactive", "moderately active" or "highly active".

In the China Kadoorie Biobank, because of the way the different types of activities were combined to generate the METs, it was not appropriate to apply cut-off points and determine the physical activity level in the same way as in the other studies. For example, sitting for longer hours added up to the total METs and hence gave a higher value. Therefore, sex-specific tertiles of the METs were used to categorise participants into one of the three physical activity categories. Unlike in the other cohorts, in the Adventist Health Study-2 the multiple questions of physical activity were computed to generate the minutes of exercise per week, which was used to categorise participants into three levels of physical activity categories. In the Oxford Vegetarian Study, the frequency of different activities was combined, and tertiles of this new combined frequency were used to assign participants as "inactive", "moderately active" or "highly active".

*Alcohol intake*

Alcohol intake was assessed at baseline across all cohorts. In the Adventist Health Study-2, participants were asked how often they consumed beer (12 oz.), wine coolers (12 oz.), wine (3.5 oz.) or liquor (1 oz.) in the past one year, with the possible responses ranging from "less than 1 drink per month" to "7+ drinks per week". In EPIC-Oxford, participants were asked about their frequency of drinking beer or cider, wine, port, sherry, vermouth, liqueurs, and spirits in the last 12 months, with the possible responses ranging from "never or less than once/month" to "6+ per day". In the UK Women's Cohort Study, participants were asked how often they drank alcohol, with the possible responses being "more than once a day", "once a day", "a few times a week", "once a week", "a couple of times a month", "once a month", "less than once a month", and "never drink alcohol". In the China Kadoorie Biobank, participants were asked how often they drank alcohol during the past 12 months, with the possible responses being "never or almost never", "only occasionally", "only at certain seasons", "every month but less than weekly", and "usually at least once a week". If participants in this study responded "usually at least once a week", they were further asked "1-2 days/week", "3-5 days/week", or "daily or almost every day". In the Million Women Study, participants were asked how many drinks of alcohol they consumed each week. In the NIH-AARP, participants were asked how often they consumed beer, wine, and liquor in the last 12 months, with the 10 possible responses ranging from "never" to "6+ times per day". In the UK Biobank, participants were asked how often they drank alcohol with the possible responses being "daily or almost daily", "three or four times a week", "once or twice a week", "one to three times a month", "special occasions only", "never", and "prefer not to answer". Participants were also asked about their weekly and monthly intake of pints of beer, glasses of red wine, glasses of white wine/champagne, glasses of fortified wine, measures of spirits/liqueurs and glasses of other alcohol. Based on these responses, the amounts of alcohol consumed in grams per day were derived using standard portion sizes.

*History of diabetes*

History of diabetes was recorded as a binary variable in most of the cohorts at baseline. However, in the Adventist Health Study-2, participants were asked if they have ever been diagnosed by a doctor with diabetes, and if their response was "yes", to report the number of years since they were first diagnosed. Therefore, for participants who responded to the number of years since first diagnosis regardless of whether the condition is still active, they were considered as having a history of diabetes.

Women-specific variables

*Age at menarche*

The age at first menstrual period was asked as an open-ended question in most of the studies at the beginning of the study. However, in the Adventist Health Study-2, participants had to choose among 10 pre-coded options which ranged from "never" to "17 years or older". Participants in NIH-AARP were asked "how old were you when you had your first menstrual period?" and had to choose among one of the following: "10 or younger", "11–12", "13–14", and "15 or older". To ensure consistency in categorization, the options from the Adventist Health Study-2 were collapsed to match the age ranges, and similarly the continuous menarche variable in other cohorts was classified as "≤10 years", "11–12 years", "13–14 years" and "≥15 years".

*Parity*

The number of children women had was asked in all the prospective cohort studies included in this consortium. This was an open-ended question in most of the studies. In the NIH-AARP, participants had to select among the following options: "none", "one", "two", "3–4", "5–9", "10 or more". In the Adventist Health Study-2, parity was defined based on multiple questions such as the birth year for the first to the tenth child, number of months of pregnancy, pregnancy outcome, sex of child and birth weight. If the woman answered to any one of those questions, that contributed to the number of live births. If a woman responded to having more than 10 live births, this was truncated to count as having 10 children. The continuous parity variable from the different studies and the multiple categories in the

Adventist Health Study-2 were subsequently collapsed into the following final categories: “none”, “one”, “two”, “3–4”, “5–9”, and “≥10”.

*Age at first birth*

Age at first birth was assessed at study recruitment in all studies except CARRS-1, CARRS-2 and Tzu Chi Health Study. In most studies, women were asked “how old were you when your first child was born” and they had to respond in years. In the Adventist Health Study-2, the age at first birth was determined using the woman’s response to the question about the birth year of the first child. In the NIH-AARP, women had to select their age at first birth from eight pre-coded options: “<16”, “16–19”, “20–24”, “25–29”, “30–34”, “35–39”, and “≥40” years. Therefore, in this consortium, women were classified according to the same age ranges as in NIH-AARP.

*Menopausal status*

Menopausal status in this consortium was initially defined as premenopausal, peri-menopausal or postmenopausal. However, in most studies, no information on peri-menopausal status was available, and in studies which had such information, peri-menopausal women were likely to transition to postmenopausal during the follow-up time. Hence, women were finally categorised as pre- and postmenopausal based on self-reported menopausal status at recruitment or, if unknown, using the age at recruitment.

The average age at menopause in the UK and US is estimated to be around 51 years (16, 17), while in Asian countries a lower average age at menopause of 48 years (18, 19) has been reported. Therefore, to reach a midpoint for participants from both the Western and Asian cohorts, a cut-off age of 50 years at recruitment was used to determine menopausal status in this consortium. Women were classified as postmenopausal if they:

- Reported having undergone menopause (if information was available)
- Had a bilateral oophorectomy
- Were ≥50 years at study recruitment

Women were classified as premenopausal if they:

- Were less than 50 years at study recruitment and did not report their menopausal status

*Age at menopause*

Age at the last menstrual period was recorded as a continuous variable in all the cohorts except in the NIH-AARP. In some studies, such as EPIC-Oxford and UK Women's Cohort Study, where participants were asked "when did you last have a natural menstrual period?", responses were restricted to postmenopausal women since women who are still menstruating could also respond to that question. However, in studies like the Million Women Study and Tzu Chi Health Study, women were asked "how old were you when you had your menopause?", and in the UK Biobank women answered the question "how old were you when your periods stopped?", directly targeting postmenopausal women. In EPIC-Oxford, the lowest age at menopause was truncated at 40 years, as it could not be determined whether reported ages at menopause before the age of 40 years were reliable. The continuous age at menopause variable was finally classified to match the categories used in NIH-AARP: "<40", "40–44", "45–49", "50–54", and "≥55".

*Ever used oral contraceptives and ever used hormone replacement therapy*

The use of oral contraceptives was assessed in all the studies. In EPIC-Oxford, the Oxford Vegetarian Study, and the UK Women's Cohort Study, separate questions were asked with binary categories. In the other studies, a single question asking about never, ever or current use of oral contraceptives was available (Table S1). For the latter studies, "never" was coded as "no" and the "ever use" and "current use" were coded as "yes".

Out of the 11 studies, seven assessed the ever use of hormone replacement therapy (HRT) at recruitment. In EPIC-Oxford, the UK Women's Cohort Study, and NIH-AARP, women were asked if they had ever used HRT and had to choose between "yes" or "no". In the Adventist Health Study-2 and the Million Women Study, women had to choose between the options: "never", "ever" or "current" and these were re-coded as follows: "never" as "no", and "ever use" and "current use" as "yes".

<u>Diet groups</u>

Diet groups were defined *a priori* to facilitate the request of the relevant variables from the collaborators. Seven diet groups were predefined: meat eaters, poultry eaters, pescatarians, lacto-ovo vegetarians, lacto vegetarians, ovo vegetarians, and vegans. An "all vegetarian" group was also created by combining the lacto-ovo, lacto and ovo vegetarians, for comparison with the non-vegetarian diet groups. Before generating the diet groups, participants with more than 80% of missing FFQ data were excluded, except in EPIC-Oxford where this exclusion had already been made by collaborators before sharing the dataset.

Initially, a strict definition of 0 g/day of relevant animal foods in the respective diet groups was considered. However, in some of the cohorts where a “never/rarely” category was included in the FFQ but a zero category was not, participants in the lowest consumption frequency had been assigned a small amount of a food when the weights of the foods were calculated. Therefore, participants across the individual cohorts were assigned to a diet group based on the consumption frequency of animal foods, including red meat, processed meat, poultry, offal, oily fish, non-oily fish, dairy milk and dairy products, and eggs. An exception was the NIH-AARP study where the food weights were used to assign participants to the appropriate diet groups (further elaborated in the section below).

Consumption frequencies were multiplied by the number of portions (where a semi-quantitative FFQ was used) and the standard portion size (country-specific) to obtain the food weights in g/day. For two studies, the China Kadoorie Biobank and UK Biobank, the mean intakes for the different diet groups from resurvey 2 and the multiple 24-h dietary assessments, respectively, were assigned to the baseline diet groups to obtain the food weights. It is important to note that results from these two cohorts should be interpreted carefully because they are not actual intakes at baseline.

*Adventist Health Study-2*

Diet over the last year in this study was assessed at baseline using a self-administered semi-quantitative FFQ that included 130 food items (20). The consumption frequency ranged from “never/rarely” to “more than 4 times per day”. The FFQ had been previously validated for foods and nutrients against six repeated 24-h dietary recalls. Participants were categorised into the seven diet groups described in **Table 2**.

*CARRS-1*

In this study, diet over the last year was assessed using a 26-item non-quantitative FFQ adapted from the INTERHEART study. The FFQ included four consumption frequency categories: “never or less than once a month”, “monthly”, “weekly” and “daily”. Participants were assigned to one of the seven diet groups using the definitions shown in **Table 2**.

During the fifth follow-up, with a median (IQR) follow-up time of 5.9 (5.4–6.2) years, a 33-item FFQ with the same categories was used for the repeat assessment of dietary intakes. The categorisation of diet groups at resurvey was done for 7,389 cancer-free participants.

*CARRS-2*

In CARRS-2, a semi-quantitative FFQ was used to assess participants' intake of 33 food groups over the last year. Similar to CARRS-1, the predefined frequencies included "never or less than once a month", "monthly", "weekly" and "daily". Participants were classified into one of the seven diet groups using the definitions shown in **Table 2**.

*EPIC-Oxford*

A semi-quantitative FFQ was used to assess the diet of participants over the past 12 months. Participants completed a 130-item FFQ, which was previously validated (4). The FFQ included nine consumption frequency options ranging from "never or less than once/month" to "6 or more times daily". In addition to the FFQ, the baseline questionnaire asked separately if participants consumed any of the following: meat, fish, dairy products and eggs. Responses to these four questions were used to categorise participants into meat eaters, fish eaters, vegetarians, or vegans (21). This classification was considered sufficient to define the diet groups as this cohort targeted the recruitment of vegetarians. However, it should be noted that at the beginning of the study, EPIC-Oxford was not designed to target vegetarians, and therefore, the first version of the baseline questionnaire did not include the four questions separately on meat, fish, dairy products and eggs. Therefore, for the small minority of participants with no data on these four questions, their FFQ data were used to assign them to a diet group (22).

To further categorise participants into the different subtypes for vegetarian diets and to identify poultry eaters, the FFQ data were used as shown in **Table 2**. Participants were classified as poultry eaters if they were identified as fish eaters if they did not report to eat red or processed meat and consumed poultry more than 1–3 times per month. Lacto vegetarians included participants who were identified as vegetarians and consumed dairy milk or dairy products more than 1–3 times per month and reported eating eggs never or less than once per month. Similarly, ovo vegetarians included participants identified as vegetarians who consumed eggs more than 1–3 times per month and reported intake of dairy products as never or less than once per month.

Diet was reassessed at the third follow-up at a median of 14.3 (interquartile range [IQR]): 13.4–15.2) years using a similar FFQ in a subsample of approximately 28,000 participants. Further classification of diet groups based on these follow-up data used the same criteria as at baseline, in the subsample who were cancer-free at follow-up (n = 27,529) (**Table 2**).

*Oxford Vegetarian Study*

Diet was assessed using a simple FFQ. Similar to EPIC-Oxford, the Oxford Vegetarian Study questionnaire also included four questions which assessed participants' meat, fish, dairy product, and egg consumption as "never", "occasionally" and "regularly". Responses to these four questions were used to classify participants as meat eaters, fish eaters, vegetarians and vegans.

Poultry eaters could not be identified in this study as poultry intake was not assessed in the FFQ. Using the consumption frequency from the FFQ, participants were classified into the different vegetarian diet groups (**Table 2**). Vegetarians identified using the responses from the four questions were further classified as lacto vegetarians if they consumed dairy milk or dairy products more than "less than one per week" and reported never consuming eggs. Ovo vegetarians were defined as those identified being vegetarians using the four questions, in addition to consuming eggs more than "less than once per week" and reported never consuming dairy milk and dairy products.

There were 2,837 participants in this study who subsequently took part in EPIC-Oxford in 1993–1999. Therefore, the baseline dietary assessment in EPIC-Oxford is treated as a repeat dietary assessment for this subsample of participants in the Oxford Vegetarian Study. The diet groups defined using dietary data at these two instances are compared in this paper, and the median follow-up time was 13.6 (IQR: 12.3–14.0) years.

*Tzu Chi Health Study*

The FFQ was adapted from the Nutrition and Health Survey in Taiwan, to include more vegetarian foods and to convert open-ended staple food section to predefined food items. After this modification, the Tzu Chi Health Study (TCHS) FFQ – consisting 64 food/food groups – was modified from the one used was validated among Tzu Chi volunteers who were also TCHS cohort participants, and showed good reliability (through repeated measurements) and validity (with repeated dietary records and biomarkers) (6). Participants were classified as vegetarians if they self-reported as vegetarians in a question asking about vegetarian status, otherwise, they were classified as non-vegetarians. Poultry eaters, pescatarians and the subtypes of vegetarians were then defined by their responses in the FFQ.

*UK Women's Cohort Study*

The FFQ from the UK Women's Cohort Study was adapted from the one used in EPIC-Oxford. It assessed intake of 217 foods and included 10 predefined consumption frequency options which ranged from "never" to "6 or more times per day". The validity of the FFQ was previously assessed through comparison of the nutrient values obtained from the FFQ to the values from a 4-day food diary and fasting blood measures of specific nutrients (23). These consumption frequencies were used to define the seven diet groups as shown in **Table 2**.

A repeat FFQ, almost identical to the baseline FFQ, was completed by 1,652 cancer-free participants at a median follow-up time of 4.8 (IQR: 4.6–5.1) years, and these data were used to categorise participants into the seven diet groups using the same criteria shown in **Table 2**.

*China Kadoorie Biobank*

Diet over the past year in the China Kadoorie Biobank was assessed using a validated interviewer-administered FFQ, consisting of 5 predefined categories: “daily”, “4–6 days/week”, “1–3 days/week”, “monthly” and “never/rarely”. The intake of 12 major food groups was assessed, including rice, wheat products, coarse grain products, red meat, poultry, fish, eggs, total dairy products, fresh vegetables, preserved vegetables, fresh fruit, and soybean products. The FFQ was validated in a sub-sample of 432 participants using 12-day 24-hour recalls as a reference, and findings showed good reproducibility and validity of all the food groups (24)

After completion of the baseline survey, the participants received two follow-up surveys using similar procedures to those of the baseline survey. The second resurvey took place between August 2013 and September 2014, with approximately 25,000 participants completing the questionnaire. Unlike the baseline questionnaire, resurvey 2 used a semi-quantitative FFQ and included more information for a number of foods, e.g. three subtypes of dairy products, i.e. cow milk, yoghurt and other dairy products (e.g. cheese and milk powder). The frequency and portion size information were used to estimate the mean intake for each category of foods at resurvey 2. The mean intakes of each FFQ category at resurvey from the resurvey subsample were then assigned to the baseline sample according to their FFQ categories to provide estimates indicative of mean intakes at baseline. As noted above, these estimates should be interpreted with caution because they are not direct estimates of baseline intakes.

Participants were assigned to the diet groups at baseline as described in **Table 2**. Reclassification of diet groups, using dietary data collected at resurvey 2 (median follow-up time: 8.0 [IQR: 7.4–8.6] years), was performed for 24,586 cancer-free participants using the same criteria as at baseline.

*Million Women Study*

Dietary data in the Million Women Study were collected in the first follow-up questionnaire an average of 3.3 years after recruitment. This questionnaire was distributed between 2000 and 2004 and serves as the baseline for this consortium. The food questionnaire assessed the food intakes of participants during a typical week and included about 130 quantitative or semi-quantitative questions. The short-term repeatability of most of the diet questions was good and, in comparison with intakes of nine macronutrients estimated from 7-day food diaries (with portion sizes weighed or assessed by comparison with photographs), the median energy-adjusted correlation coefficient was 0.55 (ranging from 0.27 for polyunsaturated fat to 0.75 for alcohol) (25).

The quantitative questions asked about types of meat consumed approximately once a week or more often, while the semi-quantitative questions focussed on the number of times the food was eaten per week. The food questionnaire also asked if participants never consumed certain foods. This information was taken into account when classifying participants into the respective diet groups, considering foods such as beef, pork/ham, lamb, kidney, liver/pâté, salami, sausages, eggs, and dairy products.

Meat eaters were defined as participants who consumed at least one type of red and processed meat (e.g. beef, lamb, pork, beef burger/hamburger, bacon, ham, sausages, kidney, or liver/paté), and/or reported using lard for cooking, and/or reported eating meat one or more times per week. Participants were classified as poultry eaters if they reported never eating any red and processed meat but did eat chicken/poultry one or more times per week. Pescatarians were participants who reported eating at least one specific type of fish (e.g. tuna, salmon, cod/haddock, sardines, kippers/herring, mackerel, trout, fish & chips, or other seafood), and/or reported that they ate fish one or more times per week. Vegetarians were defined as those who reported that they never ate red and processed meat, poultry or fish. Vegetarians are further classified into lacto-ovo if they reported eating both dairy and eggs, lacto if they reported eating dairy but no eggs, and ovo vegetarians if they reported eating eggs but no dairy. Vegans were participants who reported never eating any red meat, poultry, fish, eggs, or dairy (**Table 2**).

Throughout 2010–2020, 23,525 cancer-free participants completed a minimum of three and up to 15 Oxford WebQ 24-hour dietary assessments. The median follow-up time was 13.0 (IQR: 11.1–14.2) years. The mean intakes of animal foods across all the 24-hour assessments completed for each participant were used to reclassify diet groups at follow-up based on the criteria in **Table 2**. Participants were considered non-consumers of a food if the mean intake across all 24-hour assessments was 0 g/day.

*NIH-AARP Diet and Health Study*

Diet over the past 12 months was assessed at study recruitment using a self-administered semi-quantitative FFQ, developed at the US National Cancer Institute. The FFQ included a list of 124 food items and the consumption frequency ranged from "never" to "2+ times per day" for foods and "never" to "6+ times per day" for beverages. Portion sizes for each food and beverage were also assessed using three portion size categories. The FFQ was previously validated in a subsample of the NIH-AARP cohort (n = ~2,000) using two non-consecutive 24-hr recalls (26).

The consumption frequency of specific foods was combined with additional questions, such as those about the fat content of the foods (e.g. regular or lean meat), or the type of milk/creamer used for tea and coffee, to code the frequencies using an algorithm developed by the US National Cancer Institute. Based on the coding provided for the foods, it was not possible to identify participants who never ate specific animal-based foods, therefore the diet groups could not be defined in the same way as in the other cohort studies. Hence for this study, food weights provided by the collaborators were used to assign participants to the respective diet groups. A medium serving size was assumed if no serving size was indicated.

To assess intakes of total red and processed meats as well as dairy products, multiple questions were used. To identify participants who "never ate" red and processed meat, and those who "never consumed" dairy products, cut-points of 10 g/day and 5 g/day were used, respectively. For assessing intakes of poultry, fish, and eggs, fewer questions were asked, and hence, a cut-point of 0 g/day was used to identify the never eaters for each food. The seven diet groups were defined as described in **Table 2**.

*UK Biobank*

Diet groups were categorised using the touchscreen questionnaire completed at recruitment which asked participants about the frequency of consumption of processed meat, beef, lamb or mutton, pork, chicken, turkey or other poultry, and oily and non-oily fish. Participants chose a frequency of intake ranging from "Never" to "Once or more daily". Based on these responses, participants were categorised into four diet groups (meat eaters, fish eaters, and vegetarians). Poultry eaters were those who never consumed red and processed meat but consumed poultry. Pescatarians were participants who reported that they never consumed red meat, processed meat, or poultry but ate oily and/or non-oily fish. Participants who reported that they never consumed any meat or fish were assigned to the vegetarian diet group (**Table 2**).

The baseline questionnaire also asked participants to indicate which foods they never ate such as eggs or foods containing eggs, dairy products, wheat products, and/or sugar or foods/drinks containing sugar. Using responses from this question, vegetarians were further classified as lacto vegetarians if they consumed dairy but no eggs and as ovo vegetarians if they consumed eggs but no dairy. Participants were classified as vegans if they reported never eating any meat, fish, eggs, or dairy.

In 2012 - 2013, 17,426 participants completed a repeat dietary assessment using the same questionnaire as baseline, with a median follow-up time of 4.4 (3.7–5.0) years (27). The same criteria was used to classify participants into the diet groups at resurvey.

### References

1. Butler TL, Fraser GE, Beeson WL, Knutsen SF, Herring RP, Chan J, et al. Cohort Profile: The Adventist Health Study-2 (AHS-2). International Journal of Epidemiology. 2007;37(2):260-5.
2. Kondal D, Patel SA, Ali MK, Mohan D, Rautela G, Gujral UP, et al. Cohort profile: the Center for cArdiometabolic Risk Reduction in South Asia (CARRS). Int J Epidemiol. 2022;51(6):e358-e71.
3. Nair M, Ali MK, Ajay VS, Shivashankar R, Mohan V, Pradeepa R, et al. CARRS Surveillance study: design and methods to assess burdens from multiple perspectives. BMC Public Health. 2012;12(1):701.
4. Davey GK, Spencer EA, Appleby PN, Allen NE, Knox KH, Key TJ. EPIC-Oxford: lifestyle characteristics and nutrient intakes in a cohort of 33 883 meat-eaters and 31 546 non meat-eaters in the UK. Public Health Nutr. 2003;6(3):259-69.
5. Appleby PN, Thorogood M, Mann JI, Key TJ. The Oxford Vegetarian Study: an overview. Am J Clin Nutr. 1999;70(3 Suppl):525s-31s.
6. Chiu THT, Huang HY, Chen KJ, Wu YR, Chiu JPC, Li YH, et al. Relative validity and reproducibility of a quantitative FFQ for assessing nutrient intakes of vegetarians in Taiwan. Public Health Nutr. 2014;17(7):1459-66.
7. Cade JE, Burley VJ, Alwan NA, Hutchinson J, Hancock N, Morris MA, et al. Cohort Profile: The UK Women's Cohort Study (UKWCS). International journal of epidemiology. 2017;46(2):e11-e.
8. Chen Z, Chen J, Collins R, Guo Y, Peto R, Wu F, Li L. China Kadoorie Biobank of 0.5 million people: survey methods, baseline characteristics and long-term follow-up. Int J Epidemiol. 2011;40(6):1652-66.
9. Green J, Reeves GK, Floud S, Barnes I, Cairns BJ, Gathani T, et al. Cohort Profile: the Million Women Study. International Journal of Epidemiology. 2018;48(1):28-9e.
10. Key TJ, Balkwill A, Bradbury KE, Reeves GK, Kuan AS, Simpson RF, et al. Foods, macronutrients and breast cancer risk in postmenopausal women: a large UK cohort. Int J Epidemiol. 2018;48(2):489-500.
11. Greenwood DC, Hardie LJ, Frost GS, Alwan NA, Bradbury KE, Carter M, et al. Validation of the Oxford WebQ Online 24-Hour Dietary Questionnaire Using Biomarkers. Am J Epidemiol. 2019;188(10):1858-67.
12. Schatzkin A, Subar AF, Thompson FE, Harlan LC, Tangrea J, Hollenbeck AR, et al. Design and serendipity in establishing a large cohort with wide dietary intake distributions : the National Institutes of Health-American Association of Retired Persons Diet and Health Study. Am J Epidemiol. 2001;154(12):1119-25.
13. Sudlow C, Gallacher J, Allen N, Beral V, Burton P, Danesh J, et al. UK biobank: an open access resource for identifying the causes of a wide range of complex diseases of middle and old age. PLoS Med. 2015;12(3):e1001779.
14. Singh PN, Fraser GE, Knutsen SF, Lindsted KD, Bennett HW. Validity of a physical activity questionnaire among African-American Seventh-day Adventists. Med Sci Sports Exerc. 2001;33(3):468-75.
15. Du HD, Bennett D, Li LM, Whitlock G, Guo Y, Collins R, et al. Physical activity and sedentary leisure time and their associations with BMI, waist circumference, and percentage body fat in 0.5 million adults: the China Kadoorie Biobank study. Am J Clin Nutr. 2013;97(3):487-96.
16. Sarri G, Davies M, Lumsden MA, Grp GD. Diagnosis and management of menopause: summary of NICE guidance. BMJ. 2015;351.
17. Palacios S, Henderson VW, Siseles N, Tan D, Villaseca P. Age of menopause and impact of climacteric symptoms by geographical region. Climacteric. 2010;13(5):419-28.
18. Ahuja M. Age of menopause and determinants of menopause age: A PAN India survey by IMS. J Midlife Health. 2016;7(3):126-31.
19. Wang M, Kartsonaki C, Guo Y, Lv J, Gan W, Chen Z-M, et al. Factors related to age at natural menopause in China: results from the China Kadoorie Biobank. Menopause. 2021;28(10):1130-42.
20. Jaceldo-Siegl K, Fan J, Sabate J, Knutsen SF, Haddad E, Beeson WL, et al. Race-specific validation of food intake obtained from a comprehensive FFQ: the Adventist Health Study-2. Public Health Nutr. 2011;14(11):1988-97.

21. Tong TYN, Appleby PN, Bradbury KE, Perez-Cornago A, Travis RC, Clarke R, Key TJ. Risks of ischaemic heart disease and stroke in meat eaters, fish eaters, and vegetarians over 18 years of follow-up: results from the prospective EPIC-Oxford study. BMJ. 2019;366:l4897.
22. Papier K, Appleby PN, Fensom GK, Knuppel A, Perez-Cornago A, Schmidt JA, et al. Vegetarian diets and risk of hospitalisation or death with diabetes in British adults: results from the EPIC-Oxford study. Nutr Diabetes. 2019;9.
23. Cade JE, Burley VJ, Greenwood DC, G UWsCSS. The UK Women's Cohort Study: comparison of vegetarians, fish-eaters and meat-eaters. Public Health Nutr. 2004;7(7):871-8.
24. Qin CX, Guo Y, Pei P, Du HD, Yang L, Chen YP, et al. The relative validity and reproducibility of food frequency questionnaires in the China Kadoorie Biobank study. Nutrients. 2022;14(4).
25. Roddam AW, Spencer E, Banks E, Beral V, Reeves G, Appleby P, et al. Reproducibility of a short semi-quantitative food group questionnaire and its performance in estimating nutrient intake compared with a 7-day diet diary in the Million Women Study. Public Health Nutr. 2005;8(2):201-13.
26. Thompson FE, Kipnis V, Midthune D, Freedman LS, Carroll RJ, Subar AF, et al. Performance of a food-frequency questionnaire in the US NIH-AARP (National Institutes of Health-American Association of Retired Persons) Diet and Health Study. Public Health Nutr. 2008;11(2):183-95.
27. Bradbury KE, Young HJ, Guo WJ, Key TJ. Dietary assessment in UK Biobank: an evaluation of the performance of the touchscreen dietary questionnaire. J Nutr Sci. 2018;7.
